# AI-Augmented Clinical Decision Support in a Patient-Centric Precision Oncology Registry

**DOI:** 10.1101/2022.03.14.22272390

**Authors:** Mark Shapiro, Timothy J. Stuhlmiller, Bryan Federowicz, William Hoos, Asher Wasserman, Glenn Kramer, Zach Kaufman, Don Chuyka, Julie C. Friedland, Bill Mahoney, Al Musella, Mika Newton, Zachary Osking, J. M. Tenenbaum, Kenny K. Wong, Santosh Kesari, Jeff Shrager

## Abstract

**Purpose:** xDECIDE is a clinical decision support system, accessed through a web portal and powered by a “Human-AI Team”, that offers oncology healthcare providers a set of treatment options personalized for their cancer patients, and provides outcomes tracking through an observational research protocol. This article describes the xDECIDE process and the AI-assisted technologies that ingest semi-structured electronic medical records to identify and then standardize clinico-genomic features, generate a structured personal health record (PHR), and produce ranked treatment options based on clinical evidence, expert insights, and the real world evidence generated within the system itself.

**Method:** Patients may directly enroll in the IRB-approved pan-cancer XCELSIOR registry (NCT03793088). Patient consent permits data aggregation, continuous learning from clinical outcomes, and sharing of limited datasets within the research team. Assisted by numerous AI-based technologies, the xDECIDE team aggregates and processes patients’ electronic medical records, and applies multiple levels of natural language processing (NLP) and machine learning to generate a structured case summary and a standardized list of patient features. Next a ranked list of treatment options is created by an ensemble of AI-based models, called xCORE. The output of xCORE is reviewed by molecular pharmacologists and expert oncologists in a virtual tumor board (VTB). Finally a report is produced that includes a ranked list of treatment options and supporting scientific and medical rationales. Treating physicians can use an interactive portal to view all aspects of these data and associated reports, and to continuously monitor their patients’ information. The xDECIDE system, including xCORE, is self-improving; feedback improves aspects of the process through machine learning, knowledge ingestion, and outcomes-directed process improvement.

**Results:** At the time of writing, over 2,000 patients have enrolled in XCELSIOR, including over 650 with CNS cancers, over 300 with pancreatic cancer, and over 100 each with ovarian, colorectal, and breast cancers. Over 150 VTBs of CNS cancer patients and ∼100 VTBs of pancreatic cancer patients have been performed. In the course of these discussions, ∼450 therapeutic options have been discussed and over 2,000 consensus rationales have been delivered. Further, over 500 treatment rationale statements (“rules”) have been encoded to improve algorithm decision making between similar therapeutics or regimens in the context of individual patient features. We have recently deployed the xCORE AI-based treatment ranking algorithm for validation in real-world patient populations.

**Conclusion:** Clinical decision support through xDECIDE is available for oncologists to utilize in their standard practice of medicine by enrolling a patient in the XCELSIOR trial and accessing xDECIDE through its web portal. This system can help to identify potentially effective treatment options individualized for each patient, based on sophisticated integration of real world evidence, human expert knowledge and opinion, and scientific and clinical publications and databases.

## Introduction: The Promise and Problem of Precision Oncology

The goal of precision oncology is to offer cancer patients treatment options that are tailored to the particulars of their individual disease, as well as to contextual factors, such as their treatment preferences [1]. In oncology, these particulars, which we call the patient’s “profile”, will almost always include a mix of demographics, clinical information, histopathology, and tumor biomarkers such as genetic variants that have diagnostic, prognostic, or treatment significance. Unfortunately, delivering precision oncology is exceedingly complex; thousands of unique combinations of variants have been observed [2], each potentially requiring a different treatment approach, whose knowledge is spread over an enormous volume of papers, guidelines, and databases.

Between 2017 and 2020, the number of genes and variants of significance in the CIViC database increased at compound annual growth rates of 11 and 35 percent, respectively [3]. The number, length, and complexity of NCCN treatment guidelines has been increasing at a compound annual growth rate of approximately 21% for the past 20 years [4]. Today, this equates to approximately 3,000 pages of new guideline information for a general medical oncologist to absorb each year.^1^ When there are no approved treatment options, there may be dozens of relevant clinical trials to choose from. For example, there are currently more than 5,000 actively recruiting interventional trials, and 42 active expanded access programs in cancer across the United States alone.^2^ Researching trials for patients is a time-consuming activity, and only a limited number of cancer patients qualify for interventional clinical trials [5]. It can be difficult to reliably assess patient eligibility for an oncology trial, especially for patients with advanced disease or poor performance status. While an enormous number of potential novel combination regimens could be utilized off-label, implementation of these approaches is limited by the additional time and funding required, and by a lack of information about the validity or applicability of these for specific patients.

The volume of actionable information, the speed of new information creation, and the need to quickly weigh the relative value of this information for a specific patient demands computational support to guide clinical decision making. Yet, in our experience of operating hundreds of virtual tumor boards (VTBs) in advanced cancer, physicians continue to rely on the opinions of trusted colleagues, and web searches that may or may not uncover the most relevant and recent knowledge.

To address this problem we have created the pan-cancer XCELSIOR registry (NCT03793088, Figure 1 and Supplement 1) and associated xDECIDE process. The registry aggregates real-time, real-world clinical outcomes data in the context of observational research. We leverage this data, along with insights and recommendations from leading oncologists, in xDECIDE, an AI-augmented clinical decision support system (CDSS) accessed via the web. xDECIDE is operated by human experts in cancer biology and clinical practice supported by sophisticated algorithms that together enable xDECIDE to surface and rank evidence-based personalized treatment options for any cancer patient in the US.

**Figure 1.**
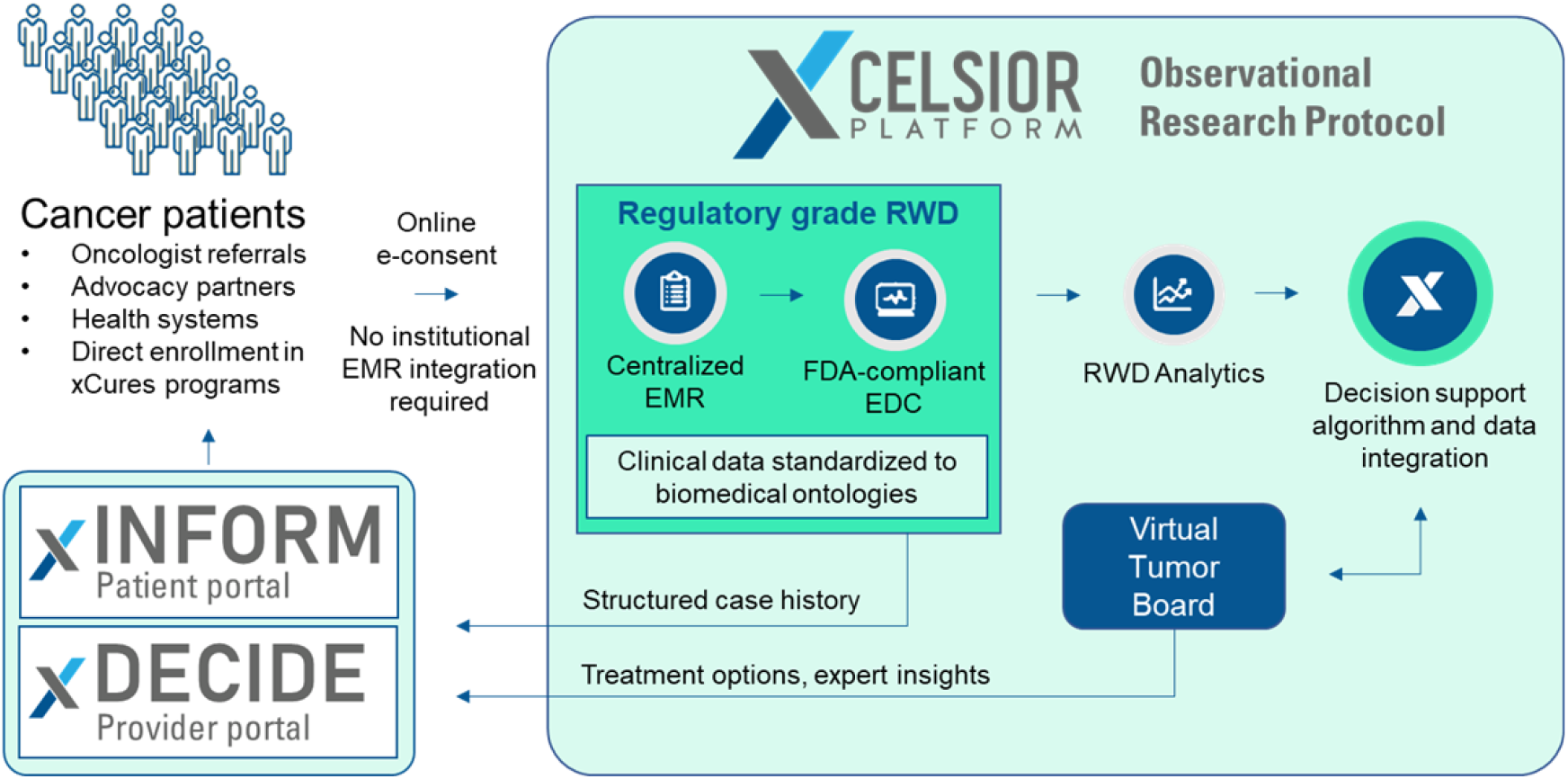
The XCELSIOR study schema (for more detail see Supplement 1)

### Overview of the xDECIDE Clinical Decision Support System

xDECIDE is a clinical decision support system (CDSS) operated by what the FDA calls a “Human-AI Team”^3^ – a collaboration between human experts and AI-based software tools. The human experts include leading clinicians, who review cases individually or together in a panel discussion called a “Virtual Tumor Board” (VTB), patient advocates, and scientists with expertise in molecular pharmacology, cancer biology, and genomics. In addition, data analysts conduct medical record reviews, medical coding, and patient tracking, and computer scientists with expertise in knowledge management, cloud and web engineering, statistics, and machine learning develop the algorithms that support the human processes, making them more accurate, convenient, and efficient so that xDECIDE is validatable, useful, and scalable.

Figure 2 depicts the overall xDECIDE flow. Briefly: Patients can directly enroll themselves, and electronically consent to the XCELSIOR study via an interactive patient portal (xINFORM: https://patient.xcures.com/signup) either on their own accord, or via referral from their treating physician (via https://provider.xcures.com/signup). Following consent, their electronic medical records are aggregated, and a discrete set of data elements are extracted. The coded data receives multiple levels of human and machine review, are mapped to a custom integrated ontology, and are combined with a natural language processing (NLP) pipeline that operates on the unstructured content of the medical records to create the patient’s *digital clinico-genomic profile*. This profile is the input to an ensemble AI algorithm that creates a ranking of potential treatments. These are reviewed by an internal team of PhD scientists with expertise in molecular pharmacology and cancer biology, and discussed, if necessary, in a VTB.

**Figure 2.**
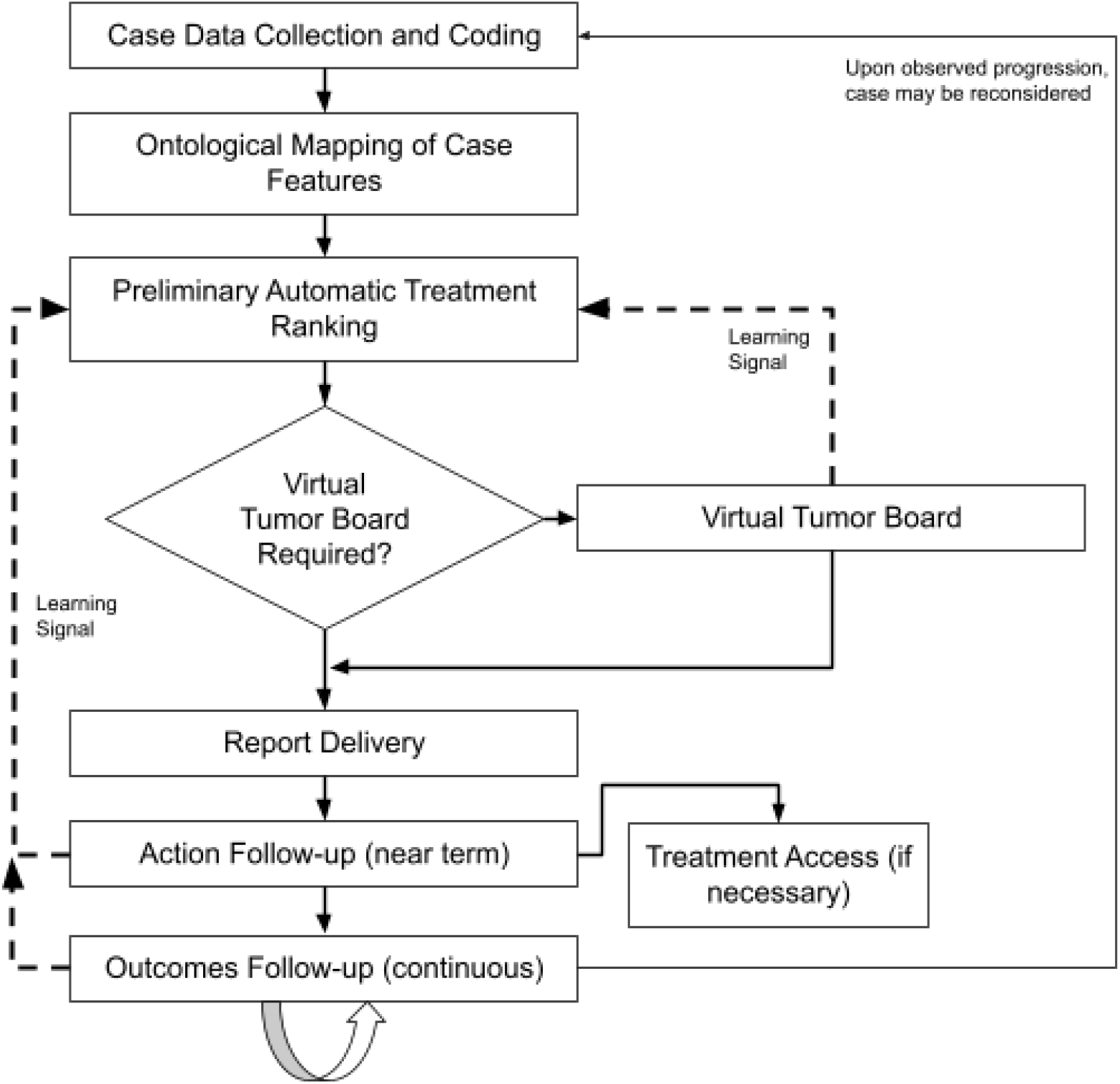
Overview of the CDSS flow. Dashed lines represent learning signals fed back to the algorithm to tune hyperparameters. Described in text.

An interactive online report of the resulting options, including scientific/medical rationales and relevant clinical citations, is simultaneously delivered to the oncologist and patient via the web-based xDECIDE and xINFORM portals, respectively. These options might include approved treatments, off label treatments, clinical trials, experimental treatments accessed via expanded access or Right-to-try as well as combinations of these. Oncologists and patients can interact via the portal to indicate their treatment decision. If they have chosen an expanded access or off-label treatment, treatment access support by internal staff is triggered. Oncologists and patients can continue to interact with the platform to update preferences and history, and to request updates or additional treatment options for consideration. Performance feedback at a number of points serves as data for Machine Learning (ML), which continuously improves the AI-based treatment ranking algorithm (dashed lines in Figure 1).

In what follows, we describe the CDSS subprocesses in more detail, identifying instances where human decision-making is augmented by machine learning, automation, and other technologies. The detailed algorithms and performance analyses are described in separate papers, in preparation.

### Case Data Collection and Coding

After a patient has consent into the XCELSIOR protocol, their electronic medical records including diagnosis pathology, clinic notes, radiology reports, and molecular profiling reports, among other records, are gathered and classified by document type and visit date. This, and all subsequent analysis take place in a 21 CFR Part 11-compliant database (Figure 3).

**Figure 3.**
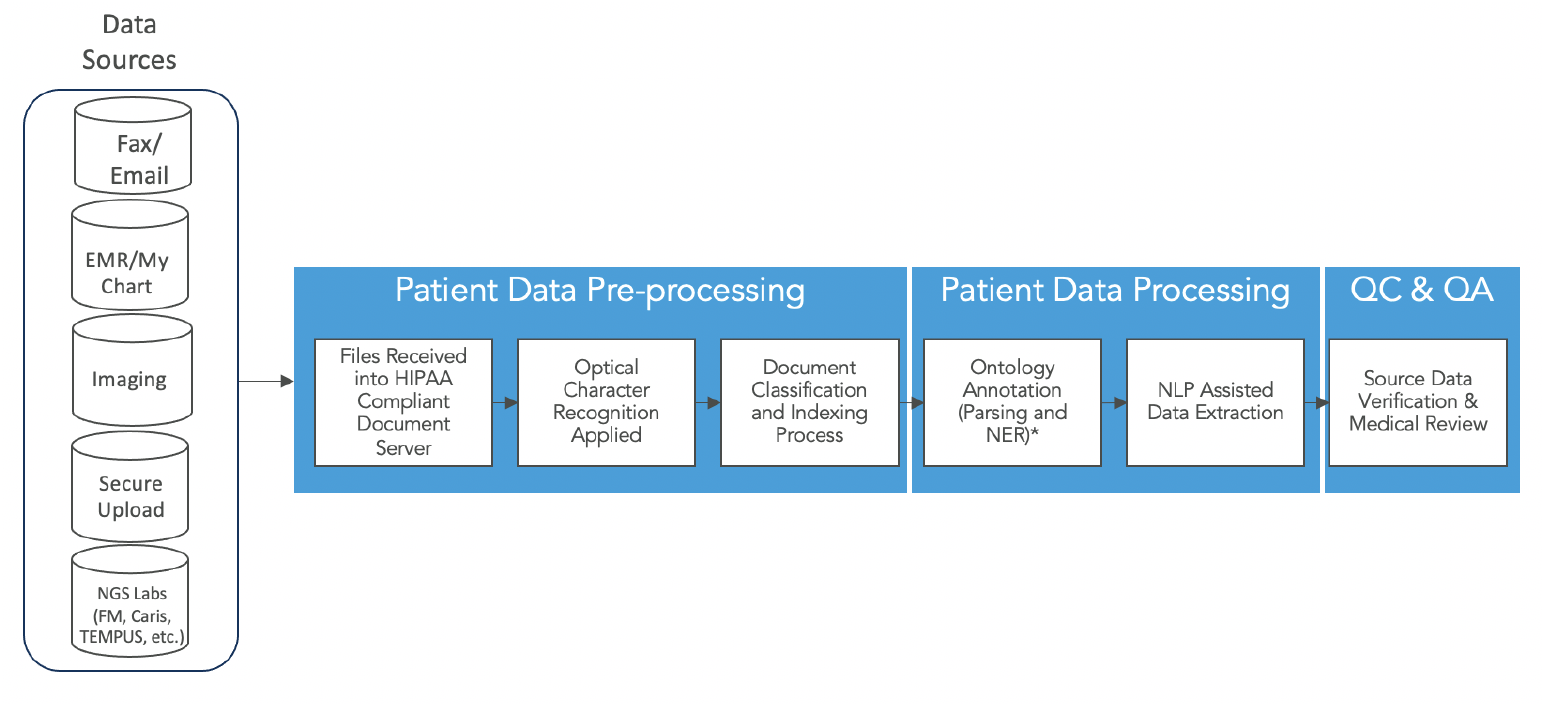
Medical record sources and storage.

The data are imported into an abstraction and annotation tool that subjects them to optical character recognition (OCR) and NLP, including a named-entity recognition (NER) pipeline specialized for cancer and cancer-related features. This facilitates the identification, coding, and mapping of information from the raw medical records into discrete data elements based on custom Case Report Forms (CRFs), modeled on standards including mCODE [19] and CDISC CDASH [20]. CRF customization includes fields for primary and secondary cancer diagnoses, tumor stage/grade, therapy regimens and their event dates (e.g., start/stop), circulating tumor marker values, and biomarker findings, among others (Figure 4). Data elements are source-verified, and direct links to elements in the source medical records, including the precise coordinates on the PDF, where possible.

**Figure 4.**
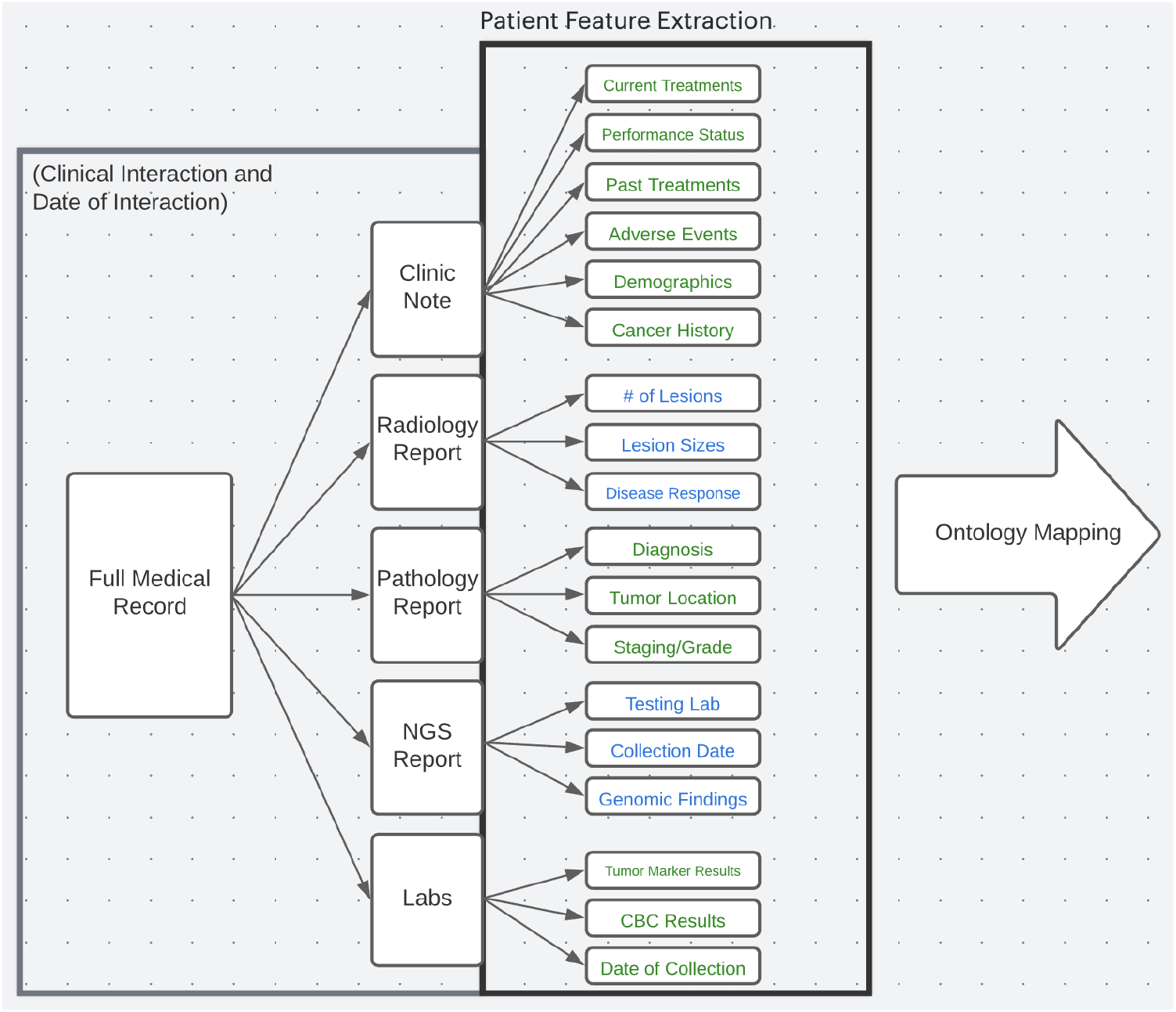
Specific categories of data extracts from medical records and passed to ontology mapping.

Data collection from medical records is overseen by human data abstractors trained in using our OCR and NER systems. Structured data elements are then subjected to a secondary review and Source Data Verification (SDV) to ensure accuracy. A final medical review is conducted to ensure case completeness, identification of any missing data elements or key facets of the individual case, and appropriate codings for any newly identified terms.

### Ontological Mapping of Case Features

The record collection and coding process extracts two primary kinds of data: structured data elements, and natural language from unstructured components of medical records (patient interval history summaries, physician treatment plans, radiology findings and impressions, among others). The structured content is standardized to patient clinico-genomic features according to a custom ontology (xOntology), which we create through integration of several databases, including the NCI Thesaurus [7] (“NCIT”), CIViC [3], MedRA [8], LOINC [9], SNOMED [10], NCBI Entrez Gene [11], UniProt [12], and additional customized internal ontologies, to provide a harmonized set of patient clinico-genomic features suitable for algorithm ingestion. Figure 5 depicts a small portion of the xOntology which, at this writing, contains over 117,000 distinct concepts and an order of magnitude more synonyms or aliases. Among enrolled patients, over 14,000 unique, unnormalized terms from medical records have been coded approximately 7,500 normalized terms across the domains of disease, treatments, labs, and genomics.

**Figure 5.**
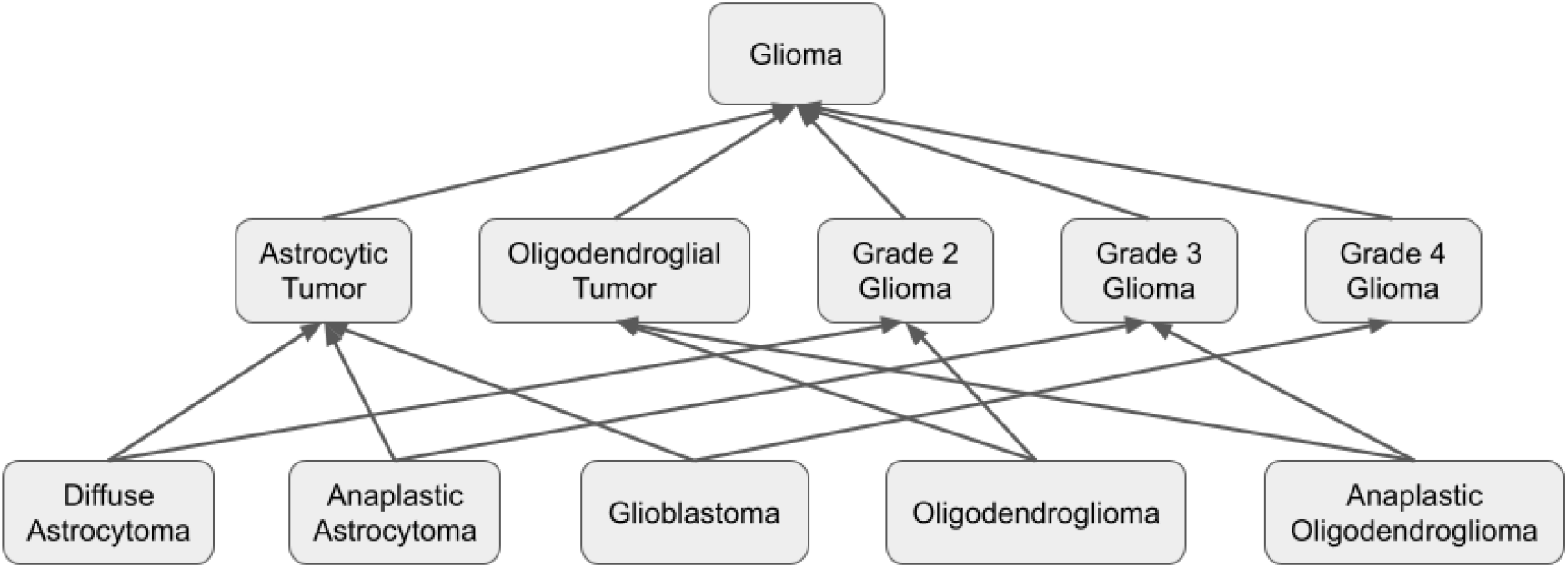
Depiction of a small portion of the xOntology concept graph, adopted from the NCI thesaurus. Upward arrows can be read as: “Tail (more specific) Concept **<u>is a type of</u>** Head (more general) Concept”, for example: “Glioblastoma” is a type of “Astrocytic Tumor”. Each term also allows for standard and/or common synonyms. Concepts can have multiple upper (more general) concepts, allowing for groupings by multiple facets (e.g., disease by cell type or by grade).

The xOntology continues to evolve as the source ontologies evolve, and as new concepts are discovered, either in incoming medical records or from review of the medical literature and other sources, such as clinicaltrials.gov [13]. Ontological terms are added from new instances in patient medical records on medical review - for example to capture a novel short-hand description of a comorbidity, or acronym for a treatment regimen that is not currently represented in the xOntology. Disease-specific concepts or patient features and their underlying ontology terms are also identified from natural language in treatment rationales from clinicians during case review and tumor board discussions. This will be described in more detail below in the section on learning.

### The Options Library and the xCORE Ranking Algorithm

The final output of xDECIDE is a report listing top-ranked treatment options from the xDECIDE Options Library, a comprehensive list of all plausible cancer treatment regimens that includes a standardized coding of abbreviations and aliases of interventions and treatment combinations. For example, “gem/abrax” is coded to the combination of “gemcitabine” and “nab-paclitaxel”. At a minimum, any option in the library has been used previously for treatment of human cancer patients. The sources for this library include (1) individual treatments and combination regimens that form the arms of all interventional cancer clinical trials abstracted from clinicaltrials.gov [13], (2) published case reports, (3) patient treatments observed in the XCELSIOR registry, and (4) novel regimens proposed during tumor board reviews by clinicians with real-world experience. Each option consists of three components, (1) a normalized nomenclature of the component interventions, for example, “ipilimumab, nivolumab, radiation therapy,” (2) scientific/medical rationale(s) derived from past cases and/or published literature, and (3) an access mechanism being one of: standard-of-care (SOC), clinical trial, off-label, expanded access, or diagnostic. For options based in clinical trials, locations and recruiting status are maintained up-to-date according to clinicaltrials.gov [13].

The Options Library consists of ∼12,000 therapeutic options involving over 4,000 discrete interventions alone or in combination. These options are derived from the arms of over 15,000 interventional oncology clinical trials over the past decade, of which ∼5,000 are currently recruiting and available as options to present in reports, as well as regimens observed in the clinical care of patients in the XCELSIOR registry. For example, in the CNS cancer cohort we cataloged 344 distinct regimens used clinically across all lines of therapy.

A case feature vector representing a patient’s digital clinico-genomic profile, generated during case preparation and mapped through the xOntology as described above, is used to query the Options Library via the xCORE treatment ranking algorithm, summarized in Figure 6. The xOntology terms, and combination of terms, are translated to 977 unique features across all CNS VTBs, and 1859 when including inferred features. Principal Components Analysis reduces these to 153 patient feature dimensions which serve as the input to xCORE.

**Figure 6.**
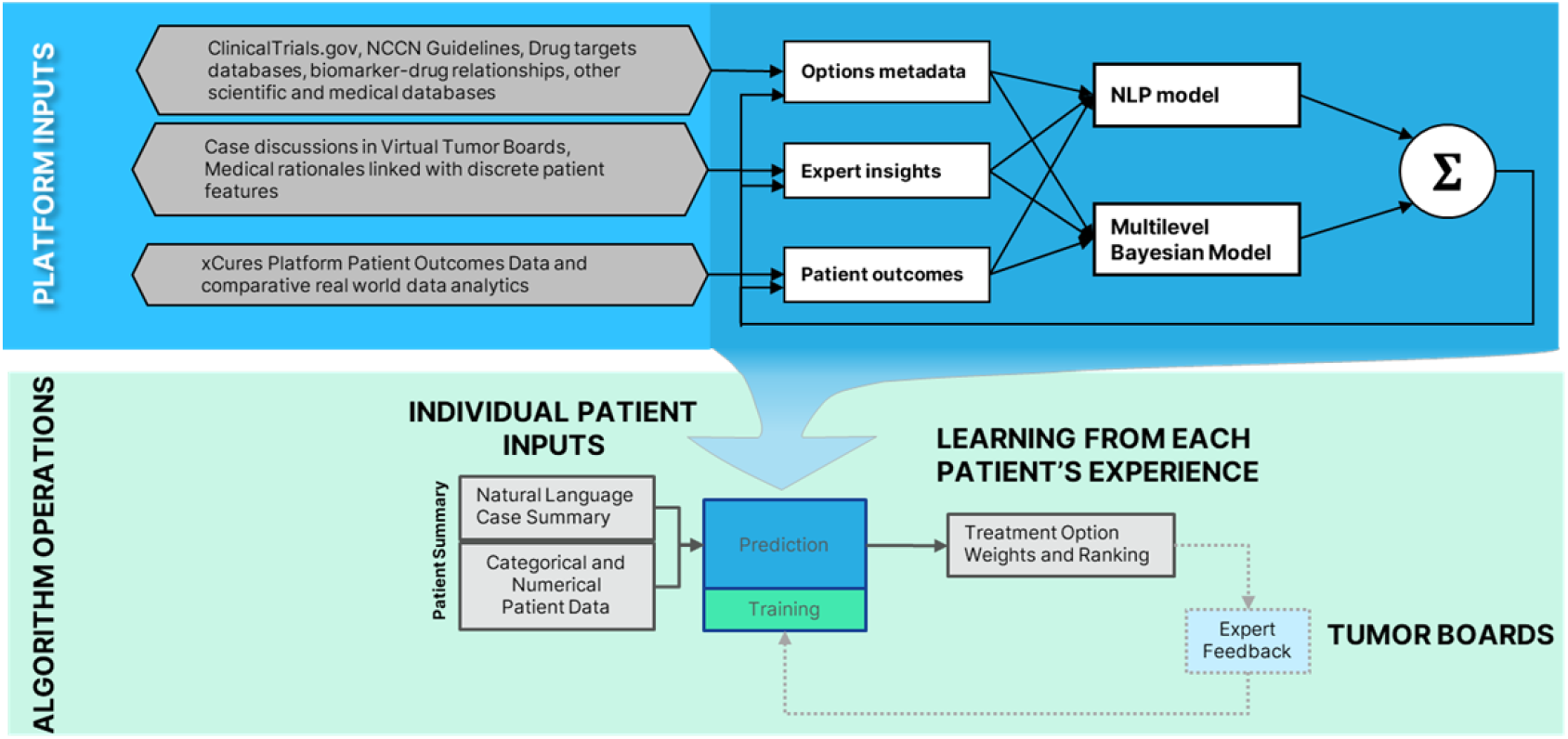
Overall architecture of the xCORE algorithm components and integrations with patient case summaries and tumor board review.

xCORE is an ensemble machine learning system comprising several submodels and a weighted integrator. Submodels are based on multiple platform inputs: (1) natural language in clinical trials linked to treatment regimens and cancer types under investigation, (2) biomarker-drug relationships integrating structured alterations from molecular profiling with annotated databases such as CiViC and the drug-gene interaction database (DGIdb [14]), (3) consensus option rankings from VTBs, (4) rationale statements supporting or opposing the use of specific treatments for discrete patient features, and (5) actual clinical outcomes of patients collected according to the XCELSIOR study. The case feature vector is independently distributed to these submodels. Each submodel scores each treatment and these scores are subsequently combined into an integrated vector of treatment scores (Figure 7). The integration algorithm and each submodel have a set of parameters, some of which are adjusted through learning, described below and in more detail in the paper in preparation (Wasserman, et al., in prep).

**Figure 7.**
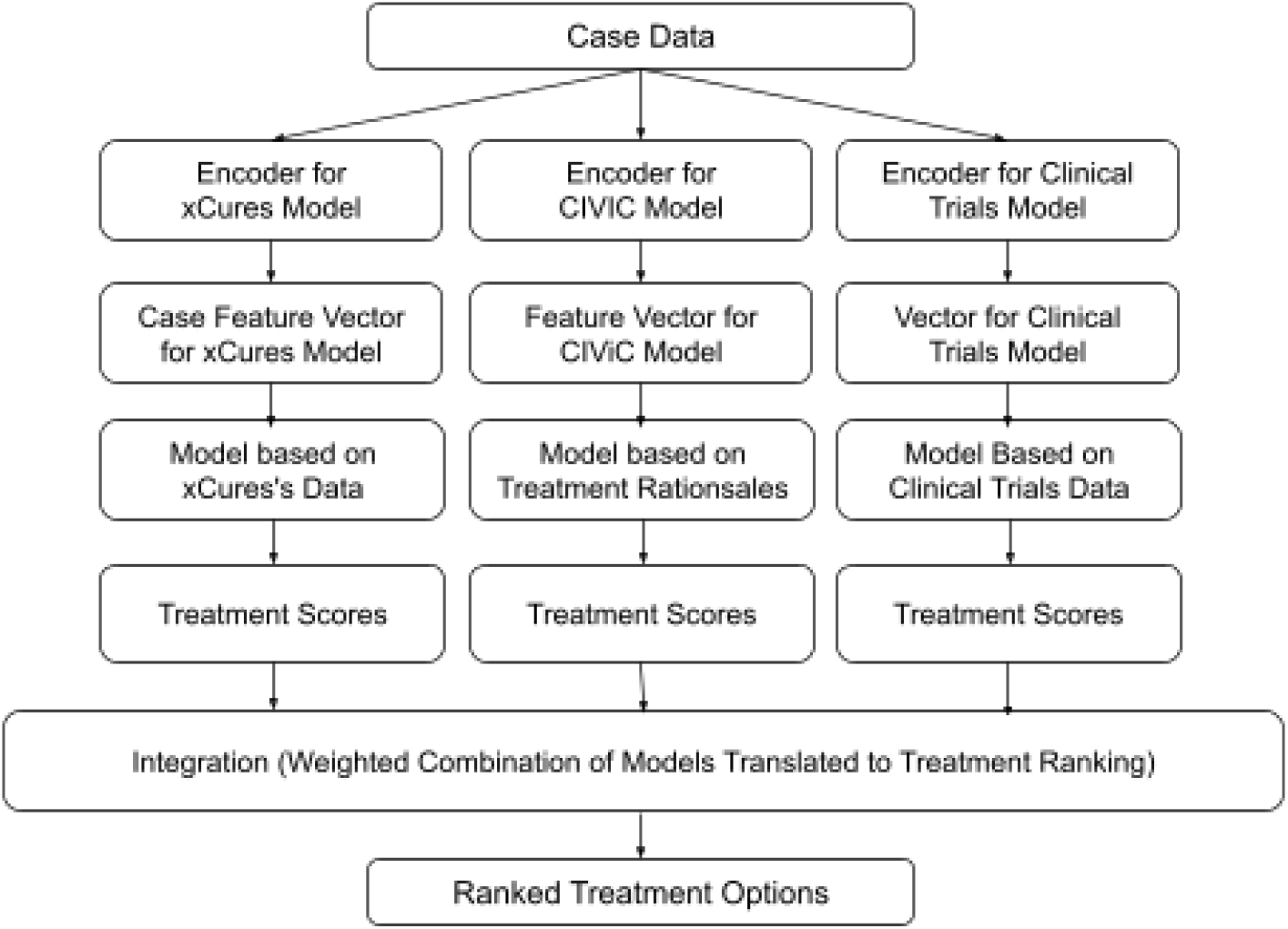
The xCORE Cancer Options Ranking Engine. Case data is encoded into model-specific feature vectors by separate encoders for each of the three models. The feature vectors are passed separately through each model, producing separate treatment feature vector predictions. These are then combined by an integrator, resulting in a final treatment option ranking.

### Virtual Tumor Boards

The output of the xCORE algorithm – patient features and ranked treatment options – are next reviewed by internal PhD scientists, and discussed in a Virtual Tumor Board (VTB). VTBs consist of 3-5 leading clinicians and 2-3 internal PhDs with expertise in molecular pharmacology and cancer biology. Clinicians are practicing medical oncologists or specialists in a particular field of oncology, such as neuro-oncologist/neurologists, who hold an M.D. and/or Ph.D. These physicians are regarded as experts in their field based on their current appointment at top US academic research institutions, generally as division leaders, and/or participation or leadership with national committees on cancer treatment guidelines (e.g., NCCN). They are actively involved in clinical research, serve as investigators on numerous clinical trials for novel therapeutics or treatment regimens, have a strong publication record in clinical and/or preclinical research, and hold expertise in one or more aspects of treatment for their disease specialty, for example immunotherapies, targeted therapies, or particular signaling pathways.

The VTB reviews the structured and unstructured patient case data and xCORE output of potential treatment options in either a synchronous virtual meeting or asynchronous dialogue, and determines the final recommendations. The patient clinical case summary is available, including the patient’s past cancer history, tumor genetics/biomarkers, results from any additional diagnostic testing, comorbidities, concomitant medications, and family history, as well as the most recent imaging report summary. In some instances, additional inputs are available, for example, from alternative genomics-based algorithms or cell- and organoid-based functional profiling assays. Importantly, statements (“rules”) supporting or opposing specific options, together with the particular clinico-genomic patient features that weighed into that decision, are solicited and coded for future learning in the xCORE algorithm of the format: Patient feature X supports/opposes Option Y for Condition Z [15]. Ultimately, the case reviewers – whether one expert or a panel of external experts in a VTB – create a consensus recommendation of ranked therapeutic options.

### Report Finalization, Delivery, and Followup

Internal PhD-level staff utilize various dashboards to examine, manipulate, and combine all of the foregoing into a report. Supplement 2 depicts some of these data management dashboards.

In this stage, treatment recommendations are augmented with rationales that help the recipient understand the underlying logic of the recommendations. Rationales contain individual patient features that relate to the recommendation of the option presented, as well as the most relevant published clinical evidence supporting the use of that option. The report is delivered simultaneously to the oncologist and patient, both in PDF format, through the xDECIDE and xINFORM portals, respectively. Supplement 3 depicts a complete sample final report, as well as a part of the information that patients and physicians can access about a case through the xDECIDE and xINFORM interactive web portals.

After report delivery, patients and their oncologist can interact with the platform to indicate their chosen treatment option and provide their reasoning for pursuing the option. This provides a rapid learning signal for the clinical utility of xDECIDE, and also provides natural language for text mining for supporting rationales and associated patient features.

If a selected option is available via off-label or expanded access, outreach is initiated to the treating physician to offer access support, including clinical evidence citations for inclusion in prior approval requests, interaction with pharmaceutical companies for expanded access requests, and/or regulatory support for single patient protocol preparation.

The patient’s medical records are periodically requested and processed to close the learning loop, but can also be manually uploaded by the patient or their oncologist via the xINFORM or xDECIDE portal. If a change in the patient’s dataset alters the xCORE algorithm’s output, a new series of treatment options is created and presented to the patient and their oncologist.

### Machine Learning and Continuous Process Improvement

Many of xDECIDE’s internal processes are self-improving through supervised learning, in which the outputs of the process are checked by either human experts or through outcomes metrics, and these signals are used to improve the process.The following signals are actively and continuously integrated into the learning system:

- Unmapped or missing terms identified during intake of electronic medical records, or identified by VTBs, are added to the medical record pipeline and/or xOntology, either as new terms or as aliases of existing terms.
- Final consensus treatment recommendations arising from the VTB process are used to directly train the ranker. Preferential weighting in algorithm training is given to cases reviewed by external oncologists versus internal scientists.
- Treatment choices indicated by the patient or physician within their respective portals xINFORM and xDECIDE provide a rapid clinical utility signal.
- Rationales that stem from expert statements in the context of individual patient cases can suggest novel terms or rules. (More detail on this appears below.)
- Clinical outcomes, e.g., changes in tumor load, clinically-relevant biomarkers, patient or clinician-reported functional status, emergence of adverse effects, and progression, treatment-changes, and survival are among the most important learning signals, but are also the most delayed.
- Real world evidence generated on the XCELSIOR platform from cohort-level analyses also provides a potential source of aggregate terms and/or rules.

### The Special Role of Treatment Rationales in Learning

As was mentioned above, rationales commonly arise in VTB discussion, and sometimes in the context of patient choices. Rationales are important because they provide a potential explanation regarding *why* a particular option was chosen or ranked higher, or why a particular option was deleted (e.g., contra-indicated) or ranked lower [15,16]. Rationales can be derived from the discussion of biomarkers or other patient features such as the presence or number of metastases in a particular organ. If these are novel to xDECIDE – that is, they are not included in the xOntology or were not being extracted from the medical records – these will be considered for inclusion in the extraction and ontology mapping processes. This approach for precision-oncology decision-making mirrors the inclusion/exclusion of clinical trials, but the xDECIDE system is dynamically learning criteria supporting or excluding treatments at an individual level, and learning to generalize these to other patients.

### Status Summary and Results

At the time of writing, over 2,000 patients have enrolled in XCELSIOR from more than 450 cancer centers and oncology practices across the United States. This includes over 650 patients with CNS cancers, over 300 with pancreatic cancer, and over 100 each with ovarian, colorectal, and breast cancers. The XCELSIOR study database contains over 14,000 unique verbatim terms abstracted from more than 750,000 pages of participants’ medical records. This data has been coded to ∼7,500 standardized terms across the domains of disease, treatments, labs, and genomics in a structured, event-driven CRF. These terms, plus ontology-implied terms, and the combinations of terms thereof are translated to an approximately 150-dimensional patient profile for ingestion by the xCORE clinical decision support algorithm. Over 150 VTBs of CNS cancer patients and ∼100 VTBs of pancreatic cancer patients have been performed. In the course of these discussions, ∼450 therapeutic options have been discussed and over 2,000 consensus rationales have been delivered. Clinical trial participation was the most common access mechanism presented; 38% of all options presented were clinical trials, with 76% of patients recommended at least one clinical trial following VTBs. Off-label, standard-of-care, and expanded access options were presented at an overall rate of 29%, 19%, and 11%, respectively with the remaining options consisting of Right-to-try and diagnostic approaches. From VTB deliberations, over 500 treatment rationale statements (“rules”) have been encoded to improve algorithm decision making between similar therapeutics or regimens in the context of individual patient features. Data produced from VTBs linked with patient features extracted and normalized from medical records form the basis for the expert-based submodel of xCORE. Our recent deployment of xCORE for validation in real-world patient populations is under the continuing oversight of experts and VTBs.

## Conclusions

We have described how the xDECIDE precision oncology process works, component-by-component. XCELSIOR offers treating oncologists a platform to participate in nationwide observational research and the Provider Portal xDECIDE delivers immediate value for enrolled patients in the form of structured longitudinal case summaries abstracted from their health records and personalized treatment options for consideration based on expert insights. The platform leverages optical character recognition, natural language processing, and machine learning to augment, but not replace, human case review and therapeutic option selection, providing a scalable computational solution to real world precision medicine applications. The ultimate goal of xDECIDE and the xCORE algorithm is to deliver treatment option recommendations whose outcomes are equivalent to or better than those of molecular tumor boards at leading institutions. Operating under the XCELSIOR protocol, in the context of observational research, xDECIDE has the potential to reach the scale necessary to undertake long term comparisons of the efficacy of treatments more efficiently than randomized controlled trials (RCTs), or even than current adaptive trial technologies, as suggested in 2013 by Shrager [17], and recently validated by Wasserman et al [18]. The xDECIDE CDSS is currently being submitted to the FDA as a “Software as a Medical Device” (SaMD), with algorithms focused on Central Nervous System (CNS) and pancreatic cancers.

## Supporting information

Supplement 1: XCELSIOR protocol

Supplement 3: Fictitious Treatment Options Report

## Data Availability

Data described are available through partnership with xCures, Inc.

## Acknowledgements

Substantial contributions to the creation of operation of xDECIDE were made by Nick Arora, Shaalan Beg, Nicholas Blondin, David Chang, Zac Cole, Colin FitzGerald, Ekokobe Fonkem, Mike Freed, Sarah Ginn, Chezney Hinton, Sabrina Irizarry, Fabio Iwamoto, Vlod Kalicun, Conan Kinsey, Amy Langville, Hope Lee, Jillian Hattaway Luttman, Erika Vial Monteverdi, Burt Nabors, David Patterson, Chris Porter, Jameson Quinn, Lola Rahib, Jeff Rapp, Charles Redfern, Alanis Sabates, Kamalesh Sankhala, Davendra Sohal, Srikar Srinath, Andrew Swain, Kristina Tadic, Matthew Warner, Eric Wong, and Alayna Wood.

---

**Supplement 1: The XCELSIOR Trial protocol**

(PDF Attached)

**Supplement 2:**
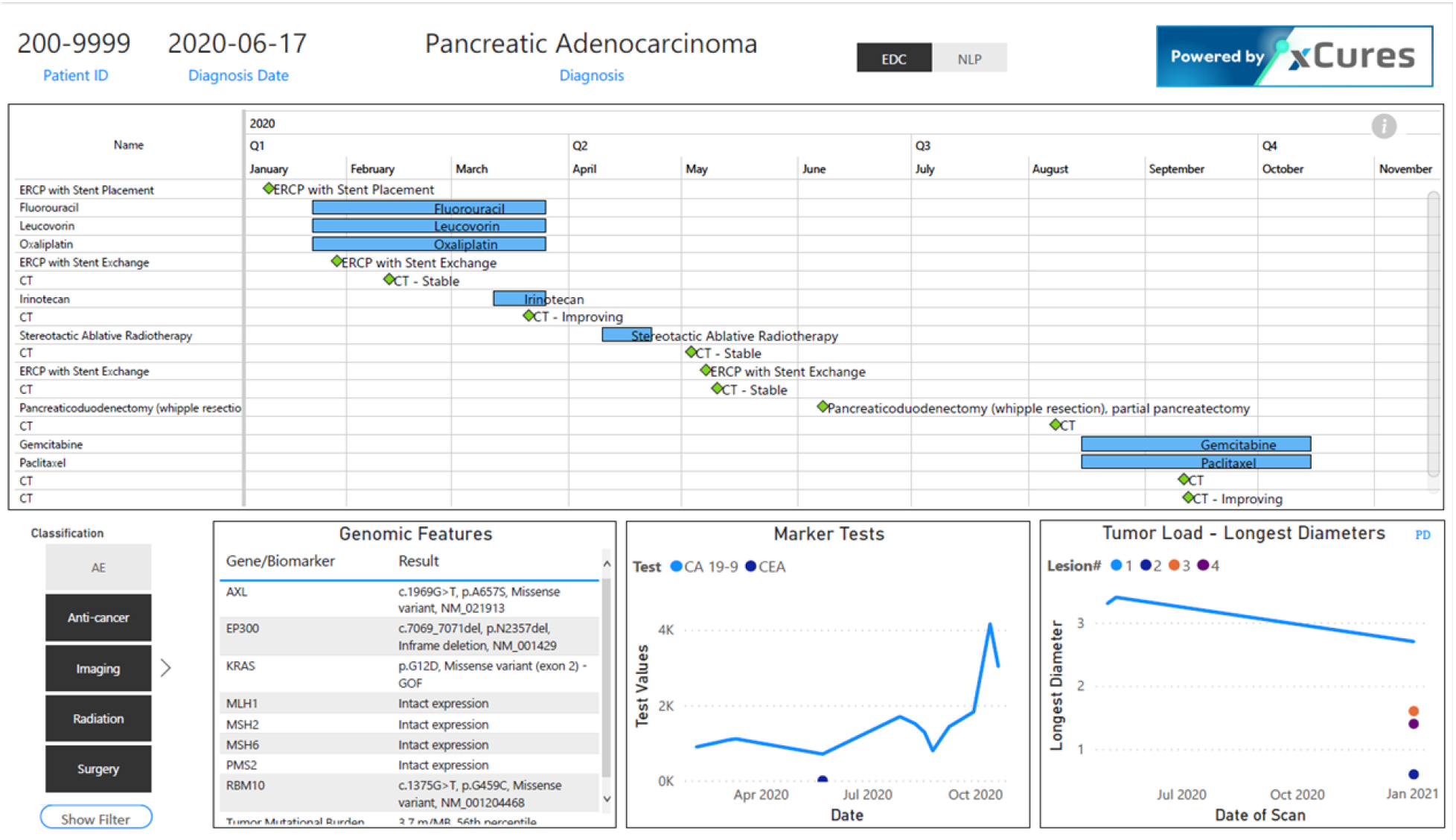
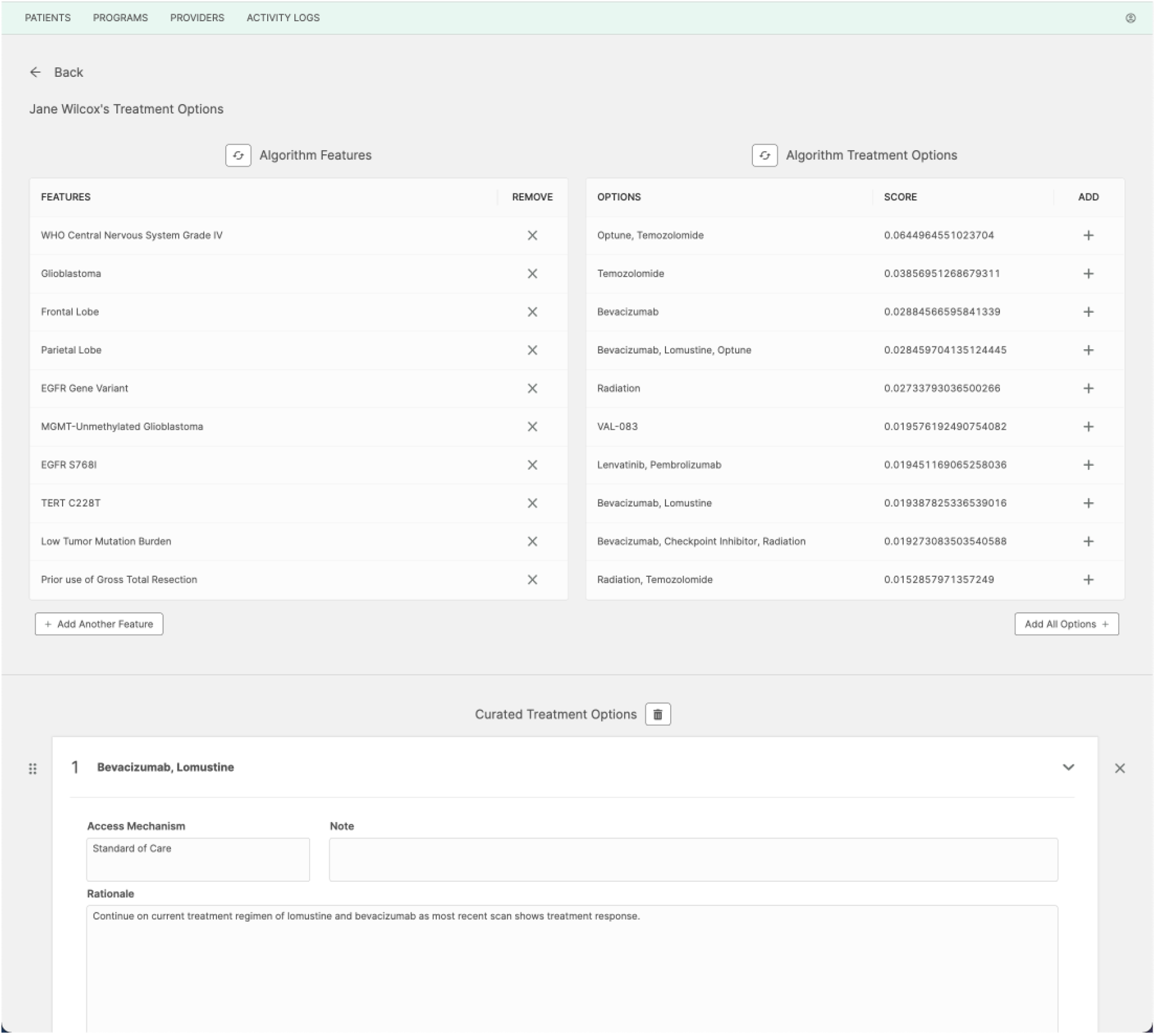
Dashboards. The Patient Case Review dashboard provides an interactive view into the extracted and unstructured data elements from individual patient’s medical records. This serves as an interactive medical chart for quality control, medical review, and case review for options research and tumor board facilitation. (PHI is anonymized.) The Treatment Option Curation dashboard is the Medical Affairs group’s portal into feature extraction, option curation, and the automatic treatment ranker. Through this dashboard internal experts can (a) query the standardized features associated with a patient’s case and update the features if necessary, (b) automatically produce a preliminary rank-ordered set of treatment options for consideration, (c) further curate the options (e.g., adding, removing, reordering, and providing rationales/notes), and (d) release the approved option report to a patient and their provider. (Patient is fictitious.)

**Supplement 3:**
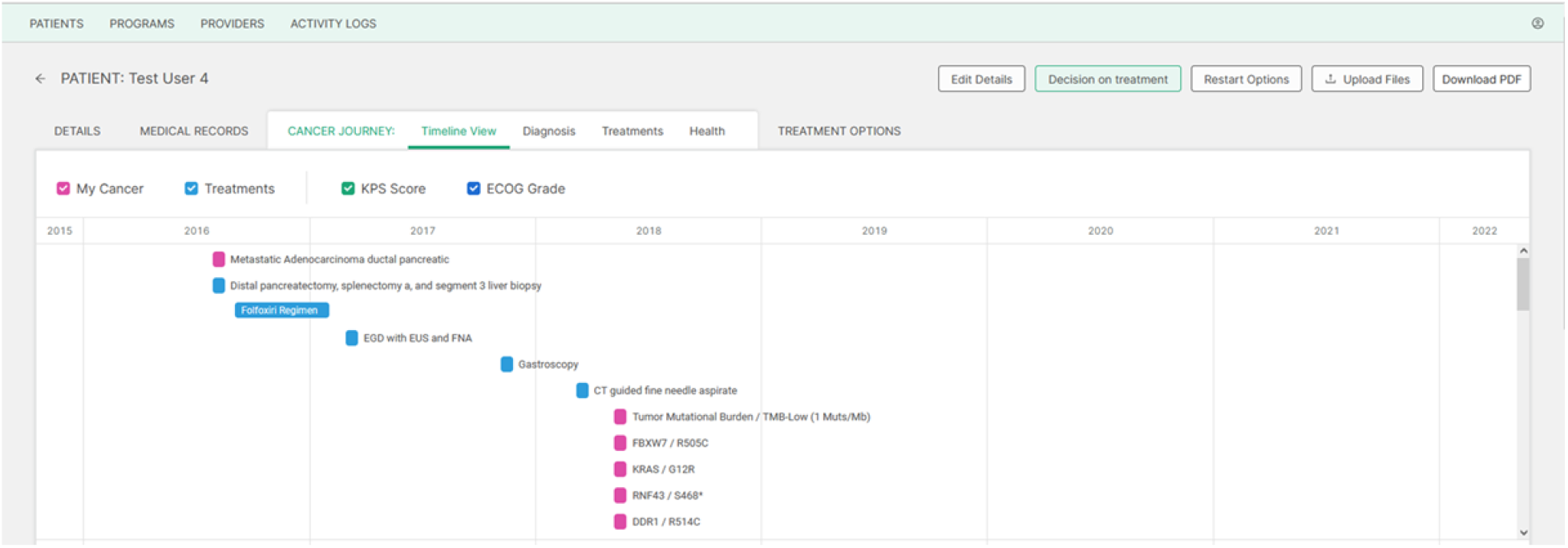
An example of an xDECIDE client report, including treatment options report and cancer journey. Screenshot of the timeline view of the Cancer Journey within the xDECIDE interface as viewed by oncology providers. (Patient is fictitious.) (PDF of sample report for fictitious patient attached)

## Footnotes

1 See Supplement 1 for supporting data analysis

2 Based upon a clinicaltrials.gov [13] search for expanded access programs in cancer, carried out on 2022-03-13.

3 The FDA’s “Good Machine Learning Practice for Medical Device Development: Guiding Principles” [6] reads, in part: “Focus Is Placed on the Performance of the Human-AI Team: Where the model has a ‘human in the loop,’ human factors considerations and the human interpretability of the model outputs are addressed with emphasis on the performance of the Human-AI team, rather than just the performance of the model in isolation.”

## Notes

### Competing Interest Statement

The authors have declared no competing interest.

### Clinical Protocols

https://clinicaltrials.gov/ct2/show/NCT03793088

### Funding Statement

This study was supported by xCures, Inc.

### Author Declarations

The XCELSIOR protocol (NCT03793088) has been approved by the Genetic Alliance IRB (IORG0003358). Details of the ethical considerations are included in the XCELSIOR protocol which appears in full as Supplement #1.

### Summary of Updates

Updated authors and acknowledgements. Added clear indications, where relevant, that figures and reports are of fictitious data. Added URLs where web-based services are described.

## References

1. Shrager J, Tenenbaum JM: Rapid learning precision oncology. Nature Reviews Clinical Oncology 11:109–118, 2014

2. COSMIC, the Catalogue Of Somatic Mutations In Cancer. https://cancer.sanger.ac.uk/cosmic

3. CIViC, Clinical Interpretations of Variants in Cancer. https://civicdb.org/home

4. Kann BH, Johnson SB, Aerts HJWL, et al: Changes in Length and Complexity of Clinical Practice Guidelines in Oncology, 1996-2019. AMA Netw Open, 2020

5. Broadening Clinical Trial Participation. https://www.aacr.org/blog/2017/08/01/broadening-clinical-trial-participation/

6. Good Machine Learning Practice for Medical Device Development: Guiding Principles. https://www.fda.gov/medical-devices/software-medical-device-samd/good-machine-learning-practice-medical-device-development-guiding-principles

7. NCI Thesaurus (NCIt). https://ncithesaurus.nci.nih.gov/ncitbrowser/

8. MedDRA. https://www.meddra.org

9. LOINC. https://loinc.org

10. SNOMED. https://www.snomed.org

11. NCBI Gene. https://www.ncbi.nlm.nih.gov/gene

12. UniProt. https://www.uniprot.org

13. http://ClinicalTrials.gov is a database of privately and publicly funded clinical studies conducted around the world. https://www.clinicaltrials.gov

14. DGIdb: The Drug-Genome Interaction database. https://www.dgidb.org

15. Mocellin S, Shrager J, Scolyer R, et al: Targeted Therapy Database (TTD): a model to match patients’ molecular profile with current knowledge on cancer biology. PLoS ONE 5(8): e11965, 2010

16. Sweetnam C, Mocellin S, Krauthammer M, et al: Prototyping a precision oncology 3.0 rapid learning platform. BMC Bioinformatics 19:341, 2018

17. Shrager J: Theoretical Issues for Global Cumulative Treatment Analysis (GCTA). 1308.1066, 2013.

18. Wasserman A, Musella A, Shapiro M, et al: Virtual Trials: Causally-validated treatment effects efficiently learned from an observational brain cancer registry. MedRxiv/10.1101/2021.06.12.21258409v1, 2021

19. mCODE, minimal Common Oncology Data Elements. http://hl7.org/fhir/us/mcode/index.html

20. CDISC, Clinical Data Interchange Standards Consortium; CDASH, Clinical Data Acquisition Standards Harmonization. https://www.cdisc.org/standards/foundational/cdash

