## Supplement 1: XCELSIOR protocol for "AI-Augmented Clinical Decision Support in a Patient-Centric Precision Oncology Registry"

### XCures/Cancer Commons Enhanced Learning Treatment Selection and Analysis with Outcomes Research (XCELSIOR) Study: A Patient-Centric Platform Trial for Precision Oncology

#### For Registration of Patients, Virtual Tumor Board Operations, and Longitudinal, Observational Registry Data Operations

| | |
|---|---|
| Protocol Number: | $XC^3$-GCTA-2018 |
| Version Number: | v.2.0 |
| Version Date: | 17 March 2021 |
| IND Number: | N/A |
| NCT Identified Number: | NCT03793088 |
| Sponsor: | xCures |
| Funded by: | xCures |
| Central IRB: | Genetic Alliance IRB |
| IORG Number: | IORG0003358 |
| Partner Organizations: | Cancer Commons, The Musella Foundation for Brain Tumor Research and Information, Inc.<br>Pediatric Oncology Experimental Therapeutics Investigators Consortium |

### Summary of Changes from Previous Version:

| Affected Section(s) | Summary of Revisions Made | Rationale |
|---|---|---|
| Cover Page | NCT number added, protocol version and date updated, sponsor, collaborators and partners updated | New partnerships, collaborations and organizational structure |
| Protocol Summary | NCT Number added, protocol version and date updated, sponsor, collaborators and partnerships updated, order of objectives serviced | New partnerships, collaborations and organizational structure |
| Protocol Summary | Definition of recalcitrant cancer updated | As new treatments emerge the 5-year survival continues to change for some cancers. New definition is more general. |
| Protocol Summary | Endpoints updated to include more specific details of PRO instruments being used | Selected more public domain PRO surveys |
| Disease Setting and Trial Context | Clarified areas of focus to include cancer types of specific interest to new partner/collaborators | New partnerships include groups with additional cancer focuses |
| Study Rationale | Added information about organoid and drug sensitivity assays | In addition to NGS tests, tumor boards now may have access to additional ex vivo testing for individualized drug sensitivity |
| General Changes | Role of Sponsor and Partner has been clarified throughout | Since launch the working process between xCures and Cancer Commons has evolved. xCures has assumed the role of funder and sponsor, with associated quality systems, while Cancer Commons is a key partner. |

### Table of Contents

STATEMENT OF COMPLIANCE 6
1 PROTOCOL SUMMARY 7
1.1 Synopsis 7
1.2 Study Schema 10
2 STUDY RATIONALE 11
2.1 Disease Setting and Trial Context 16
2.2 Research Leading to the Proposed Study 17
2.2.1 Cancer Commons, xCures, and the Musella Foundation 17
2.2.2 Precision Medicine and the Role of Molecular Tumor Boards 18
2.2.3 Patient-Centric and “Siteless” Clinical Trials and Registries 20
2.2.4 Expansion of Treatment Options 21
2.2.5 Expansion of Testing Options 22
2.2.6 Artificial Intelligence and Advanced Analytic Methods 23
2.2.7 Incorporating Patient Goals and Values in Treatment Choice 24
2.2.8 Real World Evidence 25
2.2.9 Aggregating N-of-1 Experiments and associated Patient Outcomes 26
2.2.10 Assessment of Study Feasibility 27
2.3 Specific Problem Statement 27
2.3.1 Options and Access 27
2.3.2 Outcomes and Analytics 27
2.4 Goals and Objectives of XCELSIOR 28
2.4.1 Patient Options Core 29
2.4.1.1 VIRTUAL TUMOR BOARD 30
2.4.2 Patient Access Core 31
2.4.2.1 COMPASSIONATE USE CLEARINGHOUSE 31
2.4.2.2 OFF-LABEL AND COMBINATIONS FOR TARGETED THERAPIES 31
2.4.3 Data and Analytics Core 32
2.5 Risk/Benefit Assessment 32
2.5.1 Known Potential Risks 32
2.5.2 Known Potential Benefits 33
2.5.3 Assessment of Potential Risks and Benefits 33
3 OBJECTIVES AND ENDPOINTS 34
4 STUDY DESIGN 35
4.1 Overall Design 35
4.2 Scientific Rationale for Study Design 35
4.3 End of Study Definition 35
5 PATIENT OPTIONS CORE 36
5.1 Registration of Patients 36
5.1.1 Patient Registration Process and Schema 36
5.1.1.1 Patient Registration Process Steps 36
5.1.1.2 Patient Registration Schema 36
5.1.2 Consent/assent and Other Informational Documents Provided to participants 37
5.1.3 Consent Procedures and Documentation 37
5.1.4 Selection of Patients 37
5.1.4.1 Inclusion Criteria 37
5.1.4.2 Exclusion Criteria 38
5.1.4.3 Screen Failures 38
5.2 Virtual Tumor Board Operations 38
5.2.1 Virtual Tumor Board Overview 38
5.2.2 Virtual Tumor Board Process 38

5.2.3 VTB Operations Schema 39
5.2.4 Qualification and Selection of VTB Members 39
5.2.5 VTB Financial Disclosure and Conflict of Interest Policy 39
5.2.6 Relation to vDMC 40
5.2.7 Authorship and Publication Committee 40
6 PATIENT ACCESS SUPPORT CORE 41
6.1 Treatment Access Support 42
6.2 Patient Access Support Schema 43
7 PATIENTS OUTCOMES CORE 44
7.1 Data Handling and Record Keeping 44
7.1.1 Data Management 44
7.1.2 Study Records Retention 44
7.1.3 Source Documents 44
7.1.4 Data Elements for Collection 45
7.2 Anti-Cancer and Other Therapies or Treatments 45
7.2.1 Treatment Switching 45
7.3 Discontinuation of Participation 46
7.3.1 Participant Discontinuation/Withdrawal from the Study 46
7.3.2 Lost to Follow-Up 46
7.4 Virtual Data Monitoring Committee 46
7.4.1 Signal Detection Process and Schema 47
7.4.2 Qualifications and Term for vDMC Members 48
7.5 Adverse Events and Serious Adverse Events 48
7.5.1 Definition of Adverse Events (AE) 48
7.5.2 Definition of Serious Adverse Events (SAE) 49
7.5.3 Relationship to Underlying Disease or Treatment 49
7.5.4 Expectedness 50
7.5.5 Reporting Events 50
7.5.6 Reporting Events to Participants 50
7.6 Unanticipated Problems 50
7.6.1 Definition of Unanticipated Problems (UP) 50
7.6.2 Unanticipated Problem Reporting 50
7.6.3 Reporting Unanticipated Problems to Participants 51
8 ANALYTIC METHODS AND STATISTICAL CONSIDERATIONS 52
8.1 Overview of Analytic Methods 52
8.1.1 Cluster Analysis and Patient Classification 52
8.1.2 Equipoise Options Set and Response Measurement 54
8.1.3 Patient Utility Analysis 55
8.1.4 Analytic Signal Detection 56
8.1.5 Sample Size Considerations 56
8.1.6 Populations for Analyses 56
8.1.7 Statistical Analyses 57
8.1.8 Baseline Descriptive Statistics and Annual Reporting 57
8.1.9 Exploratory and Future Analyses 57
9 SUPPORTING DOCUMENTATION AND CONSIDERATIONS 58
9.1 Regulatory and Ethical Considerations 58
9.1.1 Regulatory Considerations 58
9.1.2 Confidentiality, Privacy, and Data Security 59
9.1.3 Schema and Overview of Services Data Versus Research Data in XCELSIOR 59
9.1.4 Future Use of Stored Data 61
9.1.5 Publication and Data Sharing Policy 61
9.1.6 Conflict of Interest Policy 61
9.1.7 Ethical Considerations 62

9.2 Study Oversight and Other Considerations 62
9.2.1 Financial Considerations 62
9.2.2 Study Discontinuation and Closure 62
9.2.3 Quality Assurance and Quality Control 63
9.2.4 Key Roles and Study Governance 63
9.3 Abbreviations 64
Appendix 1: Informed Consent and Assent Text 71
Appendix 2: Examples of Functional, Quality-of-Life, and Other Response Measures to be Collected 74

#### STATEMENT OF COMPLIANCE

This trial will be carried out in accordance with International Conference on Harmonization Guideline for Good Clinical Practice (ICH GCP) and the following: United States (US) Code of Federal Regulations (CFR) applicable to clinical studies 45 CFR 46, 21 CFR Part 50 and 21 CFR Part 56, and as applicable, 21 CFR Part 312 and 21 CFR 812.

Protocols, informed consent form(s), recruitment materials, and all participant materials will be submitted for Institutional Review Board (IRB) for review and approval. Approval of both the protocol and the consent form will be obtained before any participant is enrolled. Any amendment to the protocol will require review and approval by the IRB before the changes are implemented to the study. In addition, all changes to the consent form will be IRB-approved; a determination will be made regarding whether a new consent needs to be obtained from participants who provided consent, using a previously approved consent form.

### 1 PROTOCOL SUMMARY

#### 1.1 SYNOPSIS

| | |
|---|---|
| **Study Sponsor:** | xCures |
| **Study Title:** | XCures/Cancer Commons Enhanced Learning Treatment Selection and Analysis with Outcomes Research (XCELSIOR) Study: A Patient-Centric Platform Trial for Precision Oncology |
| **Study Acronym:** | XCELSIOR |
| **Study Description:** | XCELSIOR is patient-centric study for the registration of cancer patients, operations of a virtual tumor board, insight capture in clinical decision-making, and collection of longitudinal, observational data in a cancer registry. Patient intake into XCELSIOR will occur through the Cancer Commons Web portal. This includes consent to participate in the data registry, including the collection and review of medical information by a Virtual Tumor Board, generation of patient-specific treatment options with supporting rationale, access to treatment access support services, and inclusion into a registry study that includes safety and efficacy outcomes tracking. Patients will be treated and tracked in their original treatment setting and the data generated will form part of a systematic framework combining expert judgment with artificial intelligence to maximize information gain and improve treatment option set development for individual cancer patients. |
| **Study Number:** | $XC^3$-GCTA-2018 |
| **IND Number:** | N/A |
| **NCT Identified Number:** | NCT03793088 |
| **General Study Design:** | XCELSIOR is a patient-centric treatment and outcomes registry. |
| **Objectives:** | Primary Objectives<br>• To collect patient safety and effectiveness outcomes in a longitudinal, observation data registry<br>• To implement a patient-centered, computer-enhanced learning model for optimizing treatment selection in precision oncology<br>• To improve access to precision medicine tests and therapies for patients with known or suspected recalcitrant*,advanced, or metastatic cancer<br>• |
| **Secondary Objectives:** | • To develop analytic methods and tools for patient group structure modeling using explainable artificial intelligence and machine learning<br>• To build an integrated system that combines machine learning with human experts from a Virtual Tumor Board (VTB) to generate customized patient treatment options<br>• To perform insight capture and analysis on the rationales underlying VTB recommendations and physician treatment decisions<br>• To develop methods that incorporate measures of patient goals and perspectives into cancer treatment option set development |

| | |
|---|---|
| | • To track patient outcomes over time and use analytic signal detection tools that combine machine learning and human expertise to identify meaningful safety and efficacy signals |
| **Exploratory Objectives:** | • To develop new outcomes measures, systems, tools and methods to advance patient-centered precision oncology |
| **Endpoints:** | The purpose of XCELSIOR is to develop methods, systems and algorithms for effectiveness and safety signal detection to improve treatment of cancer patients. Various endpoints will be tracked to identify signals of effectiveness or safety based on composite clinical, symptomatic, and other measures of patient disease burden and treatment response collected in routine clinical practice and directly from the patients. Examples include physician-reported Lansky, Karnofsky or ECOG performance status, patient-reported quality-of-life rating scales for the primary disease, such as the EORTC QLQ BN20 or MDASI-BT for brain tumors and the QLQ-PAN26 for pancreatic tumors, or general QOL questionnaires, such as the PROMIS survey instruments. Other patient-reported outcomes may be developed and used as part of the methods-development objective of this study, examples include patient statements and communications about their disease symptoms or functional status, photo and video submissions supporting assessment of treatment effects, or other patient submitted information may provide value for identifying treatment response or non-response and may be used as generalizable patient-reported outcomes that provide measures of cancer symptoms.<br><br>Analysis of effectiveness will be measured using responder analysis of composite clinical, functional, or symptomatic endpoints to accommodate the wide heterogeneity in disease burden and symptomatology in late-stage and recalcitrant cancer patients. As a patient-centric registry, patients themselves may propose, suggest, and/or submit evidence or ideas for functionally relevant outcomes in their disease for further development and implementation.<br><br>Additional endpoints for safety/tolerability and effectiveness will be derived from real-world data collected during routine care augmented as necessary with supplemental information from the patient or their physician with blinded reviews, as necessary, to relate information to measures collected in traditional clinical trials. Safety assessment may optionally include PRO-CTCAE as well as events identified or extracted from medical records. These may include, but are not limited to:<br><br>• CTCAE grade 3 or worse events<br>• Time-to-progression (TTP)<br>• Time-to-treatment failure (TTF)<br>• Complete Response (CR)<br>• Durable Response (DR)<br>• Response Rate (RR)<br>• Progression Free Survival (PFS) at relevant intervals<br>• Overall survival (OS) |

| | |
|---|---|
| | Endpoints may incorporate composite clinical, imaging, and laboratory markers developed as indicators of treatment response or failure. |
| **Study Population:** | This study will initially include up to 50,000 adult and pediatric patients with known or suspected recalcitrant, advanced, or metatstatic cancers. |
| **Key Inclusion Criteria:** | • Both male and female patients with known or suspected recalcitrant or advanced cancer are eligible to enroll on-line<br>• Patients with any performance status, comorbidity or disease severity are eligible<br>• Patients or their legally-authorized representative must be willing and able to provide written, informed consent (and assent, if applicable) |
| **Key Exclusion Criteria:** | • Patients must be a resident of or receiving care within the United States or US territories.<br><br>Note: Patients enrolling in XCELSIOR are not required to participate in the VTB process or access support Servicesfrom Cancer Commons or other partner organizations. |
| **Substudies:** | All patients that meet the XCELSIOR criteria and complete the sign-up process on-line will be enrolled into the registry. Those patients with specific sub-types of cancer, such as primary brain tumors or pancreatic adenocarcinoma, will be part of disease-specific substudies, which will include collection of measures specific to and appropriate for their disease. |
| **Phase:** | N/A |
| **Sites/Facilities Enrolling Participants:** | XCELSIOR is a patient-centric, real-world data registry. Patients may join the registry directly or through referral from partner or collaborator organizations, physicians, patient advocates over the Web using patient-provided and patient-authorized remote collection of medical information from their medical records. |
| **Description of Study Intervention:** | XCELSIOR is a non-interventional data registry. Information about treatments, treatment decisions and rationale, and patient outcomes including safety and effectiveness of anti-cancer therapy and associated supportive care will be collected for analysis. |
| **Study Duration:** | The longitudinal, observational study will run for an initial period of five years for patient recruitment with observation of enrolled patients indefinitely or until all enrolled patients are deceased and for a period of up to two years thereafter for additional data analysis and publication. The duration may be extended indefinitely based on the availability of funding. |
| **Participant Duration:** | Patients will be followed indefinitely until they are deceased |

*Recalcitrant cancers, as defined herein, are those cancers with a five-year survival rate of less than 50% or are likely incurable with existing therapeutic options

#### 1.2 Study Schema

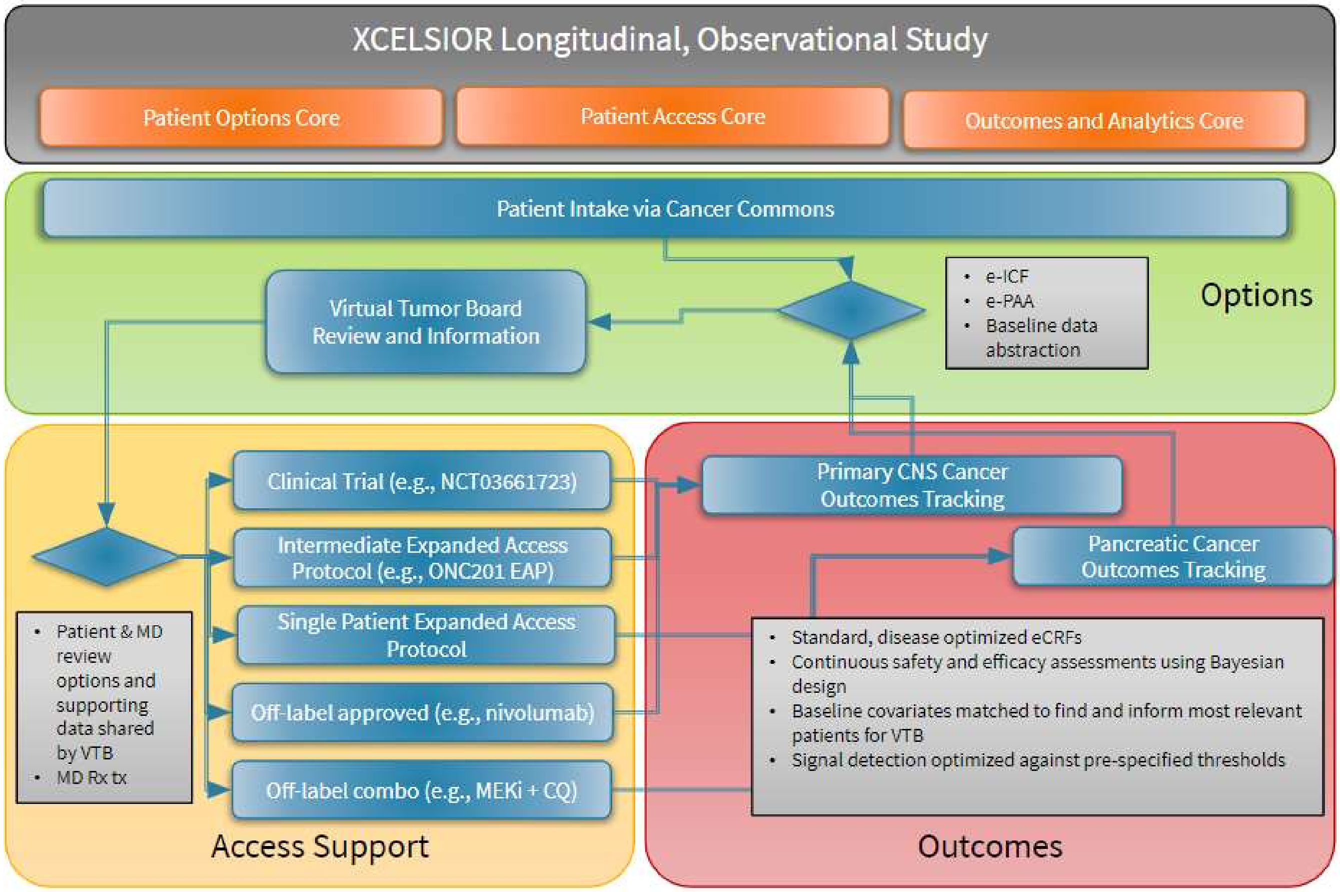

#### 2 STUDY RATIONALE

For late-stage and recalcitrant cancers, there are often no standard options, or the options are unsatisfactory with fewer than 20% of patients surviving more than five years from diagnosis. For many of these cancers, a clinical trial is the recommended, if not preferred, treatment option at some stage during therapy. However, according to the AACR, in 2015 only 5% of cancer patients (AACR, 2015) participated in a clinical trial. This number has barely increased in twenty-five years. For women and minorities, the percentage of patients that participate in clinical trials is even lower. The low rate of participation reflects certain barriers for these patients, including:

- Inclusion and exclusion criteria that limit the participation of some patients, and/or
- Geographic, financial, or logistical barriers to accessing centers that conduct clinical trials.

Patients and their oncologists also face challenges obtaining access to off-label, but medically logical therapies due to insurance or other financial barriers and to investigational drugs under expanded access programs due to unfamiliarity with medically-logical investigational options and/or the regulatory and associated institutional processes involved in acquiring drugs under expanded access.

In addition to low rates of participation, clinical trials, including newer master protocols, generally are designed to test a hypothesis regarding an investigational drug in a select group of patients. The definition of the patient population is typically defined by a deliberately narrow set of prior information, such as the tumor site of origin, histopathologic classification, number of prior therapies, and/or a single qualifying mutation. This approach provides strong mechanistic validity for specific therapies in specific tumors, but creates difficulty generalizing beyond very similar patients, with a goal of finding agents that are effective for a large percentage of the population, rather than to optimize therapy for an individual (Klement et al., 2016). In routine medical practice, drugs approved from traditional clinical trials are used to treat a more diverse group of patients than they were tested on during development.

The result of this situation is a gap in the external validity of well-controlled clinical trials from study subjects to all treated patients has been aptly described as “over-controlled empiricism” (Khozin, 2017). Thus, there remains a need to develop evidence for treatment effectiveness and safety beyond the information generated in trials designed to inform a device or drug label (Rothwell, 2005; Duan et al., 2013). In part, real world evidence (RWE) has been offered as a solution to complement traditional clinical trials. RWE is not limited just to naturalistic assessments of treatment effects in clinical practice or from medical records but can also include prospective registries that utilize validated and structured assessments (Sherman et al., 2016). XCELSIOR will use this approach as a foundation and incorporate the metadata about why decisions were made with AI to create a learning system for precision oncology.

Precision oncology seeks to determine which treatment is best for an individual patient to maximize their outcome. According to the National Library of Medicine, (NLM, 2018) this is approach is, “in contrast to a one-size-fits-all approach, in which disease treatment and prevention strategies are developed for the average person, with less consideration for the differences between individuals.” At present, much more information is required to make true precision oncology a reality, since most controlled clinical trials remain focused on treatment effects within a population. In a precision oncology registry, each patient can be considered uniquely with a treatment plan that incorporates a range of clinical and genetic information to make an individual-level hypothesis about the most appropriate treatment strategy for each specific person. This is the domain of normal clinical medicine. Each patient is considered and treated according to the best judgement of their physician, but within a registry this information can be aggregated into a system-wide learning model. This approach can help to break down data silos that exist throughout oncology research (Gyawali et al., 2017)

In oncology, there are now many targeted agents for specifically characterized mutations in the tumor genome. Use of targeted agents directed at specific, well-characterized oncogene mutations, often in concert with a companion diagnostic, has been validated as a treatment strategy through FDA approvals and improved clinical outcomes for patients. This mutation-diagnostic-therapy approach has been described as the first stage of precision oncology, or Precision Oncology 1.0. However, in cancer, the landscape for precision medicine is complex and requires knowledge about both patient and tumor. Adding to the complexity, tumors evolve over time and differently in response to different therapies. Into this complex landscape, there has been an explosion of information about patient and tumor genomes, exomes, transcriptomes, metabolomics and other information and how that information can alter disease risk, response, or trajectory. One approach has been the use of molecular tumor boards, which operate analogously to traditional tumor boards, but include a greater emphasis on, and more domain experts, including from pharmacology, systems biology, genetics, bioinformatics and other disciplines to support medical, surgical, and radiation oncologists, radiologists, and pathologists in recommending a course of treatment. Yet the pace of new information and time required for this approach makes it difficult to sustain or generalize widely. Thus, for precision oncology to advance, new approaches and tools will be required to move beyond mutation-therapy paradigms towards approaches that rapidly and continuously integrate information from clinical, “pan-omics”, pharmacology, and systems biology into specific recommendations for treatment of individual patients.

Clinical practice guidelines are helpful tools accelerating the process of integrating and evaluating the available evidence, which practicing clinicians adapt and customize for the individual patient’s situation. Yet evidence-based guidelines typically give the strongest weight to randomized controlled trials conducted to support marketing approvals and the meta-analysis thereof, which creates a potential for bias in the strength of recommendations due to the absence of randomized evidence from the sickest patients (Lim et al., 2008). Worse, these guidelines have often been used by insurers as a basis to deny coverage of potentially beneficial treatments selected by a clinician using molecular diagnostics as supporting evidence of medical necessity based on a targetable variant in the patient’s tumor (Sireci et al., 2017). Thus, in addition to the information gap necessary to implement precision oncology, there is an access gap to trials and treatments.

The goal and the promise of precision medicine is to define the best options for each specific patient. At present the available information, which includes patient and tumor genomics, transcriptomics, metabolomics, pathway analysis, and other advanced molecular techniques is outpacing the rate of discovery from clinical trials. Innovations in trial design are underway, including adaptive designs and master protocols (Woodcock and LaVange, 2017; FDA Guidance). However, these new approaches have not been without difficulty (Renfro & Sargent, 2016) and are designed to support regulatory approvals or proof-of-concept testing of drugs, rather than provide evidence on how best to treat individual patients. These studies, including NCI-MATCH, GBM AGILE, FOCUS4, LUNG-MAP, and I-SPY2, among others are an important step towards improving treatment options, but as noted by Van Allen and colleagues (2014), there is a very long tail of mutations detectable by exome sequencing in cancer patients. On average, that group found several hundred mutations in the 121 well-characterized oncogenes that they studied. This finding appears typical. Kurzrock and Giles (2015) described a similar situation in which patients were molecular snowflakes; each cancer patient’s tumor is unique and driven by a unique combination of molecular aberrations.

Today, information from next generation sequencing (NGS) and the number of approved targeted therapies already creates thousands of potential combinations of genetic target-monotherapy arms in a conventional clinical trial approach. A combinatorial view of this complexity (Figure 1, below) shows that even with the level of information available today, there are vastly more possible trial arms to study than there are cancer patients to fill them.

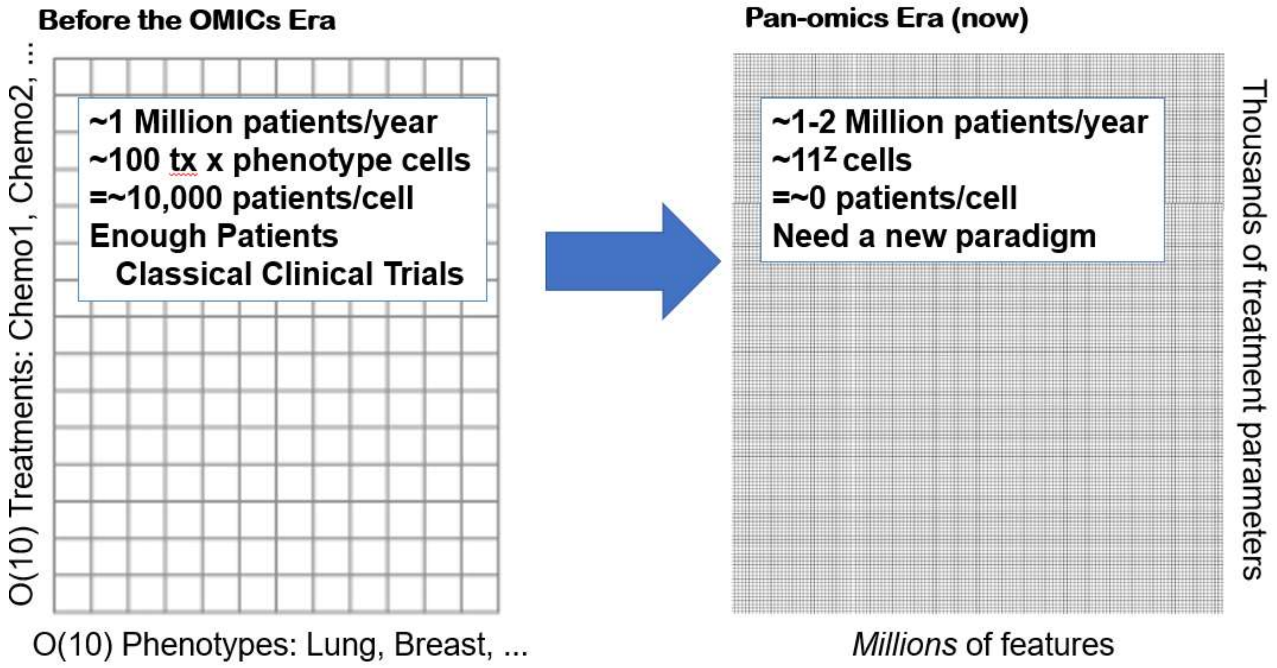

**Figure 1.** Schematic diagram of prognostic and treatment combinations courtesy of Jeff Schrager.

When considering this massive array of patient-treatment combinations, the scarcest resource in clinical research are the patients on which to test potential treatments. To move forward, many research groups have recommended the use of n-of-1 or aggregated n-of-1 style approaches to inform precision oncology (including Klement et al, 2017; Silvestris et al, 2017; and Markman et al, 2016; and Kurzrock and Giles, 2015). N-of-1 studies have their basis in early medicine and pharmacology, in which causality of drug effects is assessed with a challenge-dechallenge-rechallenge paradigm for establishing cause and effect based on an observed temporal relationship in which a patient is their own control. These types of “experiments” happen in the ordinary practice of medicine on a regular basis as physicians manage treatment response and side-effects. So, the future of precision medicine will be integrating the n-of-1 approach to treatment selection using panomic technology into clinical practice seamlessly and in a way that harnesses the data continuously (Schork, 2015).

At the opposite end of the spectrum from n-of-1 study designs are large registries and Real-World Evidence (RWE) derived from medical records. An example of a precision oncology registry is ASCO’s TAPUR registry (NCT02693535), which has been enrolling a wide spectrum of cancer patients by matching them to one of sixteen (16) arms of different targeted molecular therapies based on the most commonly detected pathway mutations for which FDA-approved therapies are available. Unfortunately, TAPUR excludes glioblastoma and other tumors that are not measurable by RECIST criteria. Similarly, NCI-MATCH, arguably the largest and most successful master protocol to date has collected more than 6,000 patient samples and matched about 18% of those patients into one of about forty (40) targeted therapy arms. More complex basket and umbrella trials have struggled during start-up and have difficulty allocating patients that match multiple targets. None of these trials is designed to answer the question of which treatment is best for a specific patient.

So, while these approaches are powerful, as Kurzrock and Giles noted, with the growing understanding of tumor genetics, monotherapy is unlikely to cure cancer. They noted that even for patients with a common aberration, the rest of their tumor is genomically distinct and thus customized, individual therapy, including combinations matched to the unique genetic aberrations that define each patient's tumor will be required. Furthermore, Klement and colleagues (2016) highlight the fact that in this situation a "vastly improved ability to establish the hierarchy of genomic alterations present," in terms of relative importance in order to make actionable treatment recommendations for individual patients. XCELSIOR aims to bridge the two approaches by creating a large patient-centered oncology registry with additional, disease-specific substudies in which individualized treatments that occur in the practice of medicine are captured prospectively in a registry on which analytic methods can help assess which combinations of factors predict favorable and unfavorable responses. This information can help to inform which treatments will work for individual patients and aid in the efficient search of the vast space of possible treatment combinations. This step is necessary to move towards the next iteration of precision oncology.

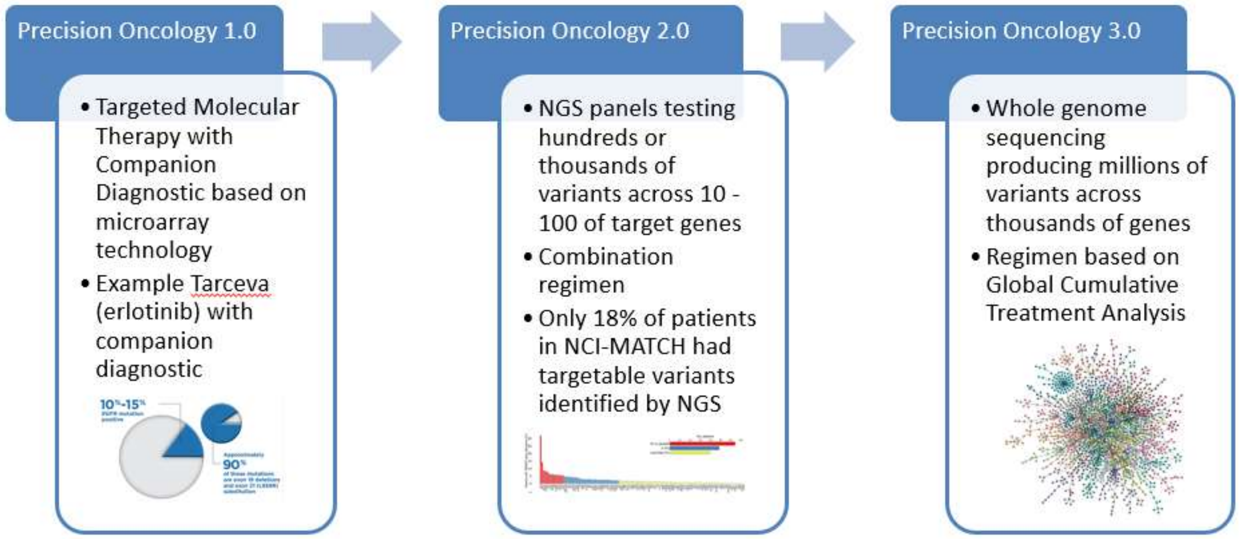

**Figure 2.** Evolution of Precision Oncology from 2000 to present.

There have been many legislative attempts to improve the development of and access to new therapies for cancer, and to amend the process of research and drug development to address issues raised by patients and their advocates. For cancer research, this began with public awareness and a declared "War on Cancer" in the 1970s. More recently, there have been several Federal laws that aim to address aspects of the challenges described above, including the:

- Childhood Cancer STAR (Survivorship, Treatment, Access, Research) Act (Public Law No: 115-180)
- FDA Reauthorization Act of 2017 Public Law No: 115-52 (H.R. 2430 / S. 934)
- 21st Century Cures Act, P.L. 114-255 (H.R. 6/H.R. 34)
- Early Act Reauthorization of 2014, P.L. 113-265 (H.R. 5185/S. 2655; 113th Congress)
- Gabriella Miller Kids First Research Act, P.L. 113-94 (H.R.2019, 113th Congress)
- Recalcitrant Cancer Research Act of 2012, P.L. 112-239 (S. Amdt. 3180 to S. 3254/H.R. 4310, 112th Congress)
- National Right-to-Try Act (S.204)

These laws reflect growing public concern with the availability of new and better therapy for cancer. Patient advocacy organizations have sought these changes and increasingly are working to enable their members to self-organize to promote or conduct research.

Cancer Commons is patient-driven nonprofit advocacy and support organization for cancer patients and their physicians. It is a community forum that is dedicated to continuous learning by bringing together patients, doctors and scientists to gather and share information that will help identify the best personalized treatment options for optimal patient outcomes. For many years, Cancer Commons has helped patients find and understand the most up-to-date research relevant to their disease and connect them with clinical trials or connect their physician with top experts for consultation and advice. Cancer Commons created xCures as a stand-alone company to further this vision through technology and as a vehicle to create a network or stakeholder relationships to extend the reach to more patient communities.

With XCELSIOR, the goal of Cancer Commons, xCures, and the Musella Foundation is to extend from just helping patients find and understand treatment options to a patient-centered system that can develop evidence for precision oncology. This study will be an “always-on” data registry or “perpetual protocol” to continuously collect and analyze longitudinal information in a way that closes the learning loop for patients, doctors, and tumor boards. This is the underlying data operations framework for XCELSIOR. Second, Cancer Commons will gather and analyze the metadata that is generated during decisions about patient care to continuously learn and incorporate new information that would partition and thereby narrow the search space for future treatment decision-making. XCELSIOR accomplishes this through the Virtual Tumor Board (VTB) process which integrates and scales human experts with AI and machine learning. Together the system is continuously learning and incorporating new information that narrows the evidence gap of what factors predict which treatments are best for an individual patient. This approach was described by Schrager and Tenenbaum (2014) as Global Cumulative Treatment Analysis (GCTA) and underlies the Precision Oncology 3.0 model.

In summary, XCELSIOR is a patient-centric approach to precision oncology that enables rapid global learning from every patient treatment. First, patients from anywhere can sign up for the Cancer Commons service, which is free and includes curation of their case by a cancer biologist and presentation to the VTB. Tools and techniques from artificial intelligence and machine learning help refine potential treatment options and analyze expert rationales, incorporate patient preferences for treatment, and present that information to the patient and their physician. Decisions about patient care remain with the patient and their doctor. Cancer Commons through XELSIOR provides a platform to track and analyze decisions, outcomes and the associated metadata. Information gain from routine care is then maximized according to theories and algorithms that are well developed in search engine optimization, but less common in medicine. In the XCELSIOR system, machine learning and artificial intelligence help integrate, assess, and inform clinicians and VTB experts while putting patients at the center of the process.

Precision medicine in oncology requires new approaches to generate information with the explicit goal of identifying the best treatment option(s) for each individual patient. This will require integration of AI and machine learning with human cancer experts to identify treatment options, ensure access to those options, and evaluate the outcomes of therapy for patients through collection of disease-specific data in a group of large, patient-focused registries. The goal of XCELSIOR is to pioneer the use of these approaches to generate actionable insights that will improve outcomes for late-stage cancer patients through better treatment selection, promote rapid learning about optimal treatments for specific subgroups of patients, and quickly share information to maximize the outcomes for cancer patients everywhere.

#### 2.1 Disease Setting and Trial Context

In the United States, there are more than 1.7 million cases of cancer and more than 609,000 cancer-related deaths per year. Despite progress in terms of lower incidence rates from major, preventable cancers, primary CNS tumors and pancreatic cancer have a dire prognosis and incidence rates have slightly increased in recent years (CDC/SEER, 2018). Cancer Commons helps patients with advanced or recalcitrant cancers across a wide range of tumor types. The initial focus of XCELSIOR will be building the infrastructure for patient intake/registration, VTB operations, access support services, and research tools and methods development to support the project aims. This will include a tumor-specific registry substudy initially for primary CNS tumors, followed later with a registry substudy focused on pancreatic cancer. Additional, tumor-specific substudies will follow, and now include pediatric cancers (leukemia, sarcoma, and brain tumors), ovarian and gynecologic cancer, colorectal cancer, bladder cancer, and cancers defined by molecular subtypes.

In the United States, more than 23,000 people per year are diagnosed with a primary brain tumor and according to the CDC/SEER Annual Report to the Nation (2018) death rates have slightly increased. Presentation of the disease is heterogeneous, requiring complex, interdisciplinary treatment and patient care. Outcomes are correspondingly diverse, but for glioblastoma multiforme (GBM), the most common intraparenchymal brain tumor, the prognosis is grim. This disease is virtually incurable and there have been several notable failures of large, randomized clinical trials in recent years (Bristol-Myers Squibb, 2017). Despite failing to meet the top line results, this trial had people with dramatic response (Dana-Farber Cancer Institute, 2017) indicating that the drug worked for an uncharacterized subset of patients, now thought to be those with high mutational burden. There are similarly positive results for nivolumab in single-case reports in pediatric GBM, despite the failure of the large, randomized trial (Alharbi et al., 2018). In contrast, Xhu and colleagues (2017) reported a life-threatening case of malignant cerebral edema in a pediatric GBM patient treated with nivolumab. In that case, causality was clearly established through a challenge-dechallenge-rechallenge paradigm.

Precision oncology requires research on how to identify those patients that will benefit or be harmed by a treatment, in order to maximize the chance that patients receive a beneficial therapy while avoiding treatments from which they are unlikely to benefit or may be harmful. A registry framework would enable these individual responses to be examined in aggregate to develop information that will guide precision medicine through the development of Bayesian hierarchical latent-class analytic models that can continuously learn and improve patient-grouping and subgrouping.

Although treatment guidelines encourage participation in clinical trials, in the absence of effective, approved therapies, many GBM patients are excluded from randomized clinical trials due to comorbid or other exclusionary conditions. In fact, one of the challenges to trial participation for late stage cancer patients can be clinical decisions made in the patient's best interest during earlier lines of therapy. Often neither clinicians nor patients are aware that such decisions can alter their eligibility for clinical trials later (Strauss 2013).

Patients and patient advocates have repeatedly sought more access to treatments and greater say in the research agenda. Most recently, the challenges experienced by late-stage cancer patients to access clinical trials or precision oncology therapy led to the passage of the 21$^{st}$ Century Cures Act (Public Law 114-255) in 2016 and S.204, the Right-to-Try law. More than forty (40) states also have some form of right-to-try law, further highlighting the demand from patients and advocates for better treatment options.

XCELSIOR will help give late-stage cancer patients agency in the care process and make their individual outcome a research priority. These patients have exhausted standard options, so the ability to access medically-logical treatments including clinical trials, off-label treatments with targeted therapies, novel combinations, or compassionate use of investigational drugs are the only chance these patients have. Cancer Commons will provide patients in XCELSIOR information by creating an equipoise set of therapeutic options determined by a VTB of disease experts. If patients and their doctors choose an option and require support accessing that option, the Patient Access Core at xCures will provide the necessary assistance on behalf of Cancer Commons, while the Data Core will track outcomes data through the XCELSIOR longitudinal, observation registry substudies. This is the overall approach for this project as shown in the Figure 3, below.

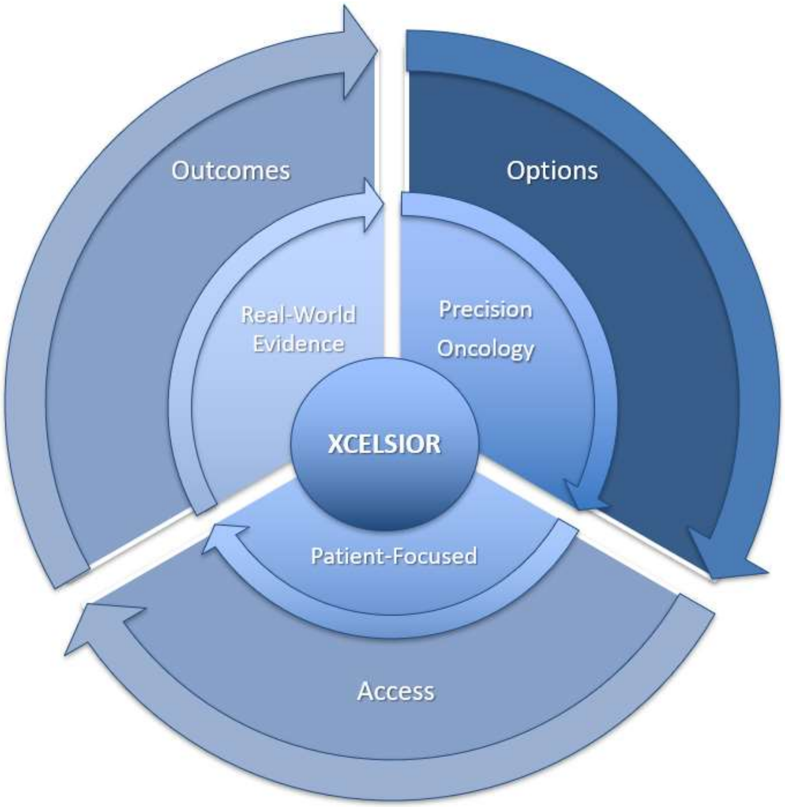

**Figure 3.** XCELSIOR model for implementing a patient-centered, enhanced learning model for cancer patients.

#### 2.2 Research Leading to the Proposed Study

##### 2.2.1 Cancer Commons, xCures, and the Musella Foundation

Cancer Commons is patient-driven nonprofit advocacy and support organization for cancer patients and their physicians. It is a community forum that is dedicated to continuous learning by bringing together patients, doctors, and scientists to gather and share information that will help identify the best personalized treatment options for individual patients. For many years, Cancer Commons has helped patients find and understand the most up-to-date research relevant to their disease and connect them with clinical trials or connect their physician with top experts for consultation and advice. More recently, Cancer Commons has been expanded this expert service to include case review and recommendations from a Virtual Tumor Board, which is a molecular tumor board of disease experts convened on-line using software.

xCures has been providing personnel, software, and services to support Cancer Commons and more recently both organizations have been working with the Musella Foundation, which had previously run a 20+ year patient-powered registry for brain tumor patients. Together the organizations have committed to helping bring patients and physicians together with other health system stakeholders, including researchers, testing labs, pharma and biotech companies, and insurers, to improve information about and access to treatments.

##### 2.2.2 Precision Medicine and the Role of Molecular Tumor Boards

The National Cancer Institute (NCI) defines precision medicine as, "medicine that uses information about a person's genes, proteins, and environment to prevent, diagnose, and treat disease. In cancer, precision medicine [also] uses specific information about a person's tumor to help diagnose, plan treatment, find out how well treatment is working, or make a prognosis." This is a major focus of the 21st Century Cures Act (Public Law 14-255). Drs. Collins & Varmus, the Directors of the NCI and NIH discuss the role of precision medicine (2015) as a," more individualized, molecular approach to cancer will enrich and modify, but not replace, the successful staples of oncology — prevention, diagnostics, some screening methods, and effective treatments — while providing a strong framework for accelerating the adoption of precision medicine in other spheres."

As the number and types of available tests has increased (see Figure 4, below), precision oncology has been undergoing an evolution with new tests that provide more information. While not ubiquitous, NGS panels for specific and general tumor types are becoming widespread. Precision oncology requires note only the use of such tests, but the ability to incorporate new information on an ongoing and continuous basis.

| Molecular Testing Method | Variant Type | | | |
|---|---|---|---|---|
| | Single nucleotide variants (SNVs) | Small duplications, insertions, indels | Exon duplications, deletions, or gene copy number changes | Structural Variant (SVs) |
| Allele-specific polymerase chain reaction (PCR) | ✓ | | | |
| PCR & Sanger dideoxy sequencing | ✓ | ✓ | | Detected if fusion RNA extracted |
| PCR & Mass spectrometry (MS) | ✓ | | | |
| PCR & single base extension | ✓ | | | |
| Multiplex ligation-dependent probe | ✓ | | ✓ | |
| Fluorescence in situ hybridization (FISH) | ✓ | | Only gene copy number variants | ✓ |
| NGS – custom panel (amplicon capture) | ✓ | ✓ | | |
| NGS – custom panels (hybridization capture) | ✓ | ✓ | ✓ | Known breakpoints |
| NGS – whole exome sequencing | ✓ | ✓ | ✓ | |
| NGS – whole genome sequencing | ✓ | ✓ | ✓ | ✓ |
| ddPCR | ✓ | ✓ | | Known breakpoints |
| BEAMing | ✓ | ✓ | | Known breakpoints |

**Figure 4**. Adapted from: Vnencak-Jones, C., M. Berger, W. Pao. 2016. Types of Molecular Tumor Testing. My Cancer Genome https://www.mycancergenome.org/content/molecular-medicine/types-of-molecular-tumor-testing/ (Updated February 8, 2018).

With many tests available and new tests, technologies and tumor markers being developed almost daily, there are significant challenges for practicing clinicians and their patients to stay up-to-date and gain access to relevant tests. Further, any research effort must be flexible enough to accommodate new and changing testing information. As this information has increased, tumors have become more narrowly defined and it is increasingly clear that even for two patients with a tumor from the same organ with the same primary mutation, the rest of the tumor genome and transcriptome will vary widely (Kurzrock and Giles, 2015).

Further complicating this picture is that the tumor genome will change over time (Wang et al., 2016b; Klement et al., 2016). As more information is developed, the number of organ x mutation x drug combinations rapidly grows to the point where, when taken to a logical conclusion, most possible arms or cells of an experiment have an "n" of zero or at most one patient. Instead of a traditional approach, a patient-centered approach for precision-oncology would as ask not which drugs work in a group of patients, but rather which drug will work for a specific patient. This approach requires the integration of large amounts of data and continuous learning so that precision medicine trials evolve towards detailed "panomic" information (including but not limited to genomic, transcriptomic, metabolomic, or other "omic" analyses of tumors), combined with every available piece of clinical, pathology, or other diagnostic data together with each treatment and outcome as an interrogation of the tumor that can be learned from and inform future decisions. However, despite the growth in possible information it is uncommon for organizations to conduct tumor boards for late stage cancer patients.

Examples of this approach include the molecular tumor board (MTB) approach used at UC, San Diego (Parker et al, 2015) which showed that molecular approaches to treatment led to a 30% increase in PFS relative to the prior line of therapy in 41% of cases considered by the MTB. Similarly, Radovich and colleagues (2016), reported that 43% of patients treated with a genomically targeted therapy achieved a 1.3 or greater PFS relative to their prior line of chemotherapy, versus only 5% of patients that received non-genomically targeted therapy. Such findings have led Markman and colleagues to suggest the use molecular tumor boards to devise targeted treatments for patients using an n-of-1 approach, with prospectively defined endpoints as a measure of efficacy. Specifically, they suggest a 1.3-fold ratio of time-to-subsequent-disease-progression as a marker for biological and clinical utility in precision oncology. Further, they implemented this approach with their MTB and present fifteen cases in which crizotinib was suggested for patients with ALK gene mutations. In that cohort 55% of patients had a PFS on crizotinib that was at least 1.3-fold longer than on their prior line of therapy. In fact, while NCI-MATCH only had about 18% of patient samples had actionable targets identified, MD Anderson's IMPACT study, which used genomic profiling to help assign patients to active Phase 1 clinical trials (and thus had access to a uniquely large pool of targeted therapies, showed nearly 50% of patients could be assigned to a clinical trial therapy based on an actionable mutation and that those patients were twice as likely to have a clinical response and experience a statistically-significantly longer median OS (Tsimberidou et al, 2017). Sireci and colleagues (2017) at Columbia University Medical Center using the Columbia Combined Cancer Panel assay detected a targetable variant in 48.7% of cases, however, they found that reimbursement from government and third-party payers using was just 19.4% of billed costs, and 55% of cases were rejected on first submission. What these experiences show is that a molecular tumor board approach with access to many targeted therapies, including investigational and off-label treatment options, is likely to improve the treatment outcome for a large fraction of cancer patients, but that work remains to ensure treatments from this approach are available to patients.

To match the best treatment for each individual patient based on what is known at the time, a patient-centric approach to clinical research is required. This allows the patient and their physician to course correct as new information becomes available in a seamless manner. The rationale for changes in response to new information fits nicely with Bayesian models and in a patient-centered research effort, the ability to learn positive and negative information and quickly share that with other patients and doctors should help improve the outcomes for all participants. The goal of this approach is not to provide evidence about which drugs work on average, but rather to generate evidence about which patient- and tumor-specific factors are associated with response to treatment, such that other patients may benefit from that information. XCELSIOR will perform this using a Virtual Tumor Board (VTB) process, which is an interdisciplinary and molecular tumor board involving experts convened using remote technology augmented with AI and machine learning analysis of other patient data. Cancer Commons and xCures have successfully piloted this process for several GBM and pancreatic cancer patients.

More recently, new types of precision testing have emerged, including circulating tumor NGS tests based on cell-free DNA, circulating tumor cells (DNA and RNA), circulating tumor and non-tumor exosomes (protein, DNA, and RNA). In addition, drug sensitivity testing, which aims to examine specific treats for an individual by screening anti-cancer compounds against tumor cells, for example in the form of organoids (microscopic tissue grown directly from patient tumor cells). These approaches add to the nature and amount of information available for consideration and can be helpful in cases where a tumor appears “cold” under standard NGS.

The goal of the VTBs to design a set of best options for each specific patient, considering not just tumor location, histopathology, or a molecular marker, but rather take a holistic approach that integrates all available information with a large database of patients with treatments and clinical outcomes. In that sense, each patient is an n-of-1 and forms their own arm within a large registry. The VTB receives case information and options prepared by a cancer biologist with support from the xCures AI system which is informed by the XCELSIOR dataset and employs state-of-the-art analytic techniques to identify similar patients and use information about their treatments, treatment-decision process, and safety or effectiveness outcomes to inform treatment recommendation. After that, the assigned VTB members review the case and available information, apply their own expert knowledge about pathways or mechanisms, and then prepare a limited set of options for treatments, or recommend additional testing. This information is provided to patients and their oncologists as a set of options for consideration with supporting evidence only. The patient and their doctor ultimately make a treatment decision, as they do already in normal clinical care. The VTB will gather information about what treatment they choose and why, and then incorporate that information into the analytic model and utilize it to inform treatment options for future patients. This iterative process is important to develop personalized medicine information in a systematic way that puts the individual patient outcome at the fore.

##### 2.2.3 Patient-Centric and “Siteless” Clinical Trials and Registries

Patients and patient-advocacy organizations have increasingly sought to drive the research agenda related to their diseases. This includes development of assessment methods, creation of research infrastructure and increasingly patient-centered or patient-led research programs. Examples include PanCAN’s Know Your Tumor program, and numerous “Patient-Powered Research Networks” established by the Patient-Centered Outcomes Research Institute (PCORI) as authorized by the Affordable Care Act. This program includes a myriad of patient registries, including Improve Care Now: A Learning System for Children with Crohn’s Disease and Ulcerative Colitis, the NIMH Child & Adolescent Psychiatry Trials Network (CAPTN), and the Multiple Sclerosis Patient Powered Research Network (PPRN). The Cancer Commons and xCures team includes veterans of several of these initiatives.

Patient-centered registries and patient-led studies are thought to be more generalizable than randomized clinical trials because they can capture information about a sizeable percentage of the entire population of interest. The internet and electronic medical records (EMR) have proven to be enabling technologies, but access to technology can create a theoretical bias in these samples, making them complementary to traditional RCT designs. (Gliklich et al., 2018).

There are many benefits to this approach, as enumerated in the principles of patient-centered research outlined by the National Organization of Rare Diseases (NORD). They specify that patient-centered research should involve patient-focused methodologies, collect interoperable and externally-relevant data including genomic and historic patient data, ensure that patient organizations own the data generated from their patients, and include QOL measures in addition to clinical data or endpoints. Furthermore, they encourage research designs that target all patients and help facilitate access to treatment. XCELSIOR is designed to adhere to these principles in the furtherance of treatment and research for the benefit of cancer patients.

This approach has increasingly been of interest to those conducting clinical trials. In 2011, Pfizer initiated the REMOTE (Research on Electronic Monitoring of Overactive Bladder Treatment Experience) study, a trial designed to compare whether a study conducted via the internet could replicate the results of the same study design implemented in a traditional, site-based manner. This approach was explored due to the challenges of drug development, including time, cost, and complexity. More recently, many organizations have sought to move clinical trials from a doctor and site-centered model to a remote model using, for example telemedicine to assess and interact with patients (Hirsch et al., 2017). This approach has been thought to be feasible for simple diseases treated in an outpatient setting but unlikely to succeed in oncology which involves complex diagnosis and treatment (B. Hirsch, personal communication).

However, learning from both approaches, Cancer Commons believes that XCELSIOR can create a platform with a patient-led, distributed model for research in oncology. Specifically, patients register and using their right-of-access enable collection of data about their treatments. The platform will enable consideration of a wide array of treatment options and exchange between patients, their oncologists, and disease experts in order to optimize treatment options and track outcomes over time. This allows patients to band together to facilitate the study of treatments and place their outcomes into a wider context that can help other patients. For patients in XCELSIOR, who have no acceptable standard options, their treatment can be optimized based on diagnostic information in relation to a large sample of other patient experiences and becomes a series of n-of-1 experiments to find their best treatment and generalize that information.

##### 2.2.4 Expansion of Treatment Options

For several years, Cancer Commons and xCures have helped cancer patients that have exhausted or can't tolerate conventional treatment options. This service began with a second review and clinical trial matching service and evolved into a virtual tumor board, in which experts voluntarily discuss the case and share ideas for treatment options to consider in discussion with their oncologist. Moving forward, Cancer Commons will expand and formalize this service and gather data on outcomes through the implementation of the XCELSIOR protocol. This will help to better inform future patients coming to Cancer Commons, but also to publish systemic information gleaned from these patients' experiences in order to more rapidly advance improvements in cancer care.

It has been reported that about 80% of clinical trials are behind on patient enrollment. (Glass et al., 2016). By putting the patient at the center of the trial, both constructing the treatment arm specifically for the patient, and bringing the treatment to the patient and doctor in normal clinical practice, several major barriers can be eliminated. Yet some barriers to this approach remain. In the experience of Cancer Commons, many patients pay substantial out-of-pocket costs to travel long distances to get into cancer clinical trials that are not available locally. Several foundations exist to support patient out-of-pocket costs, which can be prohibitive even when a clinical trial covers most medical costs. In addition, Cancer Commons patients have several times switched doctors to get treatment at a site that hosted a clinical trial in which they wished to participate, only to find that the busy principal investigator never approached them about the trial or the trial was already closed to enrollment.

Following the provision of options to patients and their oncologists, the second function of this study is to facilitate access to treatments. In many cases, patients face difficulty access the full range of treatment options due to a variety of barriers. These barriers can exist at comprehensive cancer centers but are more prominent in community settings across the United States. The goal of the Patient Access Core is to facilitate access to the options selected by patients and their oncologists, whether they choose an option presented by the VTB or another option. What Cancer Commons has found anecdotally, is that many patients and/or their oncologists fail to consider or discuss all options because of perceived barriers. Those barriers include cost, insurance coverage challenges, and difficulty among community oncology practices to navigate the expanded access process. The challenges that patients experience at this stage cannot be over-emphasized. Not only is the process for compassionate use slow and difficult for patients and doctors to navigate, but in late-stage cancer, days or weeks matter, and this process rarely happens in less than three weeks, but often takes longer. In fact, while FDA and IRBs have become quite responsive to these requests, often administrative barriers within hospitals related to legal and billing questions can take up weeks. Worse, for patients that go through the hoops to get access, if they do not respond, the whole process begins anew with another sponsor. Therefore, we intend to build a portfolio of compassionate use programs through outsourcing relationships with sponsors. The goal is to build a clearing house where patients and their doctors can come to a single source for facilitating compassionate use for patients from who it is a recommended option by the VTB or their physician. Cancer Commons and xCures will then provide a central support mechanism to assist with access. Such activities, which are governed by 21 CFR 312 are subject to additional procedures and will operate under protocols outside of the XCELSIOR registry with additional safeguards under 21 CFR 50, 54, and 56, although patient data for those protocols will be collected under their participation within XCELSIOR.

##### 2.2.5 Expansion of Testing Options

Cancer Commons, as part of their services to patients, may help patients that may need additional sample testing to support VTB recommendations to access appropriate testing as requested by the VTB. Thus, Cancer Commons may help facilitate transfer of patient samples either from the patient or their healthcare provider for additional molecular, genetic, and histopathological examination to evaluate or identify biomarkers, or test biomarker assays in support of their treatment and development of precision medicine information more generally. Recommendations and support for testing will be on a voluntary basis and the decision to participate or not will not affect participation in the XCELSIOR. Data generated from laboratory testing will be treated as Services Data, but the results of such data may be abstracted into the Research Data. xCures alone or on behalf of a partner or collaborator may facilitate transfer of patient samples at the patient's direction to a laboratory or repository for storage to use in future research studies in de-identified form. Reports from CLIA-certified laboratories may be provided directly to the patient. Other reports generated from deidentified samples under experimental conditions would be processed and tracked only with the patient's unique study ID number and the date that the sample was obtained. This link between the patient's study ID number and their identifiable information will be available only for decode internally by xCures for purpose of providing Services, such as facilitating review by the VTB and the results of such tests may be made available to the patient's physician. Further information can be found in Section 6.1.

##### 2.2.6 Artificial Intelligence and Advanced Analytic Methods

Cluster analysis is the mathematical approach to grouping things that are alike or partitioning data into similar groups and sub-groups. Clustering models using artificial intelligence (AI) and machine learning (ML) algorithms can provide a systematic and statistically robust model of empirical patient classification. Although ML and AI are based on application of advanced, but well-studied statistical methods, what makes them different from traditional applications is the iterative nature of the algorithm when implemented in a computer program. This allows the system to optimize through use of training data and repeated application of the algorithm to optimize the weighting of coefficients. This is closely related to the use of sampling and simulation using Monte Carlo methods to assess and improve model performance. Such approaches have become essential to handle the vast amount of data now being generated in many fields. These approaches have also found powerful applications in areas where the number of parameters greatly exceeds the number of sample items, a situation which is common in Big Data applications has now become common in medicine, but medicine has been slower to adopt these approaches that other areas. In terms of classification problems, ML approaches contrast with traditional approaches to patient classification in a clinical trial in which the population is pre-specified by a set of inclusion and exclusion criteria, such as age, gender, disease stage, prior therapies, and perhaps molecular status. Such an approach is typically "by committee" with input from the clinical development team, investigators, and regulators; however, it is rarely the subject of empirical investigation after the fact. Instead, what precision medicine requires elucidation of the covariance matrix that defines the strength of knowledge about the relationships of those criteria that define the group and the ability to refine the model with even new patient experience.

The emergence of new AI and ML algorithms and computing power to use those algorithms on data has grown dramatically in the past several years. These algorithms include:

- Bayesian models
- Neural networks and deep neural networks
- Decision Trees and Random Forest Ensemble models
- K Nearest Neighbor models
- Support Vector Machine models

A common feature of these approaches is the ability to group and cluster data in order to identify very subtle patterns and to learn and continuously update the model based on new information, while at the same time making theoretically sound statistical inferences. While some of the approaches require training data, others can operate without historic training data. Either way, methods exist to incorporate new information with historic information to continuously improve the model. The theoretical mathematics underlying these approaches has grown substantially in the last 20 years, and the use of statistical inference for A/B testing has exploded with the use of those models for internet-based commerce. Because of the speed of innovation, there are many applications for these advanced testing methods using machine learning in healthcare. Many of these approaches, while based on valid statistical inference, rely on big data without underlying assumptions about the distribution or underlying structure of the data (i.e., non-parametric methods). Together with Markov Chain Monte Carlo simulation with these approaches creates the possibility of measuring prospectively, the potential value of information gained from different treatment options at any given time, making for a nimble experimental model. At the same time, there are limits to how precisely our knowledge about patients can be applied to individual cases, due to the complexity described in Figure 1, meaning that empiric clusters are probably the limit of inference possible in precision medicine. This is reviewed by Ioannidis (2009).

All these approaches to cluster analysis allow for the iterative update and improvement of group definitions using empirical approaches. Modern methods of clustering allow for learning, identification of hidden relationships, and require fewer assumptions about the underlying structure of the data. For example, Bayesian Non-parametric approaches to clustering do not require assumptions about the number of clusters, shape of the data, or number of dimensions. Such an approach also allows updates to the confidence of the groupings based on each new observation. There have been several other relevant innovations, including advances in nearest neighbor searching algorithms, which can also be used for clustering, as well as learning neural networks, and other techniques of machine learning that can inform the grouping of patients to improve the identification of similar patients based on all available information rather than a small group of inclusion and exclusion criteria. These approaches have increasingly been used to facilitate diagnosis and prognosis for patients, which has been reviewed recently by Jiang and colleagues (2017).

It is well established that things like the location of tumor and age of onset correlate strongly with molecular features in some tumor types. Machine learning is ideal for understanding the hidden relationships between these types of data in order to build a model of patient groups. In fact, the proposed approach of Bayesian NP clustering also provides a mechanism to fit patients to clusters without needing the same or even a complete set of data for all patients. This technique allows the mapping of a new and different dataset on to clusters produced from another dataset. For example, using registry data from xCures partner, the Musella Foundation for Brain Tumor Research and Information, Inc, a cluster model can be produced and new registry data from patients collected under this protocol can be fitted to existing groupings, while also informing and updating the group structure.

##### 2.2.7 Incorporating Patient Goals and Values in Treatment Choice

Another important aim of XCELSIOR is to develop quantitative methods to better incorporate patient preferences into the treatment decision-making process. This is important to ensure that patients values and preferences are matched with possible treatment options.

As Kahneman and Thaler explain in their 2006 review (Kahneman and Thaler, 2006), the concept of utility as introduced by Jeremy Bentham more than 100 years ago is concerned with happiness “as the temporal integral of momentary experienced utility.” However, the meaning has evolved in psychology and economics research to refer to utility as an aspect of decision utility. They characterize the distinction as follows: “In the older interpretation of utility, the question of whether choices maximize utility has a simple meaning: do people choose the options that they will most enjoy? In modern decision theory, which ignores the distinction, the question is quite different: are preferences consistent with each other and with the axioms of rational choice?”

In modern decision science, individual utility can be described as a mathematical function constructed based on paired choice experiments. In other words, by making complex trade-offs, consumers or patients’ preferences can be revealed through mathematical (regression) analysis. Mohamed and colleagues (Mohamed et al., 2006) used this approach for patients with renal cell carcinoma to quantify patients’ benefit-risk preferences regarding benefits, toxicities, and serious adverse events associated with different treatments for advanced renal cell carcinoma. Perhaps unsurprisingly, this analysis revealed that the prospect of a seven-month increase in progression free survival (PFS) far outweighed severe chronic fatigue, severe gastrointestinal problems, or a 2% risk of very serious and potentially fatal adverse events. In fact, for patients with a baseline expectation of only three-to-four months of PFS, for every one-month increase in PFS, patients would accept a 1% increase each in the risk of potentially fatal adverse events, lung and liver damage.

When considering the incorporation of AI or machine learning algorithms in medicine, it is important to recognize that algorithms reflect the choices of the builder. Specifically, learning algorithms can incorporate bias through the selection of a training data set and definition of “success” built into the learning algorithm.

For that reason, when building a system to integrate expertise with machine learning, it is very important that a quantitative measure the patient’s goals and values is built into process. Although incorporation of quantitative measures is not yet widely used enabling technology exists and there are several arguments for increasing use of this approach, which is generally called shared decision making (SDM). SDM is an approach to clinical decisions in which patients and clinicians work together to reach a mutually agreed on decision that is consistent with the best available evidence, as well as patients’ preferences. This frame is designed to assist in cases where there is equipoise between options, as is the vase for late-stage and recalcitrant cancers, where available options are all poor, and investigational options are at equipoise due to overlap in variance. In these situations, Clinicians’ recommendations can differ based on their interpretation of uncertain data, which can pose challenges for patients trying to make sense of the information. Thus, patients’ preferences for the possible risks, benefits, and trade-offs between options can help guide the process.

##### 2.2.8 Real World Evidence

Real-world evidence (RWE) is defined as data regarding the usage, the potential benefits or risks, of treatments from sources other than randomized clinical trials, including disease registries. Use of such evidence has the potential to allow researchers to answer questions about treatment effects and outcomes efficiently, saving time and money while yielding answers relevant to broader populations of patients than would be possible in a specialized research environment, such as a randomized clinical trial (Sherman et al, 2016).

Real-world evidence has been increasingly valuable in oncology across several dimensions. First, through the collection and analysis of medical records information, RWE has allowed for the creation of "synthetic control arms" for randomized trials. This approach, in which co-variate matched patients are drawn randomly from a large population to serve as matched controls for patients in a clinical trial provides several benefits (Abernathy et al., 2017). First, it reduces the exposure of patients to placebo in large clinical trials and second, it reduces the cost of clinical trials, encouraging more investigation of active compounds.

The 21st Century Cures Act directed FDA to develop formal mechanisms to make greater use of RWE in support of drug labeling. Several companies have found creative ways to use RWE to support their marketing approval applications. For example, Ultragenyx has used RWE collected on patients in their Expanded Access programs to augment regulatory submissions in rare disease (E Kakkis, personal communication). Similarly, Novartis has used RWE collected in expanded access programs to support approval of development of ruxotilinib in myelofibrosis and ceritinib in lung cancer (P Alio, personal communication).

##### 2.2.9 Aggregating N-of-1 Experiments and associated Patient Outcomes

In his 2006 review of Bayesian clinical trial designs, Dr. Don Berry discussed the appropriateness of this approach for cancer, especially in the context where prior information about drug's effectiveness exists for other types of tumors or based on molecular markers. Strong prior information invites a Bayesian model for assessing consecutive instances of treatment and response. Many others have called for aggregating n-of-1 experiments in oncology as the next step towards personalized medicine in this field. For example, Lillie and colleagues (2011) in the journal Personalized Medicine, review the historical basis for n-of-1 studies in clinical and behavioral medicine and note that while the challenge of individual n-of-1 experiments is generalizability, they fit squarely with the clinical goal of optimizing individual patient outcomes. While traditional n-of-1 studies have involved both blinding and randomization of the treatment sequence (A then B or B then A, denoted AB or BA, for example) this approach is less practical in oncology. However, the advances described by Lillie and others in developing statistical methods to aggregate and analyze individual patient treatment experiences can be applied more generally. For example, Zucker and colleagues (2010) demonstrated the use of Bayesian hierarchical methods in relation to other meta-analysis approaches, including mixed-effects regression models to combine N-of-1 trials and assess the impact of outliers, missing-data, and parametric versus non-parametric assumptions of variance to build robust estimates of treatment effects and compare those results to data from a large, multi-center RCT. Weinreich and colleagues (2017) used a similar approach that supported reimbursement approval in Europe for an off-label use of an approved drug in a rare disease population.

Randomization and blinding, while helpful for certain aspects of clinical trial validity in n-of-1 designs are not required for the aggregation or meta-analysis of data and statistical inference thereof, nor are they essential for making inferences about the effectiveness of cancer treatments. As noted in the FDA Guidance on Clinical Trial Endpoints for the Approval of Cancer Treatments, Objective Response Rate and Complete Response can both serve as approvable endpoints in single-arm, open-label cancer studies. Within the XCELSIOR study framework, use of internal controls, such as prespecified estimates for patient trajectory based on individual characteristics, for example TTP ratio, as investigated by Mick and colleagues using paired TTP analyzed as a hazard ratio (Mick et al., 2016), enable a prospective evaluation of effectiveness with each patient as their own control, the result of which can be aggregated using Bayesian or other meta-analytic methods. This approach is especially powerful in rare diseases (Abrahamyan et al., 2014), which is a useful model for precision oncology, in which NGS data with other baseline covariates places each patient into at most, a very small cluster of similar patients. Those authors further highlight the importance of looking for large effect sizes in rare diseases, which is a key aspect of signal detection in precision medicine generally and XCELSIOR specifically.

###### 2.2.10 Assessment of Study Feasibility

In the last several years, Cancer Commons, xCures, and the Musella Foundation have helped thousands of late-stage cancer patients. This has taken the form of a review of their complete medical case by a cancer biologist, recommendations for clinical trial options or additional testing that would inform potential treatment options, and in some cases helping with support for patient access to trials, and second opinions experts in specific molecular pathways or tumor types. This protocol will extend this work into a systemic framework for knowledge capture and analysis to advance and improve treatment options, access, and outcomes for late-stage cancer patients.

#### 2.3 Specific Problem Statement

The problem of internet search bears some relation to the How to turn every patient treatment into an opportunity for system-wide learning, then lead into the barriers to implementing that.

##### 2.3.1 Options and Access

For late-stage and recalcitrant cancer patients, there are often no standard options. Every year, many thousands die prematurely because their doctors do not know the optimal way to utilize currently available tests and therapies. Patients and physicians also face formidable administrative, regulatory and financial challenges obtaining the latest therapies off-label or through clinical trials and expanded access.

Developing a new cancer drug takes about a decade and a billion dollars, much of which is expended in clinical trials. Trial accrual is challenging because, as the New York Times observed, “There are too many experimental cancer drugs in too many clinical trials, and not enough patients to test them on.” Moreover, the most effective treatment protocols involve intelligently designed, individually tailored, sequences and combinations of tests and drugs, and there are exponentially more plausible regimens than can be accommodated in the current clinical trial paradigm.

Already the large research efforts in precision oncology have started to experience challenges, including “unexplained drug resistance, genomic heterogeneity of tumors, insufficient means for monitoring responses and tumor recurrence, and limited knowledge about the use of drug combinations.” (Collins & Varmus)

##### 2.3.2 Outcomes and Analytics

There are several challenges to developing precision oncology information with traditional clinical trial approaches that can be overcome in a patient-centric registry study. As noted, most clinical trials based on frequentist designs aim to test a hypothesis about a treatment, rather than about a patient. This means that for patients that do not respond to treatment, the deduction is that the treatment didn’t work for that patient. For example, in a randomized clinical trial group definition may not include all available information. Self-imposed limitations in the knowledge used to characterize the study groups can bias or even invalidate the conclusions. This issue, involving hidden variables, has been identified as an underlying cause for the reproducibility issue in clinical research (Ioannidis, 2005). In other cases, new information emerges during the study that impacts the interpretability based on how the population was defined or the outcomes were measured. Another challenge with traditional approaches occurs with rare cancers, which is the fundamental challenge of precision oncology. Already, FDA Guidance for Developing Targeted Therapies in Low-Frequency Molecular Subsets of a Disease recognizes that traditional approaches will not suffice in such settings. However, the approach outlined requires new analytic methods from combinations of in silico modeling and simulation with supportive evidence for the underlying hypothesis. Similarly, designs for rare tumors may be single arm and rely on response rate-based endpoints. Guidance supporting this approach for non-randomized designs in oncology is available. However, it remains challenging to find these patients. There may be few clinicians that specialize in treating patients with rare cancers, and to conduct a clinical trial, a large number of sites is often required. Base of the time required to recruit patients into these trials the reporting of result often takes years. During delays or long periods before trial reporting, other patients may be exposed to substandard treatments or not get access to effective treatments. New approaches are needed for rapidly learning and continuous dissemination of clinically relevant information.

#### 2.4 Goals and Objectives of XCELSIOR

XCures alone or on behald of a partner or collaborator through the XCELSIOR registry will facilitate the development of actionable insights for precision oncology. The XCELSIOR system will allow patients to drive the evaluation of treatments with the goal of optimizing the selection of effective treatments for late stage cancer patients. This will be accomplished by expert review in the form of a virtual tumor board (VTB) aided by machine learning (Patient Options Core) paired with a patient-centered RWE data registry (XCELSIOR) that integrates structured and unstructured longitudinal data (Patient Outcomes Core). Bridging these is the services component of Cancer Commons and xCures, the Patient Access Core, which exists to support access to treatment options for patients, a necessary step to ensuring the evaluation of real-world treatments for safety and effectiveness is not inhibited by biases or limitations in access to therapies. Together, this approach, Options, Access, and Outcomes, enables a continuous learning model on which Global Cumulative Treatment Analysis (GCTA) can take place and advance the state of precision medicine in oncology through iterative, systematic investigation and advanced analytic methods.

The XCELSIOR framework involves three key areas that will generate data that will inform future treatment decisions in a continuously-learning framework modeled on search methodology. This approach complements traditional clinical trials by trying to glean knowledge from every patient treatment experience continuously in a manner that can maximize information gain. To do so, this protocol will build a patient-focused framework for oncology drug development.

This protocol will facilitate an expanded list of patient-specific treatment options for consideration by their oncologist that is curated by experts, informed by machine-learning, and customized for individual patients; Second, the protocol will facilitate access for the patient and their oncologist to their treatment-of-choice by providing support for treatments utilizing a combination of foundation funds to support access to treatment or care; negotiation directly with sponsors and insurers on behalf of the patient for coverage, when applicable; creating a centralized clearinghouse to support oncologists with expanded access for compassionate use. Finally, this protocol will provide a mechanism to gather this data into a longitudinal, operational study (registry) framework that layers disease-specific outcome measures over medical record data and combines that with innovative signal detection algorithms into a Bayesian hierarchical framework for safety or effectiveness signal detection to continuously update information facilitate global learning.

##### 2.4.1 Patient Options Core

Experts on the VTB review the case and develop an equipoise set of options that have the strongest rationale for success. The VTB platform created by Cancer Commons will use analytic techniques to provide additional information to the patient and their physician about the known risks and effectiveness of each treatment in the equipoise set. Specifically, the patient's physicians will be provided information about (a) when particular treatments were previously used or rejected by other clinicians, including cohort level and non-PHI individual case-level details, and the rationale(s) for using or rejecting treatments in these patients, and (b) an explanation of the information that could be gained through the use of each treatment expressed as an information to information-gain ratio, which augments the description of the global benefits by quantifying how much can be learned from different treatment options within the equipoise set.

The system uses machine learning to facilitate the recollection of recommendations made for similar patients in the past, but human expertise oversees this at multiple stages. The patient and their doctor can then review the suggestions and supporting evidence and decide without concern about access, knowing that xCures' Patient Access Core team is available to help facilitate access to the selected options. That data will be collected and if a similar patient is identified in the future, it becomes available for reference. Using a Bayesian non-parametric clustering model for example to conduct patient grouping, the system and the VTB experts can learn as new data is generated. The approach includes a prospective element, since the clustering model can create explicit forecasts for patient trajectory with and without various treatment options, for example using Bayesian Additive Regression Trees. This allows for statistically valid inferences to be developed about optimal treatments for specific patient subgroups, and this can be used for supportive evidence both for Patient Access Core operations and in published literature.

This group of patient-centric registries will advance the knowledge base necessary to implement what the FDA describes as patient-focused drug development in oncology. Specifically, they describe this as doing a better job incorporating the patient's voice in drug development, including but not limited to:

- Facilitating and advancing the use of systematic approaches to collecting and utilizing robust and meaningful patient and caregiver input to more consistently inform drug development and regulatory decision-making

- Encouraging identification and use of approaches and best practices to facilitate patient enrollment and minimizing the burden of patient participation in clinical trials

- Enhancing understanding and appropriate use of methods to capture information on patient preferences and the potential acceptability of tradeoffs between treatment benefit and risk outcomes

- Identifying the information that is most important to patients related to treatment benefits, risks, and burden, and how to best communicate the information to support their decision making.

XCELSIOR explicitly addresses these aims at various steps in our process, although the focus of XCELSIOR is a patient-centric registry that builds knowledge about what treatment option is best for specific patients, this is accomplished through a systematic data collection model that incorporates the collection of real world evidence

(RWE) with validated patient and clinician rating scales that are widely used for efficacy and safety monitoring in clinical trials, but less common in clinical practice. In addition, Cancer Commons, xCures, and the Musella Foundation have many anecdotes of clinically meaningful outcomes achieved by patients that could provide direct measures of disease mitigation and patient benefit.

In clinical practice, and for patients enrolled in the XCELSIOR protocol, if a patient has multiple mutations or molecular markers that are logical therapeutic targets, the VTB may propose targeting any or all in combination. Treatments with the strongest rationale including molecular, genomic, clinical, and other supporting evidence may all be considered to create a patient-specific specific plan. These patient-specific plans are a hypothesis and through the analytics within XCELSIOR have both supporting evidence and a prospective forecast for disease trajectory. The plan is not limited to single agents, or even to approved treatments. Patients' best option may be a clinical trial or compassionate use. XCELSIOR supports both. Patients best option may be additional diagnostic testing before a treatment plan is defined. In such cases, XCELSIOR will support additional testing. This is a fundamentally patient-centric approach in which the treatments are matched to the patient rather than matching a patient to clinical trial. Within the XCELSIOR registry model, the patient isn't allocated to specific trial arm that is limited by pre-specification at all; their arm is treatment that their clinician decides will give them the best chance of response. The purpose of the registry is to create a systemic framework so that these experiences, which occur in clinical practice every day, can be mined for data that can improve patient care, by identifying which treatments work or do not work for patients based on identifiable factors.

###### 2.4.1.1 VIRTUAL TUMOR BOARD

XCELSIOR will further develop and leverage the operations of XCures alone or on behalf of a partner or collaborator sponsored Virtual Tumor Board, which operates analogously to an interdisciplinary andmolecular tumor board, but involves the remote review and discussion of patient cases using software. The discussion is facilitated by a moderator, typically a scientist with a background in cancer research. The goal of the VTB is to review an abstracted patient case that incorporates the critical information distilled into a summary of including clinical, surgical, radio- and chemo-therapy, with tumor histopathologic, molecular, pathway, "omics", and pharmacodynamic information, as available.

While the software supporting the VTB will identify prognostic factors for patient outcomes with potential moderators or meditators of patient response to treatment, such as molecular targets for which there are known drugs, and with the input of the scientist/moderator, support the identification of potential therapy options from existing or experimental drugs, alone or in combination. The functional VTB and supporting software will underly the patient Options Core to ensure that high-quality information with supporting rationale is clearly communicated to patients and their physicians.

Ultimately, the analysis of VTB decision-making should make optimal use of molecular and panomic information that can underly a continuously-learning treatment recommendation engine, that can help experts to navigate the relative value of combinations of large numbers of molecular targets and targeted therapies. This is the implementation of the Global Cumulative Treatment Analysis (GCTA) model that aims to maximize the information gain across the global system by analyzing the informational value of every treatment option in relation one another within an equipoise treatment set. Since each XCELSIOR participant is a cancer patient with relatively short expected survival, each patient is a scarce resource, with only two or three treatment options possible. GCTA analysis will augment the VTB equipoise option set by ensuring that each recommendation comes with information about the relative global value of information that may be learned from that treatment, which optimizes patient participation.

##### 2.4.2 Patient Access Core

XCures alone or on behalf of a partner or collaborator will continue to build Patient Access services, to ensure that expert VTB recommendations are easily available for patients and physicians that wish to use them as part of their cancer care. While access support services are not part of the XCELSIOR registry, these activities are enabling activities provided under XCures alone or on behalf of a partner or collaborators' patient advocacy services. Information stored or generated within the Patient Access Core is Services Data, but elements may be abstracted into the XCELSIOR Research Data.

Examples of activities that the Patient Access Core may offer as service to XCELSIOR participants include facilitating access to treatments or testing through partnerships with pharmaceutical, biotech, medical device, and testing companies. These activities are performed in furtherance of ensuring that patients can access the best options, as recommended by the VTB and that to the extent possible, lack of access is not a barrier to patients. This may also include providing transportation or logistical support to patients to ensure they can access VTB recommendations. The information generated may be used as Services Data to ensure that information relevant to securing access to treatment or testing is used for that purpose for the specific patient. Information generated as a result of access to treatment or testing support services may be abstracted into the Research Data. Research Data, including data generated from the Patient Access Core, may be used to assist in the development of new molecular or "omics" tests, or to support developers of drugs and devices to treat cancer, or to promote insurance coverage for the off-label use of anti-cancer therapies. All these activities are consistent with XCures and their partners' and collaborators' advocacy, research, and patient care missions. These partnerships will be done using suitably de-identified datasets.

###### 2.4.2.1 COMPASSIONATE USE CLEARINGHOUSE

To facilitate access to investigational therapies for patients that cannot access those therapies through a clinical trial, XCures alone or on behalf of a partner or collaborator will build a Compassionate Use Clearinghouse (CUC) to support access for patients in the XCELSIOR registry. Because investigational therapies used in a compassionate use setting must be provided under an open IND, the CUC will provide services to patients' physicians to support filing of single-patient expanded access. In addition, under a separate protocol, XCures alone or on behalf of a partner or collaborator will submit INDs to support oncology protocols for both research and treatment, including multiple Intermediate Expanded Access protocols. These activities may require separate protocols and informed consent with separate IRB approval at the site-level, i.e. the patient's physician will be responsible for obtaining IRB approval and informed consent consistent with their institutional policies in coordination with the IND sponsor. Patient data, such as demographics or adverse events, collected in XCELSIOR may then be submitted, exchanged or transferred in a suitable format in support of requirements related to investigational drug protocols for which the XCLESIOR patient participates.

###### 2.4.2.2 OFF-LABEL AND COMBINATIONS FOR TARGETED THERAPIES

In some cases, off-label use of targeted therapies is the medically logical choice, either alone or in combination with other anti-cancer therapies. Patients and physicians sometimes have difficulty accessing these through insurance, as noted above. In cases where such options are recommended by the VTB, processes will be established to support insurance coverage, for example by providing clinical information in support of medical necessity and conducting analysis of coverage decisions for the purpose of improving access for future patients. In addition, the Patient Access Core will work to establish a bridging mechanism whereby sponsors provide drugs off-label for patients while insurance coverage is pending while Cancer Commons collects outcomes to establish individual patient effectiveness in support of insurance coverage.

##### 2.4.3 Data and Analytics Core

XCures alone or on behalf of a partner or collaborator will work to establish a Data and Analytics Core for XCELSIOR that will ensure that software technology infrastructure is established to handle patient intake, tracking, outcomes and analysis. This may include the use of electronic surveys through the Web or via smartphone apps, electronic intake and abstraction of patient health records from external platforms and establish the software and services expertise to manage and analyze the data generated. In addition, as there are methodological aims within XCELSIOR, the Data and Analytics Core may develop and validate new patient-focused outcomes assessment tools.

#### 2.5 Risk/Benefit Assessment

##### 2.5.1 Known Potential Risks

XCELSIOR is a non-interventional study. The purpose is to analyze the case-history for late-stage cancer patients and facilitate the sharing of information related to potential treatment options and supporting rationales with patients and their clinicians. The focus of XCELSIOR is on cases where there are no standard options and survival rates are historically below 20% at five years post-diagnosis. The review process for the patient's case includes the development of potential options and review by an expert VTB, which is essentially a technology-supported second-opinion service. Many cancer patients seek second opinions regarding diagnosis or treatment options and patient-driven second opinions are becoming more common in oncology (Hillen et al., 2017). They are most common in late-stage cancer, where recommended standard options are unavailable. Although the research is far from definitive in this area, it is thought that second-opinions could impact the doctor-patient relationship, which is a potential risk (Hillen et al., 2017). XCures alone or on behalf of a partner or collaborator intends to provide patient advocacy and support services through provision of information or data and will carefully liaison with physician directly and on behalf of patientsseeking assistance from XCures or a partner or collaborator and will make every effort to do so in a collaborative manner that does not jeopardize the doctor-patient relationship.

This include sharing detailed rationales and supportive data from the network with treating physicians and engaging, supporting, if requested, doctor-to-doctor communication between the patient's physician and members of the VTB. Furthermore, while XCures alone or on behalf of a partner or collaborator may share recommendations including sharing information pertaining to clinical trials for which the patient may be eligible or investigation or off-label drug treatments that are supported by a medical rationale, this information is not considered a potential risk because treatment itself is not provided under this study and treatment decisions remain with the patient's physician.

To assess patient cases and provide information about treatment options and treatment assistance, XCELSIOR collects identifiable patient health information (PHI). This information will be kept secure and confidential to the extent possible, but may be used for authorized purposes, including assistance with determining treatment options, communication with physicians, insurers, laboratories, or other healthcare providers about the patient's case or payment for treatments, or as otherwise required by law. Information may also be disclosed under contract to partners, vendors, or business associates of XCures alone or on behalf of a partner or collaborator in relation to the conduct of this study.

##### 2.5.2 Known Potential Benefits

Cancer Commons, operating under the XCELSIOR study framework, intends to continue its mission as a patient-advocacy organization providing information to late-stage cancer patients. While XCELSIOR is non-interventional, patients and their doctors may receive information that supports their medical care, such as suggested treatment options based on molecular, genetic or other information about the patient and their tumor. In such cases, information is shared with the supporting rationale based on expert judgment through the VTB. In some instances, patients' medically logical option, as determined by the VTB may be a clinical trial for which the patient is not eligible due to various factors, including enrollment constraints (e.g., all cohort slots already filled), the severity of their disease or presence of comorbidities, or inability to travel to an active trial site. In those cases, Cancer Commons may aid the patient and/or their physician and depending on the circumstances, this aid may include help to overcome administrative, regulatory, and/or logistical to barriers to treatments either through expanded access or insurance support.

This information may help patients feel empowered within the treatment process and may provide them or their physicians with information about how specific treatments impacted similar patients leading to more informed choices in the absence of accepted standards. If patients and/or their physicians require assistance accessing a selected option, whether it was an option suggested by the VTB, XCures alone or on behalf of a partner or collaborator may provide assistance to help the patient can access the selected option, although access cannot be guaranteed. This may be of benefit to the patient in terms of reducing the barriers to treatment access. Patient participation will help inform the care of other patients collectively by developing precision medicine algorithms to select cancer treatments.

##### 2.5.3 Assessment of Potential Risks and Benefits

The two primary risks of this project are disruption of doctor-patient relationships and loss of patient confidentiality. Regarding the former, the Cancer Commons Ask service has helped more than 2,500 patients to date and disruption of the doctor-patient relationship has not been observed or reported. With respect to the latter, Cancer Commons will take all commercially reasonable and customary efforts to protect patient confidentiality, while ensuring that data collected benefits patients through the options development and access support processes. Future patients will benefit from the outcomes tracking process. The risks are thought to be minimal in accordance with the definition of Minimal Risk in 21 CFR 50 and 56 and outweighed by the potential benefits.

### 3 OBJECTIVES AND ENDPOINTS

The primary objective of XCELSIOR is to collect data into a registry that enables the implementation of a patient-centered, AI-enhanced learning model for optimizing treatment selection in precision oncology. This includes necessary activities to support creation of that model, including processes, tools, and systems to support patient options, including the VTB, including insight capture and analysis of VTB recommendations and physician treatment selection, as well as improving patient access to precision medicine tests and therapies, and collection of associated information with patient safety and effectiveness outcomes in a longitudinal, observation data registry.

Secondary objectives include measurements of safety, tolerability, and effectiveness of therapies measured using outcomes and endpoints derived from RWE supplemented with patient or physician surveys and tracked at the individual patient level and used for aggregated n-of-1 analyses. Patient endpoints will include responder analysis using a composite of clinical response rates measured through disease- or therapy-relevant laboratory changes (e.g., CA19-9 for pancreatic adenocarcinoma), symptomatic improvement, health-related quality-of-life changes, functional performance, therapeutic switching, and radiographic response. Outcomes for specific treatments may be based on, as appropriate, time-to-treatment-switching and/or time-to-progression ratios, as well as response rates measured using durable response relative to historical controls or in relation to prospective forecasts. Additional secondary endpoints may be assessed for comparative effectiveness in relation to traditional clinical trials, including progression free survival at various time points, high-grade complications (SAEs or CTCAE v5.0 grade 3 or higher events)

Exploratory objectives include the development of analytic methods and tools for patient group structure modeling and trajectory forecasting using artificial intelligence and machine learning. To incorporate measures of patient goals and perspectives into the treatment option set development process. To use machine learning to understand and maximize system-wide information gain from each patient treatment decision and outcome. To develop and validate disease-specific Patient Reported Effectiveness and Functional Outcomes, which may include Patient-reported outcomes, including patient statements communications, photo and video submissions, may provide value for identifying treatment response and may be used as generalizable patient-reported outcomes.

### 4 STUDY DESIGN

#### 4.1 Overall Design

XCELSIOR is patient-centric platform study for the registration of cancer patients, operations of a Virtual Tumor Board, insight capture in clinical decision-making, patient access support services, and collection of longitudinal, observational data in disease-specific registries. Patient intake into XCELSIOR will occur through the XCures Web portal, which has been designed to support intake on behalf of partner and collaborator organizations. This process includes authorization to provide advocacy support services and consent to participate in the data registry, including the collection and review of medical information by a Virtual Tumor Board, generation of patient-specific treatment options with supporting rationale, access to treatment access support services, and assignment to a registry study that includes safety and efficacy outcomes tracking. Patients will be treated and tracked in their preferred treatment setting and the data generated will form part of a systematic framework combining expert judgment with artificial intelligence to maximize information gain and improve treatment option set development for individual cancer patients.

#### 4.2 Scientific Rationale for Study Design

To develop information that will enable precision oncology, XCures alone or on behalf of a partner or collaborator believes that three key pieces must be in place. First, patients need treatment options that are matched to their specific condition based on all available information. New tools and algorithms are necessary to accomplish this, including machine learning enhanced by human expertise. Second, access to medically logical options as determined by the VTB and their physician based on advanced testing methods, such as NGS, whether the best option is readily available. Third, outcomes tracking in a longitudinal, observational registry that combines unstructured data from medical records with carefully abstracted, disease-specific information to facilitate assessment of treatment effects continuously and incorporate new information into an enhanced learning model.

The purpose of a patient-centered registry is to serve one or more predetermined scientific or clinical purposes. In this, case the purpose is to advance the development of algorithms that enhance global learning about precision oncology and improve the access of cancer patients to appropriate precision oncology testing and treatments. (Gliklich et al., 2010). While not limited only to patients that are unable to participate in clinical trials, registries are valuable for providing information about the safety and effectiveness of treatments in patients that not typically studied in clinical trials. Furthermore, because a major purpose is to develop methods and systems to advance this field a registry offers more adaptable and flexible approaches to data collection and analysis.

#### 4.3 End of Study Definition

XCELSIOR is a longitudinal, observational study that will run for an initial period through enrollment of 10,000 patients through confirmation of the date-of-death of the last patient in the cohort. The goal will be to develop an infrastructure for a “perpetual protocol” that may be amended to add additional disease-specific elements. Based on the ongoing analysis of data, additional patients may be necessary, if which case the protocol will be amended.

### 5 PATIENT OPTIONS CORE

#### 5.1 Registration of Patients

##### 5.1.1 Patient Registration Process and Schema

###### 5.1.1.1 Patient Registration Process Steps

Patients will be registered for XCELSIOR through the XCures Website, which has been designed to support multiple partners and collaborators, who can refer patients directly or indirectly. Following a patient or legally-authorized representative registering through execution of a patient advocacy agreement, the study registration process follows these steps:

1. Review and completion of an Informed Consent Form to participate in XCELSIOR, including consent to:
    a. Allow their information to be collected and reviewed by the VTB as described in Section 6, if desired
    b. Allow XCures alone or on behalf of a partner or collaborator to ask them or their doctors questions related to their medical care
    c. Allow XCures alone or on behalf of a partner or collaborator to discuss their case and share information in relation to access support activities
    d. Allow XCures alone or on behalf of a partner or collaborator to track their outcome
    e. Allow XCures alone or on behalf of a partner or collaborator to analyze their data and use that data for research purposes
2. Collect patient medical information from EMR, pathology report, imaging, surgical reports, mutation and NGS lab reports, and other information as available as the patient's advocate, a designated third party under the HIPAA individual right-of-access mechanism
3. Collect patient treatment preferences and objectives through a patient utility questionnaire, for example the EORTC QLU-C10D, which is a subset of the QLQ-C30 or similar form
4. Perform case abstraction and generate case summary with overview of clinical, pathology, molecular lab data, treatment history, and current status
5. The abstracted case will provide a baseline for assessment by cluster analysis to identify similar patients and treatment options supported by the information known about the patient
6. Calculate a trajectory forecast for the patient based on the calculated trajectory for the cluster to which they belong
7. Perform treatment option search and calculate overall information gain for options
8. Prepare initial treatment option set
9. Send case summary and initial treatment option set to VTB

###### 5.1.1.2 Patient Registration Schema

#### Patient Intake / Registration

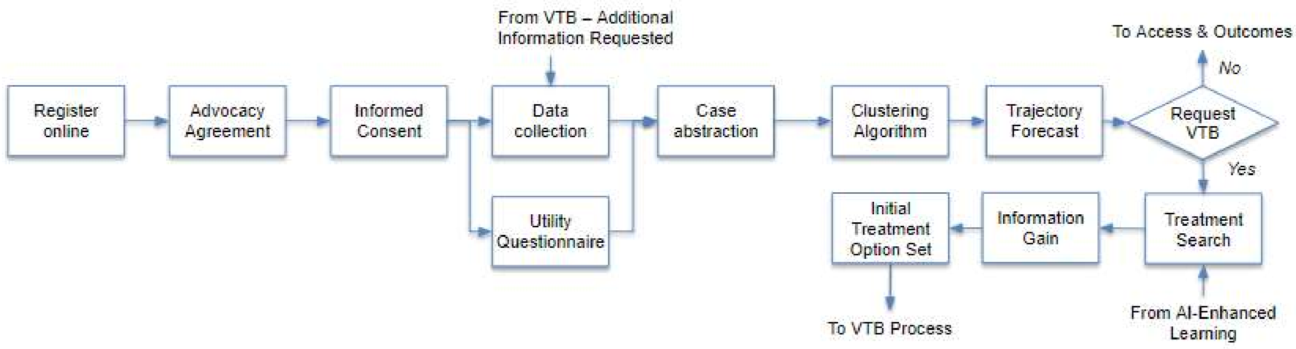

##### 5.1.2 CONSENT/ASSENT AND OTHER INFORMATIONAL DOCUMENTS PROVIDED TO PARTICIPANTS

Patients registering at xCures Web sitewill complete consent forms describing in detail the XCELSIOR registry including the processes, aims, and risks, which will be completed electronically, and a copy will be emailed to the participant. Patients will also complete a Patient Advocacy Agreement which outlines the relationship, responsibilities and obligations of the study team in relation to the patient for Services provided by xCures alone or on behalf of a partner or collaborator prior to initiation of any research activities.

##### 5.1.3 CONSENT PROCEDURES AND DOCUMENTATION

Informed consent is a process that is initiated prior to the individual’s agreeing to participate in the study and continues throughout the individual’s study participation. Consent forms will be Institutional Review Board (IRB)-approved and the participant will read and review the document electronically. Study team personnel or their designees may answer any questions that may arise. Patients will complete a Patient Advocacy Agreement which outlines the role and responsibilities of Cancer Commons and the patient in relation to one another. Participants must sign the informed consent document and Patient Advocacy Agreement prior to participating in the XCELSIOR registry. Participants will be informed that participation is voluntary and that they may withdraw from the study at any time, without prejudice by following procedures outlined on the xCures Website. A copy of the informed consent document will be emailed to participants for their records. The informed consent process will be conducted and documented in the electronic data capture system and stored in a secure 21 CFR part 11 compliant database. The rights and welfare of the participants will be protected by emphasizing to them that the quality of their medical care will not be adversely affected if they decline to participate in this study.

##### 5.1.4 SELECTION OF PATIENTS

###### 5.1.4.1 INCLUSION CRITERIA

As a patient-focused, real-world data registry, any cancer patient or their legally-authorized representative can register and participate in XCELSIOR. To participate an individual must meet all the following criteria:

1. Patient or legally authorized representative able to understand and willing to provide electronically signed and dated Informed Consent. Patient able to provide assent, if applicable.
2. Patient or legally-authorized representative willing and able to provide electronically signed and dated Patient Advocacy Agreement
3. Medical history or diagnosis of cancer

###### 5.1.4.2 Exclusion Criteria

XCELSIOR is a patient-focused, real-world data registry. Patients that meet the inclusion criteria and wish to participate, as indicated by signing an informed consent form will not be excluded.

###### 5.1.4.3 Screen Failures

Screen failures are defined as participants who consent to participate in the clinical trial but do not meet the inclusion criteria. Specifically, people that register and complete the intake procedures but do not have a medical history or diagnosis of cancer will be considered a screen failure. A minimal set of screen failure information is required to ensure transparent reporting of study results, including to meet publishing requirements and potentially to respond to queries from regulatory authorities. Minimal information includes registration and patient intake information, medical information abstraction and review, demography, eligibility criteria and screen failure details, and any serious adverse event (SAE) or outcome information.

Individuals who do not meet the criteria for participation in this trial (screen failure) for example due to a lack of a cancer diagnosis may be rescreened and may enroll in the XCELSIOR registry later if they meet the entry criteria. Rescreened participants will be assigned the same participant number as for the initial screening.

#### 5.2 Virtual Tumor Board Operations

##### 5.2.1 Virtual Tumor Board Overview

XCures acting alone or on behalf of a partner or collaborator will support activities by using a “Virtual Tumor Board” or “VTB”. The operations of the VTB are in many respects analogous to the tumor boards run at major cancer centers or molecular tumor boards described in section 2.2.2. The processes outlined in this study protocol describing the VTB are information, as the function of the VTB is to provide individual information services to patients and their physicians. However, research will be conducted on the decision-making process to understand the rationale underlying VTB recommendations in relation to the tracking of patient outcomes within XCELSIOR. Information derived from research on treatment rationales will be available to the VTB to facilitate treatment option identification and associated outcomes as part of the overall learning process.

##### 5.2.2 Virtual Tumor Board Process

Following registration and consent, XCures acting alone or on behalf of a partner or collaborator will contact the patient to begin collection and abstraction of their medical information into a patient summary to be provided to the VTB. This process includes identification of similar patients medically-logical treatment options, including clinical trials, that have been suggested by the VTB for similar patients or have otherwise been identified as medically logical for similar patients, with relevant information from the study Leaderboard, if available. The completed summary is then submitted to at least three VTB members selected based on their availability and expertise for review. The VTB using will review the case, options, and discuss using on- or off-line technology support, to reduce the options into a list of best options at clinical equipoise. Rationale capture for addition, inclusion, or removal is collected for global learning analysis. Additional rounds of review and discussion may be undertaken to further refine the option list and VTB members may request additional information, including review of source records, to support their review. Once the option set is reduced the patient utility is applied and simulation analysis is run to assess the global information gain value of the options in the equipoise set. Together these are provided to the VTB to determine whether they then wish to update their selection of options. Finally, the refined option list, with supporting information and rationales, plus information gain value and alignment with patient preference are provided to the patient and their doctor for consideration.

##### 5.2.3 VTB Operations Schema

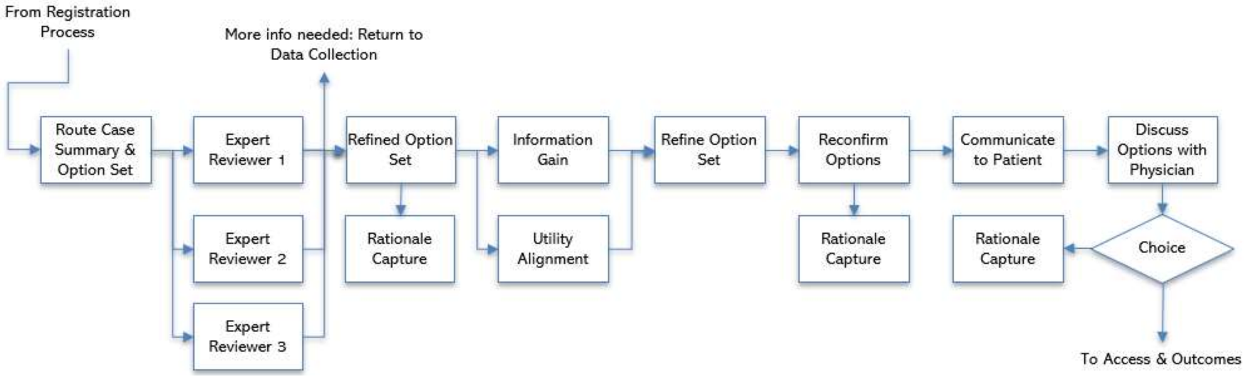

##### 5.2.4 Qualification and Selection of VTB Members

Virtual Tumor Board members are independent experts that work with XCures acting alone or on behalf of a partner or collaborator to provide review and assessment of patient cases. Members are selected based on clinical and academic expertise in the treatment and study of specific tumor types. Members of the VTB will include appropriately qualified individuals representing clinician disciplines, including medical, surgical, or radiation oncologists, as well as pathologists and/or radiologists; pharmacologists; molecular, cancer or systems biologists; and others, as needed to fulfil the VTB function. Members should be free of conflicts of interest in accordance with the policies described herein and in compliance with applicable regulations.

##### 5.2.5 VTB Financial Disclosure and Conflict of Interest Policy

VTB members of should be free of conflict of interest and measures will be implemented to minimize perceived conflict of interest. Specifically, the VTB will operate under the rules of an approved charter or SOP, and discussions will be documented and archived for future analysis. VTB members should disclose to XCures acting alone or on behalf of a partner or collaborator real or perceived conflicts-of-interest, which will be documented at the time of their appointment. VTB members should refrain from reviewing cases for which they believe they have a conflict-of-interest. Members will serve a two-year term of service, which may be renewed indefinitely. Upon renewal of service, VTB members will update their disclosures and from time-to-time as necessary for the publication of findings in which they have been named as an author. VTB members should comply with their institutional and other state and local conflict-of-interest laws, regulations, or policies. XCures acting alone or on behalf of a partner or collaborator will review disclosures against public information maintained by the Centers for Medicare and Medicaid Services (CMS) in the OpenPaymentsData on-line database and search engine.

##### 5.2.6 Relation to vDMC

Members of the VTB may participate in the vDMC, the purpose of which is to review the overall data generated in XCELSIOR and identify insights for dissemination.

##### 5.2.7 Authorship and Publication Committee

VTB members that are involved in the review of patient cases may be named as authors on publications which result from analysis of those cases. Other personnel involved with the collection, management, analysis, or interpretation of data will be named as authors either individually or in the form of group authorship with their contributions identified in accordance with ICMJE policy.

The VTB and vDMC may assemble ad hoc publication committees to author posters or publications on specific topics as data from the registry generates meaningful information. XCures, as the sponsor, will have overall responsibility for the management, analysis, interpretation, and publication of the data. Therefore, Cancer Commons will appoint representatives to serve on all publication committees to ensure that the perspectives of patients and advocates are incorporated in the dissemination of information and retains ultimate authority over these activities.

### 6 PATIENT ACCESS SUPPORT CORE

The purpose of the Patient Access Core is to provide support and assistance to ensure that access to medically-logical options suggested by the VTB are available for patients.

There are many barriers to access facing late-stage cancer patients and for these patients especially, time is of the essence. These barriers can be administrative, logistical, geographic and financial. However, patients are the most valuable resource in precision oncology. Barriers to access not only cost lives but the prevent essential learning that will help other patients. Thus, a continuously-learning model for precision oncology must ensure that patients receive medically-logical treatments and that the results of that treatment are captured for learning purposes.

To overcome these barriers, XCures acting alone or on behalf of a partner or collaborator will attempt to create a clearinghouse for patients trying to access therapies either through clinical trials or through expanded access programs. This clearinghouse would facilitate the matching of targeted anti-cancer therapies made available in clinical trials or under compassionate use to patients and their doctors. This type of central player is absent in the compassionate use arena, leading to the many challenges matching patients to trials. Although there are firms doing trial matching with varying degrees of success, there are many more barriers that must be overcome. For example, after matching patients to potential trials, patients must be transferred to sites for screening, which isn't typically covered by sponsors or insurers. This, ensuring financial support for travel is required—medical bills, and in particular, cancer treatment is a leading cause of personal bankruptcy in America (Murphy, 2018).

The patient access core can provide liquidity by helping to match patients to trials and prevent failures by stepping in when patient can't access a trial to assist with getting them a medically-logical therapy and collecting the data in a registry. That data may prove of use later both directly and indirectly to patients or sponsors. Conversely, sponsors may be unwilling to risk trials for certain patients due to fear of failure but could provide single patient drugs under compassionate use terms, which can immediately help patients and build evidence to support larger trials later.

Similarly, XCures acting alone or on behalf of a partner or collaborator with XCELSIOR can facilitate pay-for-results between insurers and sponsors in cases where a patient may benefit from the off-label use of a drug, but a payor is unwilling to cover the drug without evidence of medical necessity. In such cases, XCELSIOR can work with willing sponsors to provide a bridge of drug access to a time point in where evidence of drug effect can be assessed and provided as support. During that time, negotiations with an insurer can take place, but the patient does not have to defer treatment. Similarly, private oncology practices sometimes end up taking on this risk, by procuring and administering treatment under the hope that the insurer will agree to compensate them at a later date. This arrangement is becoming less common as the costs of treatments increase. However, this type of pay-for-results market-making can be of benefit to patients, insurers, and manufactures at the same time.

These activities represent a service, however information collected or generated in the process of providing access services may be used as part of research and publication efforts. For example, Sireci and colleagues (2017) reported on the rate of insurance reimbursement received for patients receiving an off-label targeted cancer therapy identified by their organizational molecular tumor board. The information collected to support patient access services are Services Data rather than Research Data, but specific information about Services provided to patients participating in XCELSIOR and associated Services Data may be abstracted into the Research Database where such information becomes part of the specified Research Data elements. For example, if travel funds were provided to a registry participant to facilitate enrollment in an externally-sponsored clinical trial, information about the patient's participation in that trial will be collected.

#### 6.1 Treatment Access Support

Acting on behalf of Cancer Commons or other partners and collaborators, xCures may receive commercially available drugs directly from sponsors/manufacturers for the purposes of providing expedient treatment access services to patients. XCures acting alone or on behalf of a partner or collaborator many negotiate directly with sponsors to provide a supply of approved drugs to be stored at a central pharmacy provider, which may then provide matched therapies to sites designated for administration to specific patients participating in XCELSIOR. This process is envisioned in two primary ways. First, sponsors may provide drugs for patients eitheras bridging therapy, enabling patients to start treatment rapidly without concerns about payment while XCures acting alone or on behalf of a partner or collaborator works with patients' physicians to secure insurance coverage due to medical necessity. It is expected that within two months of bridging therapy, information related response through clinical, imaging, or laboratory testing would be developed and form the basis of support for effectiveness in a specific patient. This information would support insurance coverage without delaying the start of treatment. In addition, analytic methods may be incorporated to demonstrate effectiveness in similar patients based. Second, sponsors may provide drug directly out of altruism, in which case the donation may be a charitable contribution to a 501(c)3 entity, or in exchange for access to de-identified data that improves their understanding about which patient characteristics correlate with effectiveness. In addition, XCures acting alone or on behalf of a partner or collaborator will attempt to help patients enroll in co-payment support programs and may raise a charitable fund to help patients with insurance cover co-payments when external copayment support is unavailable.

For patients that are matched to clinical trials, XCures acting alone or on behalf of a partner or collaborator may notify sponsors in cases where a patient faces a logistical, geographic, or financial barrier preventing them from traveling to a clinical trial site. In those cases, XCures acting alone or on behalf of a partner or collaborator may work with the sponsor to provide travel support, open a clinical trial site for the patient that is more convenient, or if the treatment is medically logical, but a clinical trial isn't possible, to work with the sponsor and patient's oncologist to provide the treatment under compassionate use.

Treatment Access Support Services are services provided to patients by XCures acting alone or on behalf of a partner or collaborator or their designee, agent or affiliate to further their mission of making treatment options available to patients and their doctors. Services activities are considered distinct from research activities, but information about Services provided by XCures acting alone or on behalf of a partner or collaborator may be abstracted into the Research Dataset if those activities include information pertinent to the registry, such as the name or dose of an anti-cancer treatment provided to a patient's physician for administration on a compassionate use basis. In the event that provision of access to therapies involves investigational treatments, such treatments will be administered by the patient's physician under and approved IND and study protocol specific for the use of such treatments that is distinct from the XCELSIOR data registry.

#### 6.2 PATIENT ACCESS SUPPORT SCHEMA

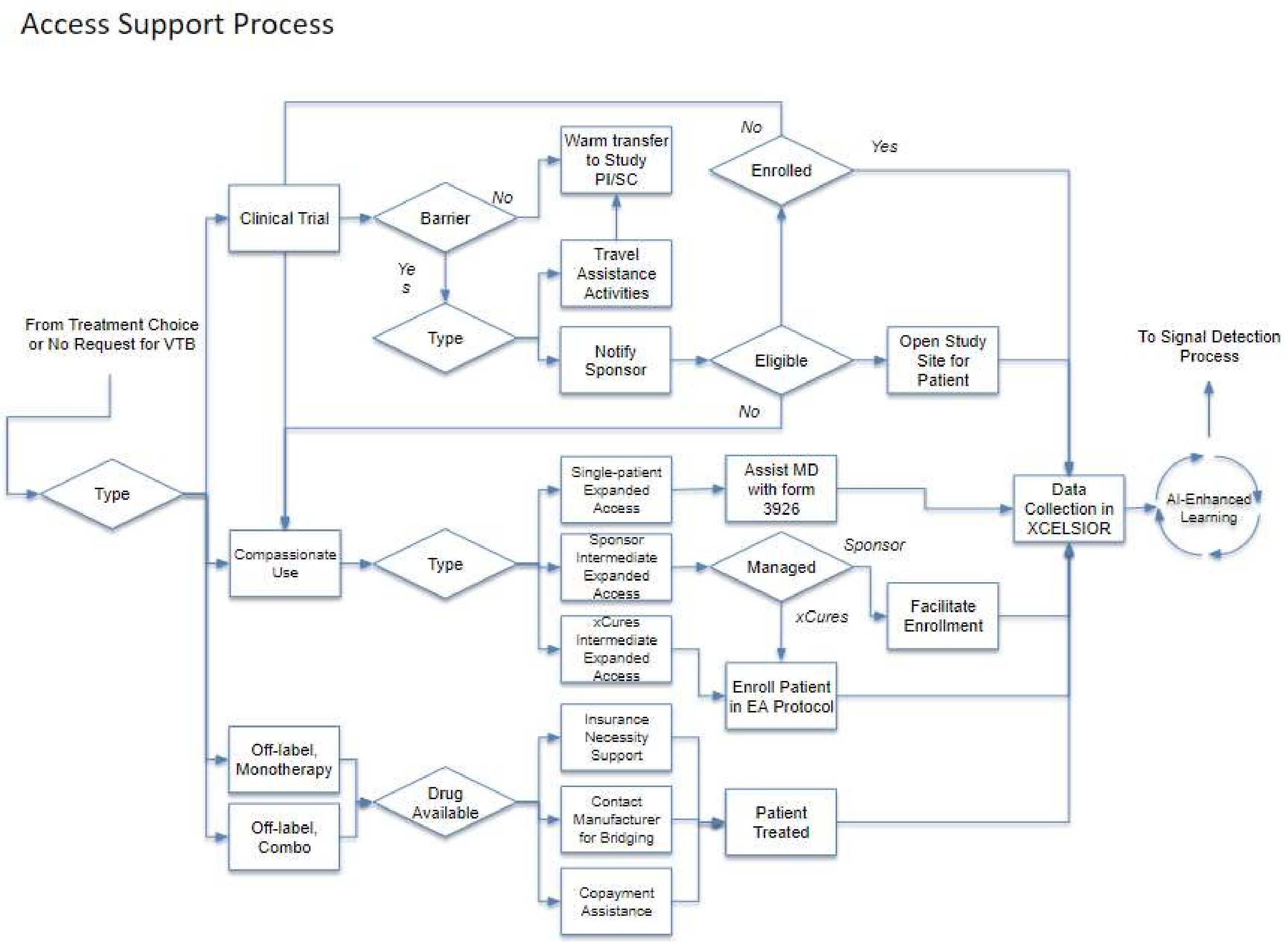

### 7 PATIENTS OUTCOMES CORE

#### 7.1 Data Handling and Record Keeping

##### 7.1.1 Data Management

A registry database will be maintained with identified registry data elements captured into the REDCap Cloud EDC system, a commercial eClinical Platform that is 21 CFR Part 11 Validated, HIPAA & FISMA compliant, and WHODrug and MedDRA certified. The system All access and activity in the system is tracked and can easily be monitored by the Administrator. The system has a login audit feature that tracks who has logged into the system, the date and time of login, and the IP address of the connection. The system also tracks failed logins and automatically locks a user's account after several failed attempts. The REDCap Cloud system has a robust audit trail that shows all changes to any records within the system including who made the change, the date and time of the change, the field that was changed, the old value of the field and the new value of the field. Access can be monitored via a dashboard or email alerts. Data management activities will follow standard operating procedures.

##### 7.1.2 Study Records Retention

Study data including source records and case reports forms will be maintained in electronic format indefinitely, but at a minimum until two years after the last participant is deceased. De-identified databases derived from the study records may be made available to other researchers in the future and may be retained by those organizations indefinitely based on the terms of the agreement under which the data was provided.

##### 7.1.3 Source Documents

Source data can include clinical findings and observations, or other information incorporated into the registry database to support analysis of outcomes, effectiveness, and safety aims. Source data is all information from which information in the registry database is derived in original form (or certified copies of an original record). Examples of these original documents and records include but are not limited to the following: electronic medical records, clinical and office charts, laboratory notes, memoranda, correspondence, subjects' diaries or patient-reported questionnaires, data from automated instruments, such as ECG machines, photographs and other imaging (DICOM) files, slides, pharmacy records, and the reports documenting medical interpretation of those files.

Data may be entered into the eCRF either manually by a study team member performing data abstraction from the EMR or electronically using direct entry of data into the or from an electronic import of data. Patients may also enter information directly into the eCRF using a patient-facing survey functionality either over the Web or using a smartphone app. Data elements originating in EMR may be automatically transmitted directly into the eCRF using a suitable API. Source data derived in that manner may have an intervening process, such as abstraction by third-party (e.g., Ciitizen) software including processing using machine learning algorithms prior to transferring to the eCRF. XCures acting alone or on behalf of a partner or collaborator will retain source records in a secure that will be maintained separately and securely from the eCRF. However, metadata tagging may be employed to electronically map source data back from the eCRF to the source data record to create an audit trail.

Upon registering for XCELSIOR at the Web site, patients under their right-of-access to their medical records may authorize XCures acting alone or on behalf of a partner or collaborator to receive any and all historic medical records available, as necessary to assess and evaluate their case history and generate recommendations through the VTB process.

###### 7.1.4 Data Elements for Collection

1. Demographics (e.g., name, date of birth, gender, ethnicity, race, contact information, name relationship and contact information for legally-authorized representative.
2. Medical Record Information (e.g., MRN, Primary Hospital, Hospital Address, Primary Doctor, Contact email, Contract number)
3. Diagnostic and Clinical Information (histopathology, imaging, symptoms, DRG, clinical examination results)
4. Laboratory Information (clinical labs, cytogenetic, molecular, NGS or other test results)
5. Anti-cancer Therapy (Radiotherapy, Adjuvant/Neoadjuvant therapy, Chemotherapy, Surgical treatment, Device treatments, supportive care, dietary or lifestyle interventions)
6. Safety and Tolerability (side-effects, hospitalization, AE, SAE)
7. Outcomes (Functional, Performance, Quality-of-life, Response measures, mortality)

Specific measures will be incorporated that are appropriate for the tumor type or disease under study. For example

##### 7.2 Anti-Cancer and Other Therapies or Treatments

For this registry information about anti-cancer treatments and related, supportive care, as well as treatment administered in response to CTCAE grade 3 or more severe adverse events will be abstracted from the medical record and stored within the CRF. In cases where an anti-cancer treatment is not considered a prescription treatment (e.g., use is not restricted to prescription only by a properly authorized/licensed clinician) such treatments will be collected to the extent possible using information in the records or provide by the patient or their physician. Examples include patient-reported regimens that include vitamins, supplements, herbal or complementary or alternative medicines taken explicitly for anti-cancer or supportive care purposes.

###### 7.2.1 Treatment Switching

Since the purpose of this study is to develop information that supports personalized medicine in oncology, patients and their clinicians may switch or modify treatments in response to clinical outcomes. Treatment switching will be collected and mark an event for outcomes assessment purposes. The cause of treatment switching will be identified via discussion with the patient and their physician in order to code the reason for switching such as side-effects or lack of efficacy. The reason for treatment discontinuation or switching will be recorded in the eCRF. Patients that switch treatments may request that their case be reassessed through the VTB process.

#### 7.3 Discontinuation of Participation

Discontinuation of treatments does not mean discontinuation from the study, and patients in XCELSIOR have the option of returning their case to the VTB process for new or additional options development if they experience side-effects or progress on a chosen therapy. Furthermore, for late-stage cancer patients, hospice is considered a likely option during the study. Patients are not considered discontinued or withdrawn from the study if they choose to receive hospice care. Patient information will be collected until death, unless the patient chooses to withdraw from the study. Patients are free to withdraw at any time by providing notice of withdrawal in writing using established procedures documented on the xCures Web site and including an option to provide a reason for withdrawal from the study. Withdrawal from the study means that no additional data will be collected on the patient (excluding public records searches to confirm dates of death) and does not constitute revocation of XCures acting alone or on behalf of a partner or collaborator right to use the data collected prior to withdrawal for the purposes described herein.

##### 7.3.1 Participant Discontinuation/Withdrawal from the Study

XCures acting alone or on behalf of a partner or collaborator or their authorized designee may discontinue a participant or withdraw a participant from the XCELSIOR study for any reason at any time.

##### 7.3.2 Lost to Follow-Up

As a patient advocate, XCures acting alone or on behalf of a partner or collaborator frequently contacts patients to share information and assist with discussions related to care, coverage, or responses to treatment. If a patient is unreachable, XCures acting alone or on behalf of a partner or collaborator may continue to attempt to contact the participant, their doctor (or other members of their treatment team), or their designated Emergency Contact by phone, email or similar method. Patients that are not reachable will be considered lost to additional follow-up. Patients that choose to receive hospice care, will not be contacted further. In either case, XCures acting alone or on behalf of a partner or collaborator or designee may undertake public records search

#### 7.4 Virtual Data Monitoring Committee

A subset of the VTB members will be designated as the virtual Data Monitoring Committee (vDMC), which will operate remotely and be charged with periodic review and discussion of safety and effectiveness signals within and among groups and subgroups of patients in the XCELSIOR registry. The vDMC will be empowered to make determinations about:

- adding or removing treatment options from additional consideration for subgroups, based on futility or harm—such determinations should be publicly reported through peer-review mechanisms promptly,

- treatment options for subgroups that no longer belong in an equipoise option set because they are clearly effective, such options may be graduated to other research endeavors, such as GBM-AGILE for randomized clinical trials, or may be the subject of analysis and publication to ensure dissemination to stakeholders.

The vDMC may identify safety signals which, while not altering treatment recommendations for a population subgroup, should be disseminated through publication and notification to clinicians, sponsors or manufactures, and the public. Related to this is information developed in support of mitigation or management strategies including supportive care recommendations identified in relation to adverse events assessed by the vDMC. The initial inclusion of potential options does not require approval by the vDMC and any treatment for which there is medically-logical evidence may be considered by the VTB members. The sequential review by the scientist, Medical Director, and VTB may provide independent adjudication of imaging or endpoints to supplement information reported by the patient or their physician or abstracted from their medical record.

##### 7.4.1 Signal Detection Process and Schema

Signal detection activities will be based on composite endpoints, as well as outcomes in relation to a baseline trajectory forecast. Because patients with late-stage and recalcitrant cancers have no proven options and only a short time in which to explore options, XCELSIOR aims to rapidly assess signals of effectiveness and safety/tolerability through composite assessment of changes in direction, magnitude, or rate of change in labs, images, disease symptoms, patient functioning, side effects, and/or changes in pharmacotherapy. Analytic methods, including traditional methods and machine learning may be used to identify and assess changes in status, which will be combined with human expert judgment to evaluate and classify signals.

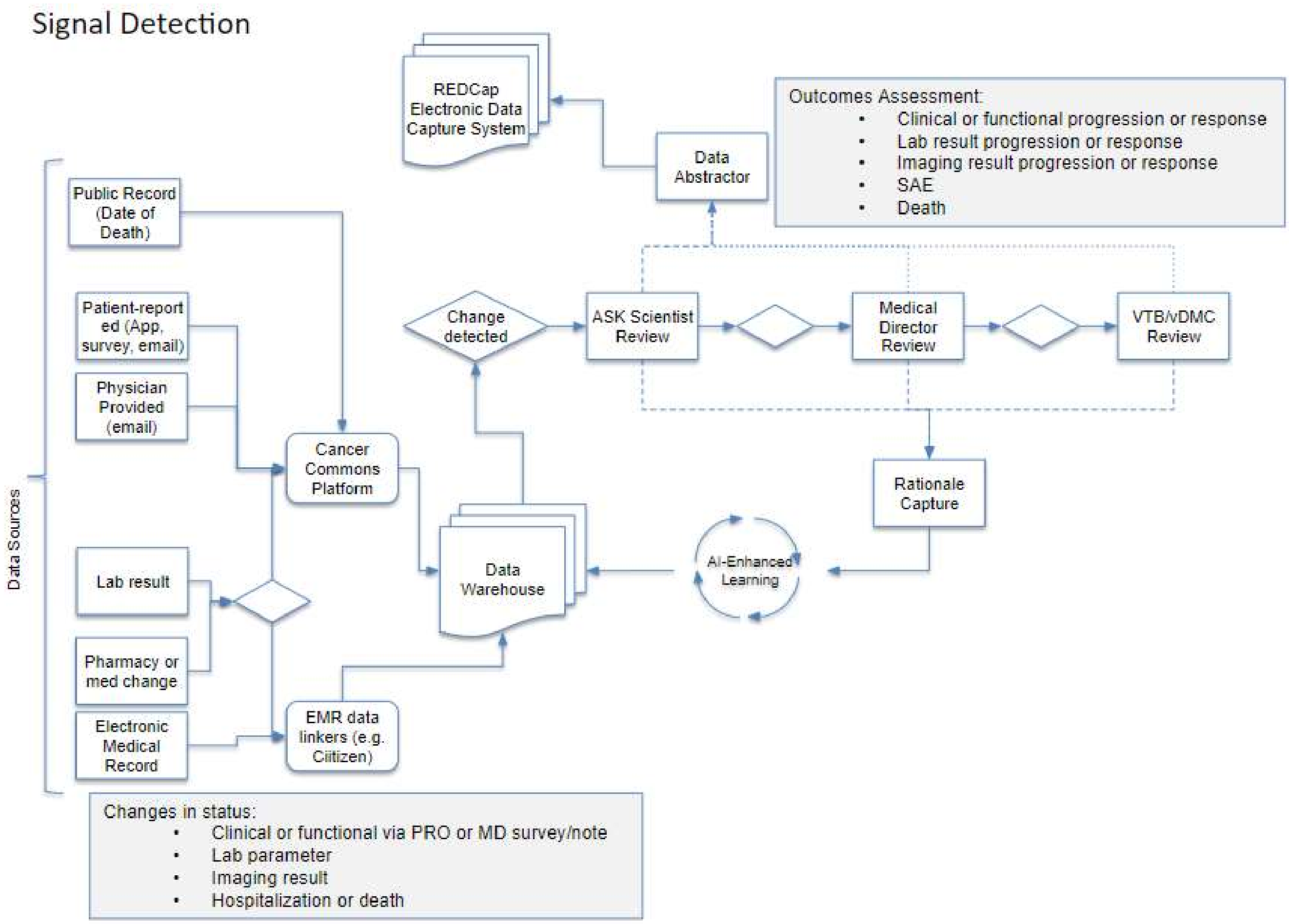

##### 7.4.2 Qualifications and Term for vDMC Members

Members of the vDMC will be drawn from the VTB supplemented with appropriately qualified individuals representing patients or advocates; clinicians; statistical or data scientists; pharmacologists; molecular, cancer or systems biologists; and others, as needed to fulfil the vDMC oversight function. Members of should be free of conflict of interest, or measures should be in place to minimize perceived conflict of interest. The vDMC will meet at least semiannually to assess safety and efficacy data and will operate under the rules of an approved charter or SOP, that will include keeping of written meeting minutes of deliberations. Members will serve a two-year term of service, which may be renewed indefinitely and must comply with the conflict-of-interest policy.

#### 7.5 Adverse Events and Serious Adverse Events

##### 7.5.1 Definition of Adverse Events (AE)

The XCELSIOR registry is not a treatment protocol, it is a non-interventional registry study. However, to facilitate the generation of real-world evidence for the effectiveness and safety of cancer treatment regimens, information about spontaneously reported adverse events or events identified through the processing of the patient medical or health records will be identified analytically based on the emergence of the event in relation to its presence or absence in the medical record at prior time points. Events may also be reported directly by the patient or their physician using surveys or directly through correspondence with XCures acting alone or on behalf of a partner or collaborator and/or xCures. In such cases, reasonable efforts will be made to gather information about the event.

Adverse events will mean any untoward medical occurrence associated with the use of an intervention whether or not considered intervention-related (21 CFR 312.32 (a)). Coding of AEs will be based on the CTEP dictionary (https://ctep.cancer.gov/protocolDevelopment/codes_values.htm#disease)and the severity of AEs will be graded based on CTCAE (https://ctep.cancer.gov/protocolDevelopment/electronic_applications/ctc.htm). In both cases, reasonable efforts will be made to code the events in accordance with standard dictionaries if the information is available directly or by imputation.

The primary focus of the Research Data will be on treatment-emergent events that meet the CTCAE criteria of Grade 3 or higher, therefore these events will be captured and assessed in the registry database to the extent possible. Assessment and analysis will focus on identifying treatment-emergent adverse events in relation to anti-cancer treatments identified within the patient's medical record, particularly those related to dose modification or interruption of anti-cancer therapies, or other therapeutic intervention or modification, including hospitalization or other medical treatment. Discontinuation of therapy due to lack of efficacy will be treated as a therapeutic outcome not an Adverse Event.

##### 7.5.2 Definition of Serious Adverse Events (SAE)

In order to enable comparison with conventional clinical trial databases and to provide real-world evidence about the effectiveness and safety of anti-cancer treatments, XCELSIOR will capture serious adverse event information, when AEs within the medical record (or reported by the patient or their physician) appear to meet the criteria for seriousness, if, in the view of either the patient's physician, the event results in any of the following outcomes: death, a life-threatening adverse event, inpatient hospitalization or prolongation of existing hospitalization, a persistent or significant incapacity or substantial disruption of the ability to conduct normal life functions, or a congenital anomaly/birth defect. Important medical events that may not result in death, be life-threatening, or require hospitalization may be considered serious when, based upon appropriate medical judgment, they may jeopardize the participant and may require medical or surgical intervention to prevent one of the outcomes listed in this definition.

##### 7.5.3 Relationship to Underlying Disease or Treatment

Adverse events that meet the tracking criteria (CTCAE 3 or higher and are treatment-emergent) will have relationship to the patient's underlying disease or treatment assessed if that assessment is available from the patient's physician and coded to the following rubric:

- Definitely Related
- Probably Related
- Possibly Related
- Unlikely to be related
- Not Related

Relationship to the treatment or underlying disease will be captured in the Research Data. Physicians may be queried directly for this information. The final determination will be made using the Signal Detection process and can include input from the patient, their physician, information from the medical record, and review and discussion by XCures acting alone or with partner or collaborator personnel and the VTB/vDMC members.

###### 7.5.4 Expectedness

Adverse events that meet the tracking criteria (CTCAE 3 or higher and are treatment-emergent) and are thought to be treatment-related will be assessed against known information for the treatment and then assessed for expectedness. Events are considered unexpected if the nature, severity, or frequency of the event is not consistent with the risk information previously described for the treatment. Physicians may be queried directly for this information. The final determination will be made using the Signal Detection process and can include input from the patient, their physician, information from the medical record, and review and discussion by XCures acting alone, on behalf of, or with partner or collaborator personnel and the VTB/vDMC members.

###### 7.5.5 Reporting Events

Adverse events may be reported by XCures acting alone, on behalf of, or with partner or collaborator to sponsors or manufactures, to health authorities and regulatory agencies, and to the general public via publication or other form of communication.

###### 7.5.6 Reporting Events to Participants

As a patient-centric registry, safety information may be made available to new and ongoing registry patients and/or their treating physicians as well as the general public.

##### 7.6 Unanticipated Problems

###### 7.6.1 Definition of Unanticipated Problems (UP)

Unanticipated problems involving risks to participants or others to include, in general, any incident, experience, or outcome that meets **all** the following criteria:

- Unexpected in terms of nature, severity, or frequency given (a) the research procedures that are described in the protocol-related documents, such as the Institutional Review Board (IRB)-approved research protocol and informed consent document; and (b) the characteristics of the participant population being studied;
- Related or possibly related to participation in the research ("possibly related" means there is a reasonable possibility that the incident, experience, or outcome may have been caused by the procedures involved in the research); and
- Suggests that the research places participants or others at a greater risk of harm (including physical, psychological, economic, or social harm) than was previously known or recognized.

###### 7.6.2 Unanticipated Problem Reporting

XCures acting alone, on behalf of, or with partner or collaborator will report unanticipated problems (UPs) to the Genetic Alliance Institutional Review Board (IRB) in accordance with the IRB's policies and procedures. The UP report will include the following information:

- Study identifying information: protocol title and number;
- A detailed description of the event, incident, experience, or outcome;
- An explanation of the basis for determining that the event, incident, experience, or outcome represents an UP;
- A description of any changes to the protocol or other corrective actions that have been taken or are proposed in response to the UP.

To satisfy the requirement for prompt reporting, UPs will be reported within 10 working days of the XCures acting alone, on behalf of, or with partner or collaborator learning of the event.

##### 7.6.3 Reporting Unanticipated Problems to Participants

Unanticipated problems will be reported to registry participants that are affected by the UP within 30 days of XCures acting alone, on behalf of, or with partner or collaborator becoming aware of the problem.

### 8 ANALYTIC METHODS AND STATISTICAL CONSIDERATIONS

#### 8.1 Overview of Analytic Methods

Patient-focused precision oncology relies on bringing together several well-defined statistical methods into a framework that is focused on determining which treatment is best for a specific patient and integrating all information into a continuous-learning model. This model is hypothesis-driven at the patient-level, rather than study-level. At the study-level, XCELSIOR operates as a discovery engine using continuous learning and reassessment methods to identify factors that influence treatment response or outcomes for cancer patients. At a patient-level, the VTB objective is to provide services to patients and their physicians that delivers suggestions about the best treatment options for the specific patient using all available baseline co-variate information and develop algorithms that incorporate outcomes from similar patients, patient preferences, and global information gain assessments. Together enable precision oncology, combining individual hypotheses that combine molecular and clinical inferences from the VTB with computational approaches to classification of prognosis for individual patients.

The analysis of XCELSIOR Research Data will lead to new information about emerging signals from treatments in late-stage and recalcitrant cancer, as well as the development of new analytic methods to organize and integrate the results of patient treatment along with the underlying rationale supporting treatment recommendations and decisions.

##### 8.1.1 Cluster Analysis and Patient Classification

Once patient data has been abstracted, a cluster analysis is required to organize and group patients along characteristic dimensions that are relevant to treatment response. The earliest clustering algorithm that has widely been used in medicine is k-means clustering, which uses Euclidian distance to partition groups of observations together such that the distance between group members is minimized while the distance between centers of groups is maximized. The clusters are then defined by linear partitions that form along the borders of the groups in n-dimensional space. This method has been used to create evidence-based subgroups within different tumor types, for example GBM. However, because this approach struggles with very-high dimensional analysis, other methods for reducing dimensionality may be combined, for example Principal Components Analysis, which has also been used along and in combination with k-means clustering for glioblastoma. More advanced clustering techniques will be explored as part of the methods development aims to implement the optimal strategy for patient classification for the purpose of precision oncology. This could include Bayesian Latent Class Trajectory Analysis, K-nearest neighbor in combination with Bayesian NP Clustering, deep neural networks, and/or Random Forest (Decision Tree Ensemble Method).

Brennan and colleagues (2009) used clinical information, including histopathology, treatment, and outcome with proteomic analysis of GBM samples to perform cluster analysis. They identified three major clusters, with the most important axes being PDGFR expression, EGFR expression and loss of NF1 signaling, which they then extended by analysis of genomic alterations that characterized each cluster. By clustering patients using NGS and other data, identification of likely driver mutation pathways can provide information to the VTB to recommendation of targeted therapies. The following figure from that article shows the utility of such classification schemes.

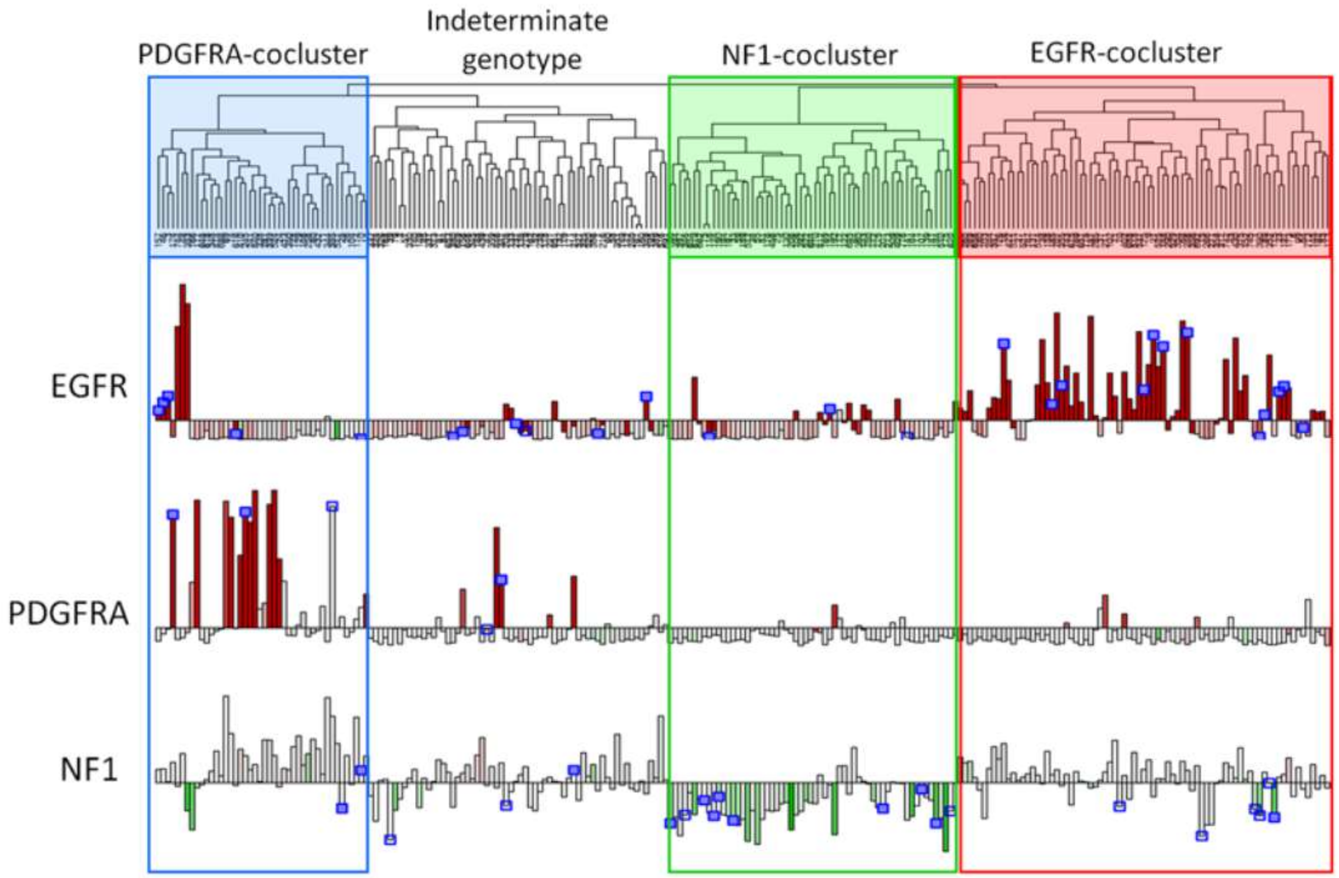

**Figure 6.** Unsupervised clustering of GBM. From Brennan et al., 2017.

This approach has been used by Wang and colleagues (2017a) to show three distinct subgroups of glioma based on genomic and transcriptomic profiling, that differ in their prognosis and treatment response. They term these subgroups, proneural, mesenchymal, and classical, which are characterized by transcriptional differences, as well as the presence of various tumor-associated cells in the tumor microenvironment. They further examine progression and the corresponding changes in the transcriptome of tumor cells between diagnosis and first progression. Similarly, Wang and colleagues (2017b), demonstrate that therapy alters the tumor genome through an evolutionary process and further traced the lineage of dominant tumor clones over time using Bayesian analysis. In glioblastoma, hypermutational states are common in IDH-wild type GBM following relapse after temozolomide. Hypermutated GBM has been associated with responsiveness to immunotherapy. Prior treatments, with histopathological and NGS information, are thus important co-variates for classifying patients.

For XCELSIOR, the goal is a continuously-learning model, which means that the overall patient clusters need to be recalculated continuously as new information is generated. The principal intent is that the clusters should consider clinical, panomic, and other information to build logical groupings of patients that provide informative structure for predicting baseline trajectory and likely treatment response. XCELSIOR will explore various clustering approaches common in machine learning, specifically the use of Bayesian non-parametric (NP) clustering. This approach has several advantages for this application. First, this approach functions similarly to a generalized k-means clustering model, but unlike k-means clustering Bayesian NP models 1) do not require that the covariate relationship between clusters to be zero, 2) does not require the pre-specification of the number of clusters in advance, 3) can handle high dimensional data without the need to reduce dimensionality, and 4) make no underlying assumptions about the shape of the data (Kulis and Jordan, 2012). Bayesian NP cluster models can also incorporate new information and make updated estimates of clustering structure continuously. They can also map two different datasets onto a common group of clusters, supporting learning models that begin with other datasets.

Another valuable aspect of Bayesian hierarchical models can incorporate prospective forecasts with the non-parametric clustering with Latent Class Models (Neelon et al., 2012; Leiby et al, 2009). Multivariate latent class models can integrate multiple response variables into a single latent variable trajectory as part of a Bayesian classification model. This approach enables patient cluster models to explicitly incorporate a baseline trajectory under different treatment conditions (i.e., responders or non-responders to a treatment within a cluster along with associated response trajectory). This approach was applied in a pediatric rare disease by Lim and colleagues (2017).

Within these Bayesian models, historic information regarding outcome can be integrated into a latent variable, to create a latent class model. This approach enables the trajectory of an outcome variable with covariate information to be incorporated into the hierarchy, which is relevant to matching patients to a group for which they are both similar and likely to have a similar response. The choice of outcome variable may include several possible choices that are both context and disease-specific. These include changes to clinical parameters, such as functional performance, quality-of-life or patient reported outcomes, and adverse effects. In addition, changes to laboratory parameters, such as molecular markers, disease markers (e.g. CA 19-9), panomic tests (e.g. circulating tumor DNA), as well as imaging changes may all factor in to a response assessment. A latent response variable can integrate a variety of potential types of response into the Bayesian hierarchical model in a non-parametric manner that does not require an assumption of identical distributions of the response variable.

Another benefit to this analytic approach is that it enables the pre-specification of a response trajectory for specific patients at enrollment into the registry. This should be possible for potential response trajectories for various treatment options within an equipoise set—defined as options where the variance around the patient-specific trajectory forecast overlaps among options.

By defining evidence-based clusters that integrate clinical information, including demographics, tumor anatomic information, histopathology, prior treatment responses, with laboratory parameters including panomic information to create a grouping of patients that appropriately weights all the information available and accounts for the covariate relationships that exist between different types of information, XCELSIOR should be able to assess individual patient treatment hypotheses prospectively against response forecasts. .

##### 8.1.2 EQUIPOISE OPTIONS SET AND RESPONSE MEASUREMENT

XCures acting alone, on behalf of, or with partner or collaborator currently maintains a curated list of treatment options for various advanced and recalcitrant cancers. This includes both regimens and clinical trials. In addition, treatment options suggested during consultation from VTB experts is incorporated with rationale. For example, if a patient with a tumor that is highly expressing a particular protein or gene for which there is a targeted therapy receives a suggested option from the VTB that includes off-label use for the targeted therapy due to the presence of amplification of the drug target, that becomes part of the global option set and can be matched to future patients with similar characteristics.

Matching of potential treatments to patients within clusters is accomplished through the VTB process, in which a cancer biologist at XCures acting alone, on behalf of, or with partner or collaborator curates the patient's case information and assembles a list of possible treatment options based on rationales for review by the VTB. The VTB can refine this list or add other options. Once implemented, the patient cluster model will facilitate this process by quickly identifying options and outcomes for similar patients.

The VTB curates individual treatment option sets with analytic support to evaluate equipoise. Equipoise is defined as an overlap in the calculated treatment outcome variance among options considered in relation to similar patients defined via clustering.

Reporting of treatments will involve a filtered “Leaderboard” based on how various options have performed for different clusters of patients. The following characteristics may be calculated as the leaderboard parameters of treatment options both overall and in relation to specific patient clusters, including:

- Number of times treatment was recommended by the VTB
- Number of times a VTB-recommended treatment was chosen by a patient's doctor
- Number of patients actively on the treatment
- Efficacy signal to date using Bayesian model that can incorporate external information
- Safety/Tolerability signal to date
- Information Score

Each leaderboard item will be scored based on an iterative ranking algorithm for pairwise comparisons, which are commonly used in many fields from sports to the famous PageRank algorithm used by Google Application of a Bayesian algorithm may be employed to incorporate external information. An example of a high-performing implementation can be found in (Negahban et al., 2017). The leaderboard of treatments filtered for existing patient clusters provides a layer of expert human assessment with outcomes data for further decision analysis.

The "Information Score" of a treatment overall or in relation to specific subgroups of patients is important for developing guided search through the universe of possible treatments. This is part of the global cumulative treatment analysis, in which treatments for which a great deal of information exists would be unlikely to add significant information relative to treatments for which little information exists.

As a treatment accumulates more data, the strength of signals increases and may facilitate partitioning or rearrangement of the overall patient cluster model as information about a treatment begins to suggest which patients within a subgroup respond differently. However, as the variance associated with treatment signals declines in response to accumulating data, less well-studied options may better align with patient utility and risk tolerance as lack of information corresponds to a wide variance in potential outcomes.

###### 8.1.3 Patient Utility Analysis

One of the goals of XCELSIOR is to use quantitative methods to incorporate patient preference, including goals and risk-tolerance into the analysis of treatment options. This approach complements the goal of assessing and monitoring global information gain. Information and uncertainty in treatment is typically expressed as variance. Similarly, in economics, including behavioral economics variance equates to risk. This suggests that by incorporating a patient goals and preferences survey using conjoint regression, a utility function expressed mathematically can be incorporated into the scoring of possible treatments and that this scoring method will closely align with information gain within the overall system. The benefit of this approach is that it can be used to help reduce the number of equipoise treatment options by adding alignment (a distance measurement between two functions—the treatment information gain function calculated for a specific patient with the patient's utility function derived from a conjoint analysis survey at registration). This information may be provided to the patient's physician and/or VTB to help them refine their considerations and incorporate patient-specific goals and values. This approach relies on a series of simple paired choice questions to derive a complex function and unlike some quantitative methods reduces the need to either patients or doctors to have high-level numeracy (Politi et al., 2012).

The goal is to help ensure that patients receive treatments that align with their goals. It is hypothesized that risk tolerance is correlated with overall information gain, and this will be an area of analysis within XCELSIOR. A similar approach was implemented by Mohamed et al (ISPOR conference poster) for renal cell carcinoma patients. In that study, the most important goal resulting from conjoint regression of the paired choice options was the prospect of a month extension of PFS was worth a 1% increase in risk of death from a fatal adverse drug effect.

###### 8.1.4 Analytic Signal Detection

General and disease-specific response signals will be detected using traditional query detection methods augmented with both human expertise and machine learning techniques. Signals will be detected by monitoring routine and spontaneous assessments collected from the medical record and augmented with surveys, which may include labs, imaging, patient- and physician-reported questionnaires, and other information such as treatment switching or progression declarations. Signals may be detected from changes in magnitude or rate of change and scored based on direction of the change and correlation with the VTB treatment rationale. Once detected, potential signals will be escalated for human review by a scientist, medical director, or the VTB/vDMC. Signals may be mapped to response criteria, for example RANO. Spontaneous response measures, such as time-to-progression, time-to-treatment switching, and/or changes to anti-cancer therapy regimen will be detected and scored using paired TTP or TTTS and in relation to baseline forecasts.

It should be noted that signal detection in non-linear networked systems, such as the composite responder analysis integrating numerous non-linear assessments into a network of aggregated n-of-1 experiments, benefits from a certain level of "noise" within the network. This phenomenon is called stochastic resonance and is widely used in telecommunications and other networked systems to amplify weak signals in a noisy environment. Such approaches may be considered for enhancing signal detection in XCELSIOR.

###### 8.1.5 Sample Size Considerations

Because this registry is not hypothesis-driven, formal prospective sample-size calculations are not required.

###### 8.1.6 Populations for Analyses

The entire research database will be subject to classification analysis using various machine learning models to build an evolving and logical framework for patient similarity using all available baseline co-variate information. Cluster models may include Bayesian hierarchical models, K Nearest Neighbor, Random Forest models, and Deep Neural Networks. The purpose of population analysis is to determine the relative importance of baseline co-variates in support of individual treatment matching. Each patient will be algorithmically matched to their nearest similar patients for the purposes of identifying treatment and response in similar patients. As new information develops continuously, the cluster algorithm will be recalculated to incorporate new information gain.

##### 8.1.7 Statistical Analyses

Because the symptoms of recalcitrant cancers can be diverse, an integrated responder analysis approach will be used for signal detection. This approach will integrate symptomatic, clinical, laboratory, imaging, or other measures of response into a responder analysis framework. This approach has been used successfully in rare disease studies that are characterized by few patients and diverse symptoms.

Because individuals in XCLESIOR are likely to have few treatment opportunities due to a short expected life-span, the goal of responder analysis is to identify response early, or in the absence of response work with the patient, their physician and the VTB to try the next equipoise option or re-run the VTB process quickly to start an alternative therapy. This approach should enable XCELSIOR to observe a series of n-of-1 treatments within patients. These changes occur naturalistically during the treatment of cancer patients and will be evaluated individually and in an aggregated n-of-1 analysis for safety and effectiveness signal detection. Effectiveness signal detection may be based on Bayesian models that incorporate hierarchical clustering with additive regression trees to both group patients in a continuously learning model, while also implementing a flexible and efficient model layer for estimating individual treatment response prospectively using Bayesian Additive Regression Trees (Logan et al., 2018).

Analyses may be conducted to assess hypotheses for both the safety and/or effectiveness of various treatments along or in combination for different patient subgroups, based on the emergence of data suggesting new hypotheses. VTB members and interested researchers may propose research plans and analyses to XCures acting alone, on behalf of, or with partner or collaborator. Such analyses will be reviewed by XCures acting alone, on behalf of, or with partner or collaborator and may be undertaken in accordance with the publication policy in Section 5.1.7.

##### 8.1.8 Baseline Descriptive Statistics and Annual Reporting

Ongoing aggregated and summary reports using descriptive statistics will be provided for the XCELSIOR registry population. These analysis reports will include demographics, disease characteristics, and prior treatments, as well as other baseline covariates, summaries of treatments, outcomes, and adverse events. Summary reports of data quality, missing data, and general operational information will be made available in aggregate and blinded form to patients, clinicians, and advocacy organizations affiliated with XCures acting alone, on behalf of, or with partner or collaborator or XCELSIOR and may be made public through media or peer-reviewed outlets.

##### 8.1.9 Exploratory and Future Analyses

Potential future research on the XCELSIOR Research Data is expected. By consenting to participate subjects are consenting to the use of de-identified data for future research purposes, without the requirement for additional informed consent. Exploratory and future analyses of Research Data or de-identified data will adhere to the overall objectives of the registry, particularly the improvement of cancer care for participating and future patients.

### 9 SUPPORTING DOCUMENTATION AND CONSIDERATIONS

#### 9.1 Regulatory and Ethical Considerations

##### 9.1.1 Regulatory Considerations

As a patient-centric, non-interventional prospective, observational study, the proposed research program meets the regulatory criteria under 21 CFR 312.2(b)(1) for an exemption from the requirement for the submission and FDA-acceptance, of an IND application. Specifically:

1. The investigation is not intended to be reported to the FDA as a well-controlled study in support of a new indication for the use of an FDA-regulated product, nor intended to be used to support any other significant change in the labeling of an FDA-regulated product. Data collected in XCELSIOR will be captured in accordance with 21 CFR 11 and is intended to be "regulatory grade" real-world evidence. Such information may be used by the FDA under the authority in the 21st Century Cures Act, however XCELSIOR is not being conducted on behalf of drug or device manufacturers for the purpose of assessing their products in a well-controlled study.

2. The investigation is not intended to support a significant change in the advertising for an FDA-regulated product. The purpose of this study is to use machine learning to identify factors which may be predictive of treatment response at the level of individual patients and disseminate that information to patients and physicians.

3. The investigation does not involve a route of administration or dosage level or use in a patient population or other factor that significantly increases the risks (or decreases the acceptability of the risks) associated with the use of an FDA regulated product. Specifically, this is a data-collection registry, treatment of patients will continue at the discretion of their physician. XCures acting alone, on behalf of, or with partner or collaborator, through the VTB, may provide information to the patient and their physician about molecularly targeted therapeutic options and/or clinical trials for which the patient may be eligible. Decisions about treatment or participation in clinical trials will be independent of XCures acting alone, on behalf of, or with partner or collaborator.

4. The investigation is subject to prior approval by the Genetic Alliance IRB, based in, which operates in compliance with the FDA regulations including 21 CFR Parts 50 and 56 and is registered with OHRP as IRB Organization number IORG0003358.

5. Neither the participants in this clinical investigation, nor their insurance providers, will be charged for the procedures associated with participation in the study. Patients in this registry will be treated according to their institution's normal standards of practice and at their oncologist's discretion.

The technology platform may be considered a Personal Health Record, due to the collection and storage of the following information:

- Contact information for the patient and his or her family members
- A list of providers involved in the patient's care and their contact information
- Information about the patient's diagnosis, medications, medical history

- Lab and test results
- Family medical history

Based on this, XCures acting alone, on behalf of, or with partner or collaborator will comply with the FTC Health Breach Notification Rule (16 CFR 318) in the event of an unauthorized breach of confidentiality of patient information. Such an incident would be reported to the Genetic Alliance IRB in accordance with the procedures for UP reporting. Note that XCures acting alone, on behalf of, or with partner or collaborator encourages patient exchanges of information to facilitate discussion among patients with cancer. Such disclosures are made by patients and are not considered unauthorized disclosure.

This registry study will comply with sections 402(i) and 402(j) of the Public Health Service Act (PHS Act) (42 U.S.C. §§ 282(i) and (j)), which was amended by Title VIII of the Food and Drug Administration Amendments Act of 2007 (FDAAA) (Public Law No. 110-85, 121 Stat. 904) by registering and reporting results in the Clinicaltrials.gov database.

##### 9.1.2 Confidentiality, Privacy, and Data Security

Participant confidentiality and privacy is strictly held in trust by XCures acting alone, on behalf of, or with partner or collaborator, their staff, VTB members and affiliates. At the same time, XCures acting alone, on behalf of, or with partner or collaborator is a patient-directed research and advocacy service that encourages the sharing of information between and among patients to facilitate better cancer care. XCures acting alone, on behalf of, or with partner or collaborator maintains the confidentiality of records and that confidentiality extends to cover test results of biological samples and genetic tests in addition to the clinical information relating to participants. Therefore, the study data will be stored in secure electronic format and personally identifiable information will not be disclosed without the express permission of the participant, however authorized representatives of XCures acting alone, on behalf of, or with partner or collaborator and representatives of the Genetic Alliance IRB, as well as regulatory agencies may inspect records in certain circumstances without prior approval.

Research data, which is for purposes of statistical analysis and scientific reporting, will be transmitted to and stored by XCures acting alone, on behalf of, or with partner or collaborator in a research study database within the REDCap EDC system. REDCap is a secure, cloud-based system that is HIPAA, FISMA, GDPR, Annex 11, and 21 CFR 11 compliance system for the collection and storage of clinical research and survey information. Services Data related to the provision of XCures acting alone, on behalf of, or with partner or collaborator' patient advocacy and support services will not be considered Research Data under this protocol and will be maintained separately. Services Data includes the participant's contact or identifying information as well as other information within a system considered a Personal Health Record, which is intended to assist with the provision of services directly to patients. For both Research Data and Services Data, sensitive personal information will be stored in a secure database using a role and permission-based system for access. Sensitive data elements, including PHI or other sensitive or identifiable information will be stored in encrypted format. Research Data will identify participants only by a unique study identification number. The decode list for this information will be maintained securely and not disclosed to recipients of the Research Data. Research Data may be disclosed outside of XCures acting alone, on behalf of, or with partner or collaborator and their agents and affiliates for the purposes of research and education, or to further the development of treatments for cancer patients.

##### 9.1.3 Schema and Overview of Services Data Versus Research Data in XCELSIOR

Figure 7 below provides a schematic of the overall relationship between Services Data and Research Data generated and maintained by XCures acting alone, on behalf of, or with partner or collaborator.

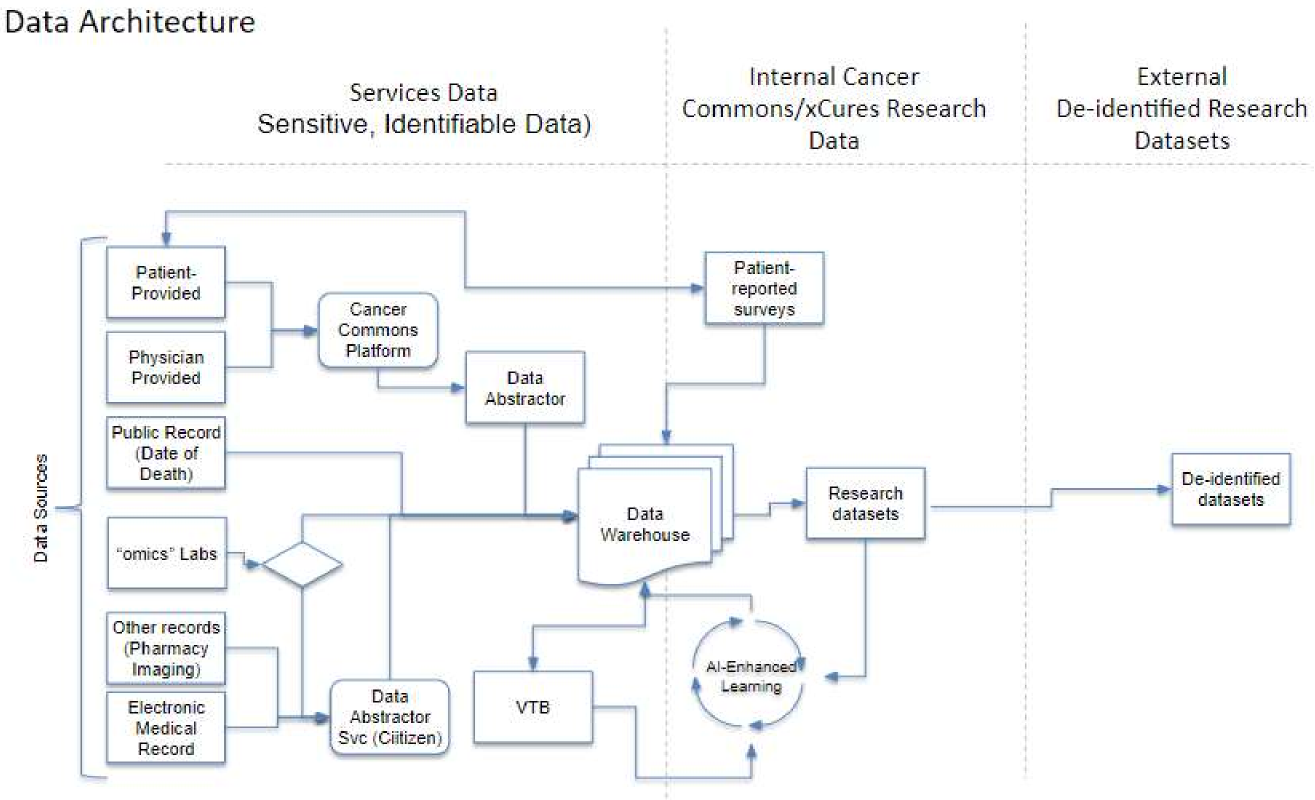

**Figure 7.** Overview of the types of information that may be considered Services Data, Research Data, or de-identified data subject to future use as stored data.

| | Services Data | Internal Cancer Commons/xCures Research Data | External De-identified Research Database |
|---|---|---|---|
| Data Use | Patient contact for options and access information, including VTB | Cluster analysis for option set evaluation and outcomes analysis, possible reporting to FDA for adverse events | De-identified datasets for laboratory test development (e.g. non-CLIA tests), academic research for publication, safety, Commercial use, public health, patient advocacy |
| Data Type | Full PHI, financial information for access support services, unredacted medical record data, patient reported questionnaires, photos and statements | Limited PHI held in encrypted and permission-controlled area within REDCap to facilitate Patient-reported outcomes collection, abstracted and coded medical data, redacted/de-identified patient source documents | Full-de-identified data exports from REDCap Cloud. Unique identifier number to enable decode only by Cancer Commons/xCures for linkage of data sets. |
| Possible Systems | Hubspot, Cancer Commons VTB Platform, REDCap Cloud, Github, Anaconda | REDCap Cloud, Anaconda, SAS | CDISC XML, SAS transport files, etc. |

**Table 1.** Summary of data types, uses, and systems.

##### 9.1.4 Future Use of Stored Data

Data collected for this study will be analyzed and stored by XCures acting alone, on behalf of, or with partner or collaborator and their designated data/technology provider(s). This study is intended to operate as a perpetual protocol, but in the event the study is terminated for any reason, the de-identified, archived Research Data will be retained for future use by other researchers possibly including those outside of the study.

##### 9.1.5 Publication and Data Sharing Policy

This study will be conducted in accordance with the ICMJE publication and data sharing policies. XCures acting alone, on behalf of, or with partner or collaborator and their designees are responsible for submitting manuscripts for publication. Every attempt will be made to publish results in a suitable, peer-reviewed journal.

This trial will be registered at ClinicalTrials.gov and/or AHRQ's RoPR is a database of registry specific information intended to promote collaboration, reduce redundancy, and improve transparency. Results information from this trial will be submitted to ClinicalTrials.gov either directly or through a reference to the peer-reviewed manuscript after publication. Information may be shared with patients, physicians, and regulatory authorities consistent with and in support of the overall mission of XCures acting alone, on behalf of, or with partner or collaborator as a patient advocacy organization.

##### 9.1.6 Conflict of Interest Policy

The independence of this study from any actual or perceived influence, is critical. Therefore, any actual conflict of interest of persons who have a role in the design, conduct, analysis, publication, or any aspect of this trial will be disclosed and managed. Furthermore, persons who have a perceived conflict of interest will be required to have such conflicts managed in a way that is appropriate to their participation in the design and conduct of this trial. The study leadership at XCures acting alone, on behalf of, or with partner or collaborator in conjunction with xCures has established policies and procedures for all study group members to disclose all conflicts of interest and will establish a mechanism for the management of all reported dualities of interest.

###### 9.1.7 Ethical Considerations

The XCELSIOR research protocol and consent form, along with other patient-facing materials will be provided to the Genetic Alliance IRB for review and approval prior to implementation.

All patients must provide informed consent. The process will be conducted electronically using the secure e-consent process in REDCap EDC. Documentation of the consent will be maintained. Patients will receive an electronic copy of their consent. Further, patients may discontinue participation at any time in accordance with the procedures outlined in the consent form. All data collected up to the point of withdrawal will remain in the Research Database and may be included in de-identified data for future analyses.

Children with late-stage or recalcitrant cancers may participate in the XCELSIOR registry. For qualifying children, consent will be obtained from the parent or legally-authorized representative.

Patients in XCELSIOR may be experiencing diminished mental capacity due to the presence of CNS tumors or brain metastases. Patients with diminished mental capacity due to their disease may participate provided that a legally-authorized representative provides informed consent.

##### 9.2 Study Oversight and Other Considerations

###### 9.2.1 Financial Considerations

XCures acting alone, on behalf of, or with partner or collaborator is responsible for the costs associated with implementing and maintaining the registry. Subjects and their providers will not incur costs for participation. Members of the VTB will be compensated for their time in accordance with their institutional policies and the XCELSIOR conflict-of-interest policy. Partner and collaborator organizations may receive donations from various sources to support their services, including work with xCures and XCELSIOR. 501(c)3 non-profit partners and collaborators, such as Cancer Commons and the Musella Foundation, make annual financial disclosures in accordance with on a publicly available IRS Form 990.

###### 9.2.2 Study Discontinuation and Closure

This study may be suspended or prematurely terminated at any time for any reason, at the discretion of XCures acting alone, on behalf of, or with partner or collaborator. Written notification, documenting the reason for study suspension or termination, will be provided by the suspending or terminating party to study participants, their legally-authorized representative (if applicable), and their physician, as well as participating members of the VTB, as well funding agencies, the IRB, and regulatory authorities, if required. If the study is prematurely terminated or suspended, XCures acting alone, on behalf of, or with partner or collaborator will promptly inform stakeholders and will provide the reason(s) for the termination or suspension.

##### 9.2.3 Quality Assurance and Quality Control

XCures acting alone, on behalf of, or with partner or collaborator will perform internal quality management of study conduct, including collection, handling and analysis of data. A quality management system will be implemented and written Standard Operating Procedures (SOPs) will be followed.

Quality control (QC) procedures will be implemented within the data collection and storage systems with quality control procedures. Any missing data or data anomalies will be investigated.

Noncompliance events that meet the IRB's reporting requirements must be reported to the IRB office within 10 working days of XCures acting alone, on behalf of, or with partner or collaborator becoming aware of the event.

##### 9.2.4 Key Roles and Study Governance

| | |
|---|---|
| San Francisco Office: | 1901 Harrison Street, Suite 1100, Oakland, CA 94612 |
| North Carolina Office: | 4509 Creedmoor Rd. Suite 201, Raleigh, NC 27612 |
| Fax: (650) 600-0095 | For Fedex/overnights requiring phone numbers use: (984) 789-5811 |

| **VP, Clinical Development and Principal Investigator** | **Chief Medical Officer (Consulting)** | **VP, Clinical Operations, Head Patient Operations and Patient Access** |
|---|---|---|
| Mark Shapiro, MA, MBA | Santosh Kesari, MD, PhD | Bryan Federowicz |
| Mobile: (919) 306-3675<br>Fax: (919) 883-4400 | Mobile: (617) 233-6202 | Office Main: (650) 530-3636<br>Mobile: (919) 283- 1899<br>Fax: (650) 289-4041 |
| | | |

| **Director of Research** | **Lead Data Scientist** | **CTO** |
|---|---|---|
| Jeff Schrager, PhD | Asher Wasserman, PhD | Glenn Kramer, PhD |
| Office: (650) 530-3636 | Mobile: (732) 567-7875 | Mobile: (415) 298-3057 |
| | | |

#### 9.3 Abbreviations

| | |
|---|---|
| AACR | American Association of Cancer Research |
| AE | Adverse Event |
| AI | Artificial Intelligence |
| ASCO | American Society of Clinical Oncology |
| CFR | Code of Federal Regulations |
| CLIA | Clinical Laboratory Improvement Amendments |
| CR | Complete Response |
| CRF | Case Report Form |
| CTCAE | Common Terminology for Cancer Adverse Events |
| CTEP | Cancer Therapy Evaluation Program |
| DFS | Disease Free Survival |
| DICOM | Digital Imaging and Communications in Medicine |
| DMC | Data Monitoring Committee |
| DR | Durable Response |
| DRE | Disease-Related Event |
| ECG | Electrocardiogram |
| eCRF | Electronic Case Report Forms |
| EMR | Electronic Medical Record |
| FDA | Food and Drug Administration |
| FDAAA | Food and Drug Administration Amendments Act of 2007 |
| FISMA | Federal Information Security Management Act |
| GBM | Glioblastoma multiforme |
| GCP | Good Clinical Practice |
| GCTA | Global Cumulative Treatment Analysis |
| HIPAA | Health Insurance Portability and Accountability Act |
| IB | Investigators' Brochure |
| ICH | International Conference on Harmonisation |
| ICMJE | International Committee of Medical Journal Editors |
| IDE | Investigational Device Exemption |
| IND | Investigational New Drug Application |
| IRB | Institutional Review Board |
| ISPOR | International Society for Patient Outcomes Research |
| MedDRA | Medical Dictionary for Regulatory Activities |
| ML | Machine Learning |
| MRN | Medical Record Number |
| MTB | Molecular tumor board |
| NCI | National Cancer Institute |
| NCT | National Clinical Trial |

| | |
|---|---|
| NGS | Next-generation sequencing |
| NIH | National Institutes of Health |
| NLM | National Library of Medicine |
| NP | non-parametric |
| OS | Overall Survival |
| PCORI | Patient Centered Outcomes Research Initiative |
| PFS | Progression Free Survival |
| PHI | Private Health Information |
| PI | Principal Investigator |
| QA | Quality Assurance |
| QC | Quality Control |
| RECIST | Response Evaluation Criteria in Solid Tumors |
| RR | Response Rate |
| RWE | Real-world evidence |
| SAE | Serious Adverse Event |
| SAP | Statistical Analysis Plan |
| SDM | Shared decision making |
| SOP | Standard Operating Procedure |
| TTF | Time-to-treatment failure |
| TTP | Time-to-progression |
| UP | Unanticipated Problem |
| US | United States |
| USC | United States Code |
| vDMC | Virtual Data Monitoring Committee |
| VTB | Virtual Tumor Board |
| WHO | World Health Organization |
| WHODrug | WHO Drug Dictionary |

Abernethy A, Arunachalam A, Burke T, McKay C, Cao X, Sorg R, et al. (2017) Real-world first-line treatment and overall survival in non-small cell lung cancer without known EGFR mutations or ALK rearrangements in US community oncology setting. PLoS ONE 2017;12(6): e0178420.

Alharbi M, Mobark N, Mobark N, Almubarak L, Aljelaify R, Alsaeed M Almutairi A, Alqubaishi F, Hussain E, Balbaid A, Marie A, Alsubaie L, Alshieban S, Altassan N, Ramkissoon S, Abedalthagafi M. Durable Response to Nivolumab in a Pediatric Patient with Refractory Glioblastoma and Constitutional Biallelic Mismatch Repair Deficiency. The Oncologist 2018; 23:1–6.

Abrahamyan L, Diamond I, Johnson S, Feldman B. A New Toolkit for Conducting Clinical Trials in Rare Disorders. J Popul Ther Clin Pharmacol 2014;21(1): e66-e78.

American Association for Cancer Research (AACR); Participation in Cancer Clinical Trials; [Accessed Nov 2018]. *https://blog.aacr.org/broadening-clinical-trial-participation/*

Berry D. Bayesian Clinical Trials. Nature Reviews 2006; 5:27-36.

Brennan C, Momota H, Hambardzumyan D, Ozawa T, Tandon A, et al. Glioblastoma Subclasses Can Be Defined by Activity among Signal Transduction Pathways and Associated Genomic Alterations. PLoS ONE 2009;4(11): e7752.

Bristol-Myers Squibb Announces Results from CheckMate -143, a Phase 3 Study of Opdivo (nivolumab) in Patients with Glioblastoma Multiforme. Business Wire, April 3, 2017; [Accessed Nov 2018]. Available from: https://www.businesswire.com/news/home/20170403005964/en/Bristol-Myers-Squibb-Announces-Results-CheckMate--143-Phase

Collins F and Varmus H. A New Initiative on Precision Medicine. NEJM 2015 Feb;372(9):793-5.

Dana-Farber Cancer Institute. A Brain Tumor Kept at Bay by Immunotherapy. May 29, 2017; [Accessed Nov 2018]. Available from: https://blog.dana-farber.org/insight/2017/05/a-brain-tumor-kept-at-bay-by-immunotherapy/

Duan N, Kravitz R, Schmid C. Single-patient N-of-1 Trials: A pragmatic clinical decision methodology for patient-centered comparative effectiveness research. J. Clin. Epidemiol 2013; 66(8): S21-8.

Gliklich RE, Dreyer NA, eds. Registries for Evaluating Patient Outcomes: A User's Guide. 2nd ed. (Prepared by Outcome DEcIDE Center [Outcome Sciences, Inc. d/b/a Outcome] under Contract No. HHSA29020050035I TO3.) AHRQ Publication No.10-EHC049. Rockville, MD: Agency for Healthcare Research and Quality. September 2010

Glass H, Glass L, Glass J, Tran P. Are Phase III Clinical Trials Really Consistently Behind Schedule? Applied Clinical Trials 2016 Jun.

Gyawali B, Parsad S, Feinberg B, Nabhan C. Real-World Evidence and Randomized Studies in the Precision Oncology Era: The Right Balance. *JCO Precision Oncology* 2017 Oct; 1-4.

Hillen M, Medendorp N, Daams J, Smets E. Patient-driven second opinions in oncology: A systematic review. The Oncologist 2017; 22:1197-22.

Hirsch I, Martinez J, Dorsey E, Finken G, Fleming A, Gropp C, Home P, Kaufer D, Papapetropoulos S. Incorporating Site-less Clinical Trials into Drug Development: A Framework for Action. Clin Ther. 2017 May;39(5):1064-1076.

Ioannidis J. Why most published research findings are false. PLoS Med 2005;2(8): e124.

Ioannidis J. Limits to forecasting in personalized medicine: An overview. International Journal of Forecasting 2009; 25:773–83.

Jiang F, Jiang Y, Zhi H, Li H, Ma S, Wang Y, Dong Q, Shen H, Wang Y. Artificial intelligence in healthcare: part, present and future. Stroke and Vascular Neurology 2017; 230-243.

Kahneman D and Thaler R. Utility Maximization and Experienced Utility. J Econ Perspect 2006 Winter;20(1):221–34.

Khozin, S. Keynote at the 13th Annual Personalized Medicine Conference. 11/17/2017; [Accessed Nov 2018]. Available from: https://www.seankhozin.com/journal/category/all

Kulis B and Jordan M. Revisiting k-means: New Algorithms via Nonparametrics. Proceedings of the 29th International Conference on Machine Learning, Edinburgh, Scotland, UK, 2012.

Klement, Arkun, Valik, Roffidal, Hashemi, Klement, Carmassi, Rietman, Slaby, Mazanek, Mudry, Kovacs, Kiss, Norga, Konstantinov, André, Slavc, van Den Berg, Kolenova, Kren, Tuma, Skotakova, and Sterba. Future paradigms for precision oncology. Oncotarget 2016; 7(29):46813-31.

Kurzrock R and Giles F. Precision oncology for patients with advanced cancer: the challenges of malignant snowflakes. Cell Cycle 2015; 14(14):2219-21.

Leiby B, Sammel M, Ten have T, Lynch K. Identification of multivariate responders and non-responders by using Bayesian growth curve latent class models. J Royal Stat Soc Appl. Statist 2009;58(4):505–24.

Lillie E, Patay B, Diamant J, Issell B, Topol E, Schork N. The n-of-1 clinical trial: the ultimate strategy for individualizing medicine? Per Med 2011; 8(2):161-73.

Lim L, Pullenayegum E, Moineddin R, Gladman D, Silverman D, Feldman B. Methods for analyzing observational longitudinal prognosis studies for rheumatic diseases: a review & worked example using a clinic-based cohort of juvenile dermatomyositis patients. Pediatric Rheumatology 2017; 15:18

Lim W, Arnold D, Bachanova V, Haspel R, Rosovsky R, Shustov A, Crowther M. Evidence-Based Guidelines—An Introduction. Hematology 2008; 1:26-30.

Logan B, Sparapani R, McCulloch R, Laud P. Decision making and uncertainty quantification for individualized treatments using Bayesian Additive Regression Trees. Stat Methods Med Res 2017; 1-22.

Markman M, Kramer K, Alvarez, R, Weiss G, Ahn E, and Daneker G. Evaluating the Utility of a “N-of-1” Precision Cancer Medicine Strategy: The Case for ‘Time-to-Subsequent-Disease Progression.’ Oncology 2016; 91:299-301.

Mick R, Crowley J, Carroll R. Phase II Clinical Trial Design for Noncytotoxic Anticancer Agents for which Time to Disease Progression is the Primary Endpoint

Mohamed A, Yang J, Hauber A, Liu Z, Wong M, Rogiero J, Garay C. Patient Benefit-Risk Preferences for Advanced Renal Cell Carcinoma Treatments: Results from a Conjoint Analysis Study. ISPOR 17th Annual International Meeting, Washington, DC, United States 2006.

Murphy, T. Cancer patients are twice as likely to declare bankruptcy. Chicago Tribune, May 22, 2018; [Accessed Nov 2018]. Available from: www.chicagotribune.com/news/nationworld/ct-cancer-treatment-debt-20180522-story.html

National Library of Medicine; Definition of Precision Medicine [Internet]. [Accessed Nov 2018]. Available from: https://ghr.nlm.nih.gov/primer/precisionmedicine/definition

Neelon B, O'Malley A, Normand S. A Bayesian Two-Part Latent Class Model for Longitudinal Medical Expenditure Data: Assessing the Impact of Mental Health and Substance Abuse Parity. Biometrics 2011;61(1):280-9.

Negahban et al. Rank Centrality: Ranking from Pairwise Comparisons. Operations Research 2017;65(1):266-287

Parker B, Schwaederle M, Scur M. Breast Cancer Experience of the Molecular Tumor Board at Univ of Cal, San Diego Moores Cancer Center. J Oncol Practice 2015; 11:442-449.

Politi M, Lewis C, Frosch D. Supporting Shared Decisions When Clinical Evidence Is Low. Medical Care Research and Review 2013;70(1):113S-128S.

Radovich M, Kiel P, Nance S. Clinical benefit of a precision medicine-based approach for guiding treatment of refractory cancers. Oncotarget 2016; 7:56491-56500.

Renfro L and Sargent D. Statistical controversies in clinical research: basket trials, umbrella trials, and other master protocols: a review and examples. Annals of Oncology 2017; 28: 34–43.

Schmid V, Whitcher B, Padhani A, Taylor N, Yang G. A Bayesian Hierarchical Model for the Analysis of a Longitudinal Dynamic Contrast-Enhanced MRI Oncology Study. Magnetic Resonance in Medicine 2009; 61:163-74.

Schork N. Time for one-person trials. Nature 2015 Apr 30;520(7549):609-11

Sherman R, Anderson S, Dal Pan G, Gray G, Gross T, Hunter N, LaVange L, Marinac-Dabic D, Marks P, Robb M, Shuren J, Temple R, Woodcock J, Yue L, Califf R. Real-World Evidence: What Is It and What Can It Tell Us? NEJM 2016 Dec; 375(23):2293-7.

Shrager J and Tenenbaum J. Rapid learning for precision oncology. Nat Rev Clin Oncol 2014 Feb; 11(2): 109-18.

Silvestris N, Ciliberto G, De Paoli P, Apolone G, Lavitrano M, Pierotti M, Stanta G et al. Liquid dynamic medicine and N-of-1 clinical trials: a change of perspective in oncology research. J Experimental & Clinical Cancer Res 2017; 36(128):1-5.

Sireci A, Aggarwal V, Turk A, Gindin T, Mansukhani M, Hsiao S. Clinical Genomic Profiling of a Diverse Array of Oncology Specimens at a Large Academic Cancer Center Identification of Targetable Variants and Experience with Reimbursement. J Mol Diagn 2017; 19:277-87.

Strause L. Patient-First Approach to Improve Oncology Clinical Trials. Applied Clinical Trials 2013 Mar; 26-31.

Subbiah V, Chuang H, Dhiraj G, Kairemo K. Defining Clinical Response Criteria and Early Response Criteria for Precision Oncology: Current State-of-the-art and Future Perspectives. Diagnostics 2017;7(10):1-17.

Tsimberidou, Hong, Ye, Cartwright, Wheler, Falchook, Naing, Fu, Piha-Paul, Janku, Meric-Bernstam, Hwu, Kee, Kies, Broaddus, Mendelsohn, Hess, Kurzrock. Initiative for Molecular Profiling and Advanced Cancer Therapy (IMPACT): An MD Anderson Precision Medicine Study. JCO Precis Oncol 2017; 1-24.

Van Allen, Wagle, Stojanov, Kryukov, Marlow, Jane-Valbuena, Friedrich, Kiezun, Carter, McKenna, Sivachenko, Rosenberg, Huang, Voet, Lawrence, Lichtenstein, Gentry, Lander, Fostel, Farlow, Barbie, Ghandi, MacOnaill, Gray, Joffe, Janne, Garber, Bariel, Lindeman, Rollins, Kantoff, Fisher, Getz, Garraway. Whole-exome sequencing and clinical interpretation of FFPE tumor samples to guide precision cancer medicine. Nat Med. 2014 June; 20(6): 682–688.

Vieira R, McDonald S, Araujo-Soares, Sniehotta F, Henderson R. Dynamic modelling of n-of-1 data: powerful and flexible data analytics applied to individualized studies. Health Psychology Review 2017; 11(3):222-34.

Von Hoff D, Stephenson K, Rosen P. A Pilot Study Using Molecular Profiling of Patients' Tumors to Find Potential Targets and Select Treatments for their Refractory Cancers. J. Clin. Oncol 2010; 28:4877-4883.
Rothwell P. External Validity of Randomized Controlled Trials: 'To whom do the results of this trial apply?' Lancet 2005; 365(9453): 82-93.

Wang Q, Hu B, Hu X, Kim H, Squatrito M, Scarapace L, de Carvalho A, Lyu S, Li P, Li Y, Barthel F, Cho, H, Lin Y et al. Tumor Evolution of Glioma-Intrinsic Gene Expression Subtypes Associates with Immunological Changes in the Microenvironment. Cancer Cell 2017; 32:42–56.

Wang J, Cazzato E, Ladewig E, Frattini V, Rosenbloom D, Zairis S, Abate F, Liu, F, Elliott O, Shin Y, Lee Y, Lee I, Park W, Eoli W, Blumberg A, Lasorella A, Nam D, Finocchiaro G, Iavarone G, Rabadan R. Clonal Evolution of Glioblastoma under Therapy. Nat Genet. 2016 Jul; 48(7): 768–776.

Wheler J, Lee J, and Kurzrock R. Unique Molecular Landscapes in Cancer: Implications for Individualized, Curated Drug Combinations. Cancer Res 2014 Dec; 74(24):7181-4.

Weinreich S, Vrinten C, Kuijpers M, Lipka A, Schimmel K, van Zwet E, Gispen-de Wied C, Hekster Y, Verschuuren J, Cornel M. Aggregated N-of-1 trials for unlicensed medicines for small populations: an assessment of a trial with ephedrine for myasthenia gravis. Orphanet Journal of Rare Diseases 2017; 12:88.

Woodcock J and LaVange L. Master protocols to Study Multiple Therapies, Multiple Diseases, or Both. NEJM 2017; 377(1):62-70.

Zhu X, McDowell M, Newman W, Mason G, Greene S, Tamber M Severe cerebral edema following nivolumab treatment for pediatric glioblastoma: case report. J Neurosurg Pediatr. 2017 Feb;19(2):249-253.

Zucker D, Ruthazer R, Schmid C. Individual (N-of-1) trials can be combined to give population comparative treatment effect estimates: methodologic considerations. J Clin Epidemiol 2010; 63:1312-1323.

https://ctep.cancer.gov/protocolDevelopment/codes_values.htm#disease

https://ctep.cancer.gov/protocolDevelopment/electronic_applications/ctc.htm

US FDA. Guidance for Industry: Clinical Trial Endpoints for the Approval of Cancer Drugs and Biologics. U.S. Department of Health and Human Services. Food and Drug Administration. Center for Drug Evaluation and Research (CDER). Center for Biologics Evaluation and Research (CBER). May 2007

US FDA. Guidance for Industry: Patient-Reported outcomes

US FDA. Guidance for Industry: Rare Molecular Subtypes

#### APPENDIX 1: INFORMED CONSENT AND ASSENT TEXT

##### XCELSIOR Study Informed Consent

xCures helps provide information through research. xCures ("the study team") is conducting a research study called XCELSIOR (xCures/Cancer Commons Enhanced Learning Treatment Selection and Analysis with Outcomes Research Study). The study team is made up of xCures, POETIC (The Pediatric Oncology Experimental Therapeutics Investigators Consortium, based at Stanford University) and several non-profit partners, including the Musella Foundation for Brain Tumor Education and Research and Cancer Commons.

This study will try to figure out which treatments are best for individual cancer patients. The study involves thousands of cancer patients. If you have cancer or are concerned that you may have cancer and currently live in or are receiving treatment in the United States or US territories, you can participate. If you are the parent of a minor child with cancer or are the legally authorized representative of someone with cancer, you can sign up and they can participate.

When you sign up for services, such as entering a Compassionate Use program managed by xCures, the treatment navigation service offered by Cancer Commons, or another support program offered by one of our partner organizations, you agree to provide your information for review by our scientists and members of our Tumor Board and authorize us to receive that information from you or your doctor. A Tumor Boards a group of experts who will provide information to you and your doctor about possible treatments. By being in the study, the study team will work with our partners to analyze what treatment you and your doctor choose and track how well it works. To see how you are doing, members of the study team may send you or your doctor questions about how you're doing and review changes in your medical information. You may also communicate with us directly to share information that you think is important. This information helps researchers predict which treatments will work best for cancer patients. By sharing your information, you help others, but you also get access to information from other patients that could help you and your doctor.

Being in the study should not put you at risk, but because we collect your information there is a risk of breach of confidentiality. To reduce this risk, your research information is stored in a secure and encrypted database. Information may be shared with other researchers in the future, but your identifying information will not be shared or disclosed outside of study team without your permission.

If you decide to enter hospice care and/or no longer want us to contact you, you or a caregiver can notify us by phone or email. If you change your mind about being in the study, you can stop participating at any time by notifying us in writing. If you stop, we will no longer contact you about the study, but we will keep the information that has been collected about you for research purposes.

If you have questions about the study, you can ask a member of our study team. We have experts who help patients with different types of cancer.  To talk to someone by phone call (650) 530-3636 or by. For questions about your rights as a participant in research, you can contact the Genetic Alliance IRB at 202.966.5557 or by email at. The Genetic Alliance IRB is an independent organization that oversees the research that we conduct.

By entering your information, clicking "I Agree" below, and signing, you are agreeing to the electronic consent form and to get important information about this study online. This process takes the place of traditional paper forms.

xCures and our partners are responsible for the study and for giving you written notices, forms, and other information about your participation in the study. That information is already in this consent document (collectively, the "required information"). By electronically signing this document, you are confirming that you agree:

(1) to receive online the required information, notices and other disclosures related to taking part in this research study;

(2) to continue to receive online any records, documents or other notices regarding the study until and unless you change your mind and withdraw your agreement by notifying the study team in writing; and

(3) if you decide to volunteer for the study, to use an electronic signature in place of a handwritten signature to sign the form agreeing to participate in the study. Thus, by entering your information and clicking on the "I agree" button, you confirm and agree to the above terms and conditions and agree that your electronic consent is the same as signing a paper consent form.

For parents of minor children aged 7 or older, please explain the study to your child.  The following information can be read to them aloud to help.

*A research study is when doctors collect information to learn more about something. A company called xCures is doing a research study to learn more about children with cancer.  They are working together with Cancer Commons and other groups that care about helping kids with cancer. They hope to help find better treatments for cancer. After I tell you about it, I will ask if you'd like to be in this study or not.*

*The study involves cancer experts collecting your information to see how you do on your treatments. You do not have to do anything extra to be in the study. The study team would like permission to look at your medical records. The study team will not ask you to have any testing done just for this study. There is a small chance that someone who is not working on the study could see your information and use it for some other purpose. The study team will do what they can to keep this from happening. There will be hundreds of other children with cancer in the study.*

*You don't have to be in the study if you don't want to. You can be in the study and change your mind later.*

Parents or legally authorized representative, please enter your name and check the box indicating that you have reviewed the study with the child and that they have verbally assented to be in the study.

**Agree to Participate?** *Make one selection*

I have read the consent document and I wish to participate in the study

I have read the consent document and I DO NOT wish to participate in the study

I am the legally authorized representative for the participant listed below and have reviewed the study with them and they have verbally assented to be in the study

**Participant First Name:** ____________________ **Participant Last Name:** ___________________

**Legally Authorized Representative First Name** (*if applicable*): _______________________

**Legally Authorized Representative Last Name** (*if applicable*): _______________________

**Signature of Participant or Legally Authorized Representative:** **Date of Signature:**

_______________________________________________ ______________

***If signed by a legally authorized representative, a description of the representative's authority to act is as follows:**

- ☐ **Parent**
- ☐ **Legal Guardian**
- ☐ **Healthcare Power of Attorney**
- ☐ **Administrator**
- ☐ **Executor of Estate**
- ☐ **Next of kin**
- ☐ **Beneficiary**

#### APPENDIX 2: EXAMPLES OF FUNCTIONAL, QUALITY-OF-LIFE, AND OTHER RESPONSE MEASURES TO BE COLLECTED

ECOG and Karnofsky Performance Status

| ECOG PERFORMANCE STATUS | KARNOFSKY PERFORMANCE STATUS |
| --- | --- |
| 0—Fully active, able to carry on all pre-disease performance without restriction | 100—Normal, no complaints; no evidence of disease<br><br>90—Able to carry on normal activity; minor signs or symptoms of disease |
| 1—Restricted in physically strenuous activity but ambulatory and able to carry out work of a light or sedentary nature, e.g., light house work, office work | 80—Normal activity with effort, some signs or symptoms of disease<br><br>70—Cares for self but unable to carry on normal activity or to do active work |
| 2—Ambulatory and capable of all selfcare but unable to carry out any work activities; up and about more than 50% of waking hours | 60—Requires occasional assistance but is able to care for most of personal needs<br><br>50—Requires considerable assistance and frequent medical care |
| 3—Capable of only limited selfcare; confined to bed or chair more than 50% of waking hours | 40—Disabled; requires special care and assistance<br><br>30—Severely disabled; hospitalization is indicated although death not imminent |
| 4—Completely disabled; cannot carry on any selfcare; totally confined to bed or chair | 20—Very ill; hospitalization and active supportive care necessary<br><br>10—Moribund |
| 5—Dead | 0—Dead |

Reference: *https://ecog-acrin.org/resources/ecog-performance-status* Accessed 26 Oct 2018

### M. D. Anderson Symptom Inventory - Brain Tumor (MDASI - BT)

**Part I. How severe are your symptoms?**

**People with cancer frequently have symptoms that are caused by their disease or by their treatment. We ask you to rate how severe the following symptoms have been in the last 24 hours. Please fill in the circle below from 0 (symptom has not been present) to 10 (the symptom was as bad as you can imagine it could be) for each item.**

| | Not Present 0 | 1 | 2 | 3 | 4 | 5 | 6 | 7 | 8 | 9 | As Bad As You Can Imagine 10 |
|---|---|---|---|---|---|---|---|---|---|---|---|
| 1. Your **pain** at its WORST? | ○ | ○ | ○ | ○ | ○ | ○ | ○ | ○ | ○ | ○ | ○ |
| 2. Your **fatigue (tiredness)** at its WORST? | ○ | ○ | ○ | ○ | ○ | ○ | ○ | ○ | ○ | ○ | ○ |
| 3. Your **nausea** at its WORST? | ○ | ○ | ○ | ○ | ○ | ○ | ○ | ○ | ○ | ○ | ○ |
| 4. Your **disturbed sleep** at its WORST? | ○ | ○ | ○ | ○ | ○ | ○ | ○ | ○ | ○ | ○ | ○ |
| 5. Your feeling of being **distressed (upset)** at its WORST? | ○ | ○ | ○ | ○ | ○ | ○ | ○ | ○ | ○ | ○ | ○ |
| 6. Your **shortness of breath** at its WORST? | ○ | ○ | ○ | ○ | ○ | ○ | ○ | ○ | ○ | ○ | ○ |
| 7. Your problem with **remembering things** at its WORST? | ○ | ○ | ○ | ○ | ○ | ○ | ○ | ○ | ○ | ○ | ○ |
| 8. Your problem with **lack of ap[illegible]** at its WORST? | ○ | ○ | ○ | ○ | ○ | ○ | ○ | ○ | ○ | ○ | ○ |
| 9. Your feeling **drowsy (sleepy)** at its WORST? | ○ | ○ | ○ | ○ | ○ | ○ | ○ | ○ | ○ | ○ | ○ |
| 10. Your having a **dry mouth** at its WORST? | ○ | ○ | ○ | ○ | ○ | ○ | ○ | ○ | ○ | ○ | ○ |
| 11. Your feeling **sad** at its WORST? | ○ | ○ | ○ | ○ | ○ | ○ | ○ | ○ | ○ | ○ | ○ |
| 12. Your **vomiting** at its WORST? | ○ | ○ | ○ | ○ | ○ | ○ | ○ | ○ | ○ | ○ | ○ |
| 13. Your **numbness or tingling** at its WORST? | ○ | ○ | ○ | ○ | ○ | ○ | ○ | ○ | ○ | ○ | ○ |
| 14. Your **weakness** on one side of the body at its WORST? | ○ | ○ | ○ | ○ | ○ | ○ | ○ | ○ | ○ | ○ | ○ |
| 15. Your difficulty **understanding** at its WORST? | ○ | ○ | ○ | ○ | ○ | ○ | ○ | ○ | ○ | ○ | ○ |
| 16. Your difficulty **speaking** (finding the words) at its WORST? | ○ | ○ | ○ | ○ | ○ | ○ | ○ | ○ | ○ | ○ | ○ |

SAMPLE

Copyright 2000 The University of Texas M. D. Anderson Cancer Center
All rights reserved.

MDASD-BT (page 2 of 2)

| | Not Present 0 | 1 | 2 | 3 | 4 | 5 | 6 | 7 | 8 | 9 | As Bad As You Can Imagine 10 |
|---|---|---|---|---|---|---|---|---|---|---|---|
| 17. Your **seizures** at its WORST? | ○ | ○ | ○ | ○ | ○ | ○ | ○ | ○ | ○ | ○ | ○ |
| 18. Your difficulty **concentrating** at its WORST? | ○ | ○ | ○ | ○ | ○ | ○ | ○ | ○ | ○ | ○ | ○ |
| 19. Your **vision** at its WORST? | ○ | ○ | ○ | ○ | ○ | ○ | ○ | ○ | ○ | ○ | ○ |
| 20. Your change in **appearance** at its WORST? | ○ | ○ | ○ | ○ | ○ | ○ | ○ | ○ | ○ | ○ | ○ |
| 21. Your change in **bowel pattern** (diarrhea or constipation) at its WORST? | ○ | ○ | ○ | ○ | ○ | ○ | ○ | ○ | ○ | ○ | ○ |
| 22. Your **irritability** at its WORST? | ○ | ○ | ○ | ○ | ○ | ○ | ○ | ○ | ○ | ○ | ○ |

SAMPLE

**Part II. How have your symptoms interfered [illegible]ith [illegible] li[illegible]**

**Symptoms frequently interfere with h[illegible]w we [illegible] [illegible]nction. How much have your symptoms interfered with the following items in the last 2[illegible]:**

| | Di[illegible] not interfere 0 | 1 | 2 | 3 | 4 | 5 | 6 | 7 | 8 | 9 | Interfered Completely 10 |
|---|---|---|---|---|---|---|---|---|---|---|---|
| 23. **General activity?** | ○ | ○ | ○ | ○ | ○ | ○ | ○ | ○ | ○ | ○ | ○ |
| 24. **Mood?** | ○ | ○ | ○ | ○ | ○ | ○ | ○ | ○ | ○ | ○ | ○ |
| 25. **Work (including work around the house)?** | ○ | ○ | ○ | ○ | ○ | ○ | ○ | ○ | ○ | ○ | ○ |
| 26. **Relations with other people?** | ○ | ○ | ○ | ○ | ○ | ○ | ○ | ○ | ○ | ○ | ○ |
| 27. **Walking?** | ○ | ○ | ○ | ○ | ○ | ○ | ○ | ○ | ○ | ○ | ○ |
| 28. **Enjoyment of life?** | ○ | ○ | ○ | ○ | ○ | ○ | ○ | ○ | ○ | ○ | ○ |

Copyright 2000 The University of Texas M. D. Anderson Cancer Center
All rights reserved.

**Page 2 of 2**

ENGLISH

### EORTC QLQ-C30 (version 3)

We are interested in some things about you and your health. Please answer all of the questions yourself by circling the number that best applies to you. There are no "right" or "wrong" answers. The information that you provide will remain strictly confidential.

Please fill in your initials: |_|_|_|_|
Your birthdate (Day, Month, Year): |_|_|_|_|_|_|_|_|
Today's date (Day, Month, Year): 31 |_|_|_|_|_|_|_|_|

| | Not at All | A Little | Quite a Bit | Very Much |
|---|---|---|---|---|
| 1. Do you have any trouble doing strenuous activities, like carrying a heavy shopping bag or a suitcase? | 1 | 2 | 3 | 4 |
| 2. Do you have any trouble taking a long walk? | 1 | 2 | 3 | 4 |
| 3. Do you have any trouble taking a short walk outside of the house? | 1 | 2 | 3 | 4 |
| 4. Do you need to stay in bed or a chair during the day? | 1 | 2 | 3 | 4 |
| 5. Do you need help with eating, dressing, washing yourself or using the toilet? | 1 | 2 | 3 | 4 |

| **During the past week:** | Not at All | A Little | Quite a Bit | Very Much |
|---|---|---|---|---|
| 6. Were you limited in doing either your work or other daily activities? | 1 | 2 | 3 | 4 |
| 7. Were you limited in pursuing your hobbies or other leisure time activities? | 1 | 2 | 3 | 4 |
| 8. Were you short of breath? | 1 | 2 | 3 | 4 |
| 9. Have you had pain? | 1 | 2 | 3 | 4 |
| 10. Did you need to rest? | 1 | 2 | 3 | 4 |
| 11. Have you had trouble sleeping? | 1 | 2 | 3 | 4 |
| 12. Have you felt weak? | 1 | 2 | 3 | 4 |
| 13. Have you lacked appetite? | 1 | 2 | 3 | 4 |
| 14. Have you felt nauseated? | 1 | 2 | 3 | 4 |
| 15. Have you vomited? | 1 | 2 | 3 | 4 |
| 16. Have you been constipated? | 1 | 2 | 3 | 4 |

Please go on to the next page

EORTC QLC-C30 (page 2 of 2)

ENGLISH

| During the past week: | Not at All | A Little | Quite a Bit | Very Much |
|---|---|---|---|---|
| 17. Have you had diarrhea? | 1 | 2 | 3 | 4 |
| 18. Were you tired? | 1 | 2 | 3 | 4 |
| 19. Did pain interfere with your daily activities? | 1 | 2 | 3 | 4 |
| 20. Have you had difficulty in concentrating on things, like reading a newspaper or watching television? | 1 | 2 | 3 | 4 |
| 21. Did you feel tense? | 1 | 2 | 3 | 4 |
| 22. Did you worry? | 1 | 2 | 3 | 4 |
| 23. Did you feel irritable? | 1 | 2 | 3 | 4 |
| 24. Did you feel depressed? | 1 | 2 | 3 | 4 |
| 25. Have you had difficulty remembering things? | 1 | 2 | 3 | 4 |
| 26. Has your physical condition or medical treatment interfered with your family life? | 1 | 2 | 3 | 4 |
| 27. Has your physical condition or medical treatment interfered with your social activities? | 1 | 2 | 3 | 4 |
| 28. Has your physical condition or medical treatment caused you financial difficulties? | 1 | 2 | 3 | 4 |

**For the following questions please circle the number between 1 and 7 that best applies to you**

29. How would you rate your overall health during the past week?

| 1 | 2 | 3 | 4 | 5 | 6 | 7 |
|---|---|---|---|---|---|---|
| Very poor | | | | | | Excellent |

30. How would you rate your overall quality of life during the past week?

| 1 | 2 | 3 | 4 | 5 | 6 | 7 |
|---|---|---|---|---|---|---|
| Very poor | | | | | | Excellent |

EORTC QLQ - PAN26

Patients sometimes report that they have the following symptoms or problems. Please indicate the extent to which you have experienced these symptoms or problems during the past week. Please answer by circling the number that best applies to you.

| During the past week: | Not at all | A little | Quite a bit | Very much |
|---|---|---|---|---|
| 31. Have you had abdominal discomfort? | 1 | 2 | 3 | 4 |
| 32. Did you have a bloated feeling in your abdomen? | 1 | 2 | 3 | 4 |
| 33. Have you had back pain? | 1 | 2 | 3 | 4 |
| 34. Did you have pain during the night? | 1 | 2 | 3 | 4 |
| 35. Did you find it uncomfortable in certain positions (e.g. lying down)? | 1 | 2 | 3 | 4 |
| 36. Were you restricted in the types of food you can eat as a result of your disease or treatment? | 1 | 2 | 3 | 4 |
| 37. Were you restricted in the amounts of food you could eat as a result of your disease or treatment? | 1 | 2 | 3 | 4 |
| 38. Did food and drink taste different from usual? | 1 | 2 | 3 | 4 |
| 39. Have you had indigestion? | 1 | 2 | 3 | 4 |
| 40. Were you bothered by gas (flatulence)? | 1 | 2 | 3 | 4 |
| 41. Have you worried about your weight being too low? | 1 | 2 | 3 | 4 |
| 42. Did you feel weak in your arms and legs? | 1 | 2 | 3 | 4 |
| 43. Did you have a dry mouth? | 1 | 2 | 3 | 4 |
| 44. Have you had itching? | 1 | 2 | 3 | 4 |
| 45. To what extent was your skin yellow? | 1 | 2 | 3 | 4 |
| 46. Did you have frequent bowel movements? | 1 | 2 | 3 | 4 |
| 47. Did you feel the urge to move your bowels quickly? | 1 | 2 | 3 | 4 |
| 48. Have you felt physically less attractive as a result of your disease and treatment? | 1 | 2 | 3 | 4 |

Please go to the next page

Specimen

EORTC-PAN26 (page 2 of 2)

| During the past week: | Not at all | A little | Quite a bit | Very much |
|---|---|---|---|---|
| 49. Have you been dissatisfied with your body? | 1 | 2 | 3 | 4 |
| 50. To what extent have you been troubled with side-effects from your treatment? | 1 | 2 | 3 | 4 |
| 51. Were you worried about your health in the future? | 1 | 2 | 3 | 4 |
| 52. Were you limited in planning activities, for example meeting friends, in advance? | 1 | 2 | 3 | 4 |
| 53. Have you received adequate support from your health care professionals? | 1 | 2 | 3 | 4 |
| 54. Has the information given about your physical condition and treatment been adequate? | 1 | 2 | 3 | 4 |
| 55. Have you felt less interest in sex? | 1 | 2 | 3 | 4 |
| 56. Have you felt less sexual enjoyment? | 1 | 2 | 3 | 4 |

Specimen

© QLQ-C30-PAN26 Copyright 1999 EORTC Study Group on Quality of life. All rights reserved (phase III module)

ENGLISH

### EORTC QLQ - BN20

Patients sometimes report that they have the following symptoms. Please indicate the extent to which you have experienced these symptoms or problems during the past week.

| During the past week: | Not at All | A Little | Quite a Bit | Very Much |
|---|---|---|---|---|
| 31. Did you feel uncertain about the future? | 1 | 2 | 3 | 4 |
| 32. Did you feel you had setbacks in your condition? | 1 | 2 | 3 | 4 |
| 33. Were you concerned about disruption of family life? | 1 | 2 | 3 | 4 |
| 34. Did you have headaches? | 1 | 2 | 3 | 4 |
| 35. Did your outlook on the future worsen? | 1 | 2 | 3 | 4 |
| 36. Did you have double vision? | 1 | 2 | 3 | 4 |
| 37. Was your vision blurred? | 1 | 2 | 3 | 4 |
| 38. Did you have difficulty reading because of your vision? | 1 | 2 | 3 | 4 |
| 39. Did you have seizures? | 1 | 2 | 3 | 4 |
| 40. Did you have weakness on one side of your body? | 1 | 2 | 3 | 4 |
| 41. Did you have trouble finding the right words to express yourself? | 1 | 2 | 3 | 4 |
| 42. Did you have difficulty speaking? | 1 | 2 | 3 | 4 |
| 43. Did you have trouble communicating your thoughts? | 1 | 2 | 3 | 4 |
| 44. Did you feel drowsy during the daytime? | 1 | 2 | 3 | 4 |
| 45. Did you have trouble with your coordination? | 1 | 2 | 3 | 4 |
| 46. Did hair loss bother you? | 1 | 2 | 3 | 4 |
| 47. Did itching of your skin bother you? | 1 | 2 | 3 | 4 |
| 48. Did you have weakness of both legs? | 1 | 2 | 3 | 4 |
| 49. Did you feel unsteady on your feet? | 1 | 2 | 3 | 4 |
| 50. Did you have trouble controlling your bladder? | 1 | 2 | 3 | 4 |

SPECIMEN

#### Appendix 3: Partner Organizations

**Role of Partner Organizations: Partner organizations are those for which xCures provides services under contract and who may refer patients to the XCELSIOR registry. Partners may or may not contribute to the analysis, publication, or dissemination of research findings. Each partner that will participate in research activities will designate a Principal Investigator to represent their organization in any research-related activities. These personnel will be required to submit a CV and evidence of suitable research and data privacy training.**

**Organization: Pediatric Oncology Experimental Therapeutics Investigators Consortium (POETIC)**

Web site: https://www.poeticphase1.org/

Principal Investigator (PI): Norman J. Lacayo, MD
Title: Associate Professor Pediatrics, Laurie Kraus Lacob Faculty Scholar in Pediatric Translational Medicine, Oncology Section and Clinical Research Group Interim Leader, POETIC Consortium Director, POETIC Data and Coordination Center at Stanford University
Affiliation: Bass Center for Childhood Cancer and Blood Disorders at Lucile Packard Children's Hospital, Division of Hematology-Oncology, School of Medicine,
Address: Stanford Cancer Institute at Stanford University
1000 Welch Rd, Suite 300
Palo Alto, CA 94304-1812


Research Coordinator: Ativ Zomet, PhD
Title: Pediatrics Hem Onc, POETIC Research Program Manager

POETIC DCC: 650-736-0269
Pager #: 16163
Address: 1000 Welch Road, Suite 300
Palo Alto, CA 94304.

**Organization: Musella Foundation for Brain Tumor Research & Information, Inc.**

Web site: https://virtualtrials.org/musella.cfm

Principal Investigator (PI): Al Musella, DPM
Title: President, Al Musella Foundation

Address: 1100 Peninsula Blvd.
Hewlett, NY 11557

**Organization: Cancer Commons**

Web site: https://cancercommons.org/home/

Principal Investigator (PI): Lola Rahib, PhD
Title: Director of Scientific & Clinical Affairs

Address: 650 Castro Street, Suite 120-522
Mountain View, CA 94041
