## Supplement 3: Fictitious Treatment Options Report for "AI-Augmented Clinical Decision Support in a Patient-Centric Precision Oncology Registry"

xCures

**Sam Larson** 21 Jan 1979

### Treatment Options

**Report Date** 01 Jan 2022

**What is this report?**
This report provides a list of treatment options for you and your treating oncologist to review. This report does not constitute a medical treatment plan and is not a substitute for medical advice. However, it is intended to be actionable information for you to understand and discuss with your treating oncologist.

**How was this report created?**
This report was created by a PhD scientist after reviewing your Cancer Journey, our understanding of your cancer history, based on your medical records. This may include notes from your physician(s), test results, imaging scans, genomic sequencing, and/or molecular profiling. This list of treatment options is based on the latest clinical evidence, clinical trials, and learnings from how other patients are doing in our platform based on the treatment options they have pursued.

*If your treating oncologist would like help with access to these treatment options please contact us at.*

### Your Treatment Options

1 **Bevacizumab, Lomustine**

**Access Mechanism**
Standard of Care

**Why**
Continue on current treatment regimen of bevacizumab and lomustine as most recent scan shows improvement.

2 **Bevacizumab, Lomustine, NovoTTF-100A Device**

**Access Mechanism**
Standard of Care

**Why**
Continue on current treatment regimen of bevacizumab and lomustine as most recent scan shows improvement. Consider adding the NovoTTF-100A Device (Optune) given frontopariental tumor location. This is a complete standard-of-care regimen for recurrent glioblastoma.

3 **Lenvatinib, Pembrolizumab**

**Access Mechanism**
Off-Label

**Why**
In case of disease progression, consider the combination of the immune checkpoint inhibitor pembrolizumab with the kinase inhibitor lenvatinib. In a Phase 2 trial of pembrolizumab and lenvatinib in patients with advanced solid tumors, the overall response rate in the 31 included patients with GBM was 16.1% (https://ascopubs.org/doi/10.1200/JCO.19.02627, https://pubmed.ncbi.nlm.nih.gov/32716739/, https://ecancer.org/en/news/18667-esmo-2020-first-time-data-revealed-from-two-studies-evaluating-pembrolizumab-plus-lenvatinib-in-seven-different-tumour-types).

4 **Bi-functional Alkylating Agent VAL-083**

**Access Mechanism**
Expanded Access

**Why**
In case of disease progression, consider VAL-083 given unmethylated MGMT promoter. In 72 patients with MGMT-unmethylated recurrent GBM treated with VAL-083, median overall survival time was 7.1 months. (https://www.biospace.com/article/releases/delmar-pharmaceuticals-presents-positive-interim-data-on-val-083-demonstrating-favorable-outcomes-in-both-newly-diagnosed-and-recurrent-gbm-at-the-aacr-virtual-annual-meeting-ii/). Access via expanded access trial (https://clinicaltrials.gov/ct2/show/NCT03138629).

5 **Bevacizumab, Irinotecan, NovoTTF-100A Device, Temozolomide**

**Access Mechanism**
Off-Label

**Why**
Consider combination treatment regimen of standard of care options bevacizumab, temozolomide, and NovoTTF-100A Device (Optune) with the off-label option irinotecan. A Phase 2 trial of 41 recurrent glioblastoma patients treated with bevacizumab, irinotecan, and temozolomide reported a partial radiographic response in 9 patients and stable disease in 25 patients (https://pubmed.ncbi.nlm.nih.gov/26025933/). A retrospective analysis of 48 recurrent glioblastoma patients reported increased median overall survival and progression-free survival with bevacizumab, temozolomide, irinotecan, and Optune as compared with bevacizumab alone or in combination with either irinotecan or lomustine (https://www.ncbi.nlm.nih.gov/pmc/articles/PMC6366009/).

### Cancer Journey

**Last Modified** 25 Dec 2021

My health

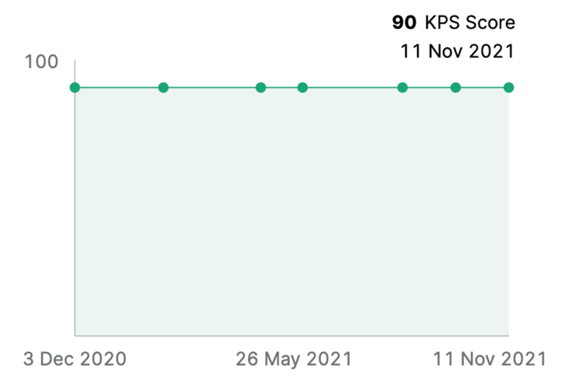


My Cancer

**09 Sep 2019 (Latest)**
**Diagnosis:** Glioblastoma
**Stage/Grade:** WHO Grade IV
**Location(s):** Brain, Right frontoparietal lobe

Biomarkers

| DATE | GENE | ALTERATION |
|---|---|---|
| 05 Jul 2021 | GFAP | Positive |
| 05 Jul 2021 | MGMT | Unmethylated |
| 05 Jul 2021 | IDH-1 R132H | Negative |
| 05 Jul 2021 | MSI | Negative |
| 05 Jul 2021 | TERT | C228T |
| 05 Jul 2021 | Tumor Mutational Burden | 1.6 mutations/MB |
| 05 Jul 2021 | p53 | Nuclear staining in a few scattered cells |
| 05 Jul 2021 | EGFR | Positive |
| 05 Jul 2021 | EGFR | Ser768Ile |
| 05 Jul 2021 | Ki-67 | 14.8% |
| 09 Sep 2019 | p53 | <5% |
| 09 Sep 2019 | ATRX | Preserved in majority of tumor cells |
| 09 Sep 2019 | EGFR | Missense mutation |
| 09 Sep 2019 | IDH1 R132H | Negative |
| 09 Sep 2019 | IDH2 | Negative |
| 09 Sep 2019 | Ki-67 | 50% |
| 09 Sep 2019 | MGMT | Unmethylated |
| 09 Sep 2019 | 1p/19q co-deletion | Negative |

**Treatment**
**Current treatment**

- **Lomustine (started 10/2/2021)**
200mg every 6 weeks
- **Valacyclovir (started 9/30/2021)**
500mg BID
- **Bevacizumab (started 9/30/2021)**
15 mg/kg every 3 weeks
- **Quercetin (started 7/31/2021)**
- **Metformin (started 5/14/2021)**
500 mg
- **Mebendazole (started 5/14/2021)**
200mg QD
- **Doxycycline (started 5/14/2021)**
100mg QD 3 months on 30 days off
- **Atorvastatin (started 5/14/2021)**
40 mg

Historical Surgeries and Therapies

**02 Oct 2021**
**Lomustine**
200mg every 6 weeks

**30 Sep 2021**
**Valacyclovir**
500mg BID

**30 Sep 2021**
**Bevacizumab**
15 mg/kg every 3 weeks

**01 Sep 2021 - 05 Sep 2021**
**Temozolomide**
150 mg/m2; cycle 13, stopped due to rapid tumor growth

**31 Jul 2021**
**Quercetin**

**08 Jul 2021 - 19 Jul 2021**
**Dexamethasone**
2mg BID

**05 Jul 2021 - 05 Jul 2021**
**Resection**
Right frontoparietal lobe

**14 May 2021**
**Metformin**
500 mg

**14 May 2021**
**Mebendazole**
200mg QD

**14 May 2021**
**Doxycycline**
100mg QD 3 months on 30 days off

**14 May 2021**
**Atorvastatin**
40 mg

**27 Jan 2020 - 11 Jan 2021**
**Temozolomide**
150 mg/m2; 12 cycles

**29 Oct 2019 - 10 Dec 2019**
**Temozolomide**

**29 Oct 2019 - 10 Dec 2019**
**Radiation Therapy**
Brain

**14 Oct 2019 - 16 Apr 2020**
**Bevacizumab**
10 mg/kg; NCT01269853

**11 Sep 2019 - 11 Oct 2019**
**Dexamethasone**
2mg

**09 Sep 2019 - 09 Sep 2019**
**Gross Total Resection**
Right frontoparietal lobe
